# Higher Nocturnal Temperatures Shorten and Worsen Sleep with No Clear Evidence of Short-Term Compensatory Patterns

**DOI:** 10.64898/2026.11.02.26367043

**Authors:** Christine Blume, Davide Mazorati, Tiziano Gerosa, Isabel Baumann, Remo Fortunato, Martina S. Ragettli

**Affiliations:** Swiss Tropical and Public Health Institute, Allschwil, Switzerland; University of Basel, Basel, Switzerland; Centre for Chronobiology, Psychiatric Hospital of the University of Basel, Basel, Switzerland; Institute of Digital Technologies for Personalized Healthcare, Department of Innovative Technologies, University of Applied Sciences and Arts of Southern Switzerland, Lugano, Switzerland; Institute of Applied Sustainability to the Built Environment, University of Applied Sciences and Arts of Southern Switzerland, Mendrisio, Switzerland; Institute of Public Health, ZHAW Zurich University of Applied Sciences, Winterthur, Switzerland

## Abstract

**Background & Objectives:** Rising nighttime temperatures menace sleep, yet evidence from large longitudinal studies combining objective and subjective sleep measures remains limited. Here, we therefore examined associations of nocturnal air and wet-bulb temperature with seven sleep parameters, including possible delayed effects.

**Methods:** We analysed wearable-derived and subjective sleep data from 326 home-care workers (270 women) in Switzerland between June and Early October 2025, resulting in 14,290 participant-nights. Nocturnal air temperature and humidity data were obtained from the dense MeteoSwiss monitoring network (mean distance from participants’ homes, 8.6 km). Bayesian multilevel models estimated nonlinear same-night within-person and between-person associations, while exploratory lag models examined whether associations with nocturnal temperature were also evident for exposures one and two nights earlier.

**Results:** Sleep maintenance efficiency, the primary outcome, showed no meaningful association with nocturnal air temperature. In contrast, warmer-than-usual nights were associated with shorter total sleep time and poorer subjective sleep quality. Compared with a participant’s usual exposure, total sleep time was 13.3 min shorter at +5°C, while subjective sleep-quality scores worsened by 1.2 points. At +5°C, the probability of sleeping less than 7 h was 10.6 percentage points higher than at-4°C relative to a participant’s usual exposure. Changes in sleep-stage percentages and recovery index were very small. Wet-bulb results were broadly comparable while exploratory lag analyses provided little evidence of consistent delayed effects.

**Conclusions:** Higher nocturnal ambient temperatures were associated with shorter and poorer sleep, with no evidence of short-term compensatory patterns. The results thus underline that sleep disturbances may represent an important pathway linking warming nights to human health.

## Introduction

High nocturnal temperatures are a well-known challenge for human sleep worldwide. When the Lancet Countdown report on health and climate change adopted “sleep hours lost due to high temperatures” as a health indicator, scientists showed that, compared to a 1986-2005 baseline, the sleep hours lost had increased by another record of 9 % in 2024 (+3 % from 2023; Romanello et al., 2025) and remained at a substantially elevated level compared to the historical baseline in 2025 (Romanello et al., 2026). Beyond the immediate effects on sleep, heat also contributes to increasing morbidity and mortality, where a lack of nocturnal rest and recovery may well play a role (Ebi et al., 2021; Liu et al., 2021; Ragettli et al., 2024; Saucy et al., 2021), although the precise contributions of sleep loss remain to be investigated.

The main mechanism by which high nocturnal ambient temperatures impair sleep is by hampering thermoregulation. Human body temperature follows a circadian rhythm (i.e., period length of ≈ 24 hours) peaking in the early evening hours and reaching its nadir in the early morning (Kräuchi, 2002). To achieve these temperature variations, the body balances heat production with heat loss, where the skin at hands and feet are particularly important (Aschoff, 1956; Kräuchi & Wirz-Justice, 2001). The relationship between heat loss and sleep is a close one, where sleep is typically initiated when body heat loss is maximal and core body temperature (CBT) drops most quickly (Kräuchi, 2007; Kräuchi et al., 2000; Murphy & Campbell, 1997). Sleep itself further supports the decrease of CBT (Barrett et al., 1993) and lower CBT has been associated with sleep improvements (Herberger et al., 2019; Kräuchi et al., 2018). Thus, if heat loss is impaired, sleep onset is delayed (Kräuchi et al., 1999). Likewise, high ambient temperature impairs heat dissipation thus undermining thermoregulation and consequently impacting on sleep (Chevance et al., 2024), particularly when behavioural thermoregulatory strategies, such as reducing the body surface covered by a blanket, are insufficient to offset the heat load (Ioannou et al., 2024).

Laboratory studies usually investigating the effects of heat on sleep in young volunteers using gold-standard sleep assessment with polysomnography (PSG) consistently found that heat increased nocturnal wakefulness thereby reducing sleep efficiency, that is the percentage of time in bed spent asleep (Ioannou et al., 2024; Libert et al., 1988; Okamoto-Mizuno et al., 2005; Tsuzuki et al., 2004). Furthermore, high ambient temperatures decreased rapid eye-movement (REM) sleep (Libert et al., 1988) and deep non-REM sleep proportions (Bonegio et al., 1988; Lan et al., 2014; Okamoto-Mizuno et al., 2005; Tsuzuki et al., 2004), increased sleep fragmentation (Libert et al., 1988), and delayed sleep onset latency (Lan et al., 2014). High ambient temperatures were also linked to worse subjective sleep quality (Lan et al., 2014).

Naturalistic studies have assessed the effects of seasonal temperature variations on sleep using mostly actimetry, that is an assessment of rest-activity patterns to estimate for instance sleep duration. More recently, consumer sleep-tracking devices have enabled the longitudinal assessment of objective sleep markers beyond just the duration of the resting phase in larger samples (Mattingly et al., 2021). In community-dwelling older adults followed across one year, increasing bedroom temperature from 20-25°C to 25-30°C was associated with meaningful reductions in sleep efficiency (i.e., the percentage of sleep among time spent in bed), sleep duration, and restfulness (Baniassadi et al., 2023). In a large international study using wearable-derived sleep data from more than 47,000 participants across 68 countries, Minor et al. (2022) found that higher minimal temperatures shortened sleep, primarily by delaying sleep onset. This effect was stronger in women and older adults, and the authors concluded there was no evidence of adaptation to warmer temperatures. However, exposure assessment was somewhat limited, the nearest weather station could be up to 100 km from an individual’s location, weather data were available only at daily resolution level, and subjective sleep information was not available.

Thus, there is a need for longitudinal studies combining objective and subjective sleep assessments with high-resolution environmental exposure data. Moreover, delayed effects of heat exposure on sleep, including possible compensatory responses across subsequent nights remain poorly characterised, although lagged effects have repeatedly been documented for other health outcomes such as mortality or hospital admissions (Bobb et al., 2014; Liu et al., 2025).

Therefore, we here sought to investigate concurrent and delayed associations between nocturnal heat exposure and objective and subjective sleep in healthcare workers providing home care in Switzerland. Additionally, we explored whether higher ambient temperatures, that is, air and wet-bulb temperature, affected mental wellbeing (Thompson et al., 2018; Thompson et al., 2023), for which sleep likely is a mediator (Freeman et al., 2020), and whether an app providing tips on coping with heat could reduce heat-related sleep disturbances (Gerosa et al., 2025).

To this end, we used a three-month longitudinal dataset collected from 342 home-care workers in Switzerland during the summer of 2025, including subjective and wearable-derived sleep measures alongside other assessments. The physically demanding nature of home care work and frequent travel between clients’ homes, in combination with increased workload during heat events, render this occupational group particularly vulnerable to the adverse effects of high temperatures (Fortunato et al., 2026). Although Switzerland is often perceived as an Alpine country characterised by glaciers, lakes, and mountain pastures, it is disproportionately affected by climate change and heat waves. Over recent decades, the number of tropical nights, defined as nights with temperatures ≥20°C, has increased in 18 of 26 Swiss cantons while the observed warming in Switzerland is approximately twice the global average (Fischer et al., 2022; MeteoSwiss & E.T.H. Zurich, 2025; Rippstein et al., 2023). The challenge is further compounded by building stock not designed for increasing temperatures and by the limited use of air conditioning, which remains uncommon and subject to regulatory restrictions (Mutschler et al., 2021). The summer of 2026 provided a stark illustration of the health risks due to heat: around 600 excess deaths were recorded in Switzerland between June and September, coinciding with repeated periods of extreme heat (Federal Statistical Office, 2026).

We hypothesised that Sleep Maintenance Efficiency (SME), our primary outcome, would decrease with increasing nocturnal temperature. Note that because the wearable device did not detect wakefulness before sleep onset or after final awakening, this measure reflects wakefulness during the sleep period rather than total sleep efficiency. Secondary (exploratory) outcomes were device-derived total sleep time, recovery index, percentages of light, deep, and REM sleep, and daily subjective sleep quality assessed with a sleep diary. We expected total sleep time, nocturnal recovery, deep, and REM sleep proportions as well as subjective sleep quality to decrease and light sleep proportions to increase with increasing temperatures. In exploratory analyses, we additionally examined whether temperatures were associated with sleep variables not only during the concurrent night, but also with temperatures on the preceding one and two nights. Beyond this, we hypothesised that higher ambient temperatures would negatively impact participants’ mental wellbeing and that the app-based intervention would ameliorate the effects of ambient temperature on sleep compared to the control group.

## Methods

### Participants

The full sample comprised 342 home-care workers from the German-and Italian-speaking parts of Switzerland, who were recruited to participate in the REHEAT study between June and Early October 2025 (for the published protocol, see Gerosa et al., 2025). The analyses presented here include data from 326 participants (270 women) with a mean age of 43.3 ± 11.4 years at the beginning of the study. Fifty percent of the sample had an 80 % employment or more (IQR 60-90 %; range 20-100 %). Only 10 participants (3.1 %) regularly worked night shifts, 31 (9.5 %) exceptionally worked during the night with all others not working night shifts. The majority of participants (*n* = 218; 67 %) worked in the canton of Ticino (i.e., southern Switzerland), and 108 participants (33 %) in northeastern Switzerland. Many were nurses (*n* = 185; 56.7 %), 71 (21.8 %) described themselves as care workers, 50 (15.3 %) had the role of a nursing assistant and 20 (6.1 %) indicated they had a different profession.

### Study procedures

Please note that the description here focuses on the measures relevant to this publication. The full protocol is described in Gerosa et al. (2025).

Following study inclusion, participants were assigned pseudorandomly to the intervention or control group in alternating fashion using a stratified random sampling approach with blocking by organization/geographical area (northeastern vs. southern Switzerland), age cohort (< 50 years, ≥ 50 years), and sex (male vs. female). Note that 8 participants were included only later and were therefore not randomised but treated like participants in the control group.

They were then familiarized with the REHEAT app, through which all questionnaires and the intervention were delivered. In brief, the intervention provided participants with practical tips on how to best manage nocturnal heat besides tips and notifications relating to self-monitoring of hydration and nutrition, sun protection, or clothing. For a concise overview of the tips and adaptation measures in the app, see Supplementary Table S1 and Gerosa et al. (2025). The control group was only granted access to the intervention module in the app after data collection had been completed.

An onboarding questionnaire assessed basic participant variables including sociodemographic variables, health status, and information relating to work (e.g., professional function, work schedules). It also served as a baseline assessment for subjective sleep quality – participants completed the brief version of the Pittsburgh Sleep Quality Index (B-PSQI; Sancho-Domingo et al., 2021). They also completed the Mental Health Index-5 (MHI-5; Rumpf et al., 2001). As the study was conducted in the German-and Italian-speaking parts of Switzerland, the app and all contents were provided in either German or Italian.

For the duration of the study, the participants received daily prompts to complete a diary to assess various symptoms. Here, they indicated whether they had worked the previous day, if yes, in which shift they worked (4-hour time slots from 7 am the previous day until 7 am that day). If they had not worked, they were asked to provide a reason (i.e., “Weekend, public holiday, or vacation”, “Illness”, “Injury”, “Other reason”). They also had to indicate whether they had slept at home and if they had worked the night. Note that we excluded days on which participants had worked a night shift (i.e., between 10 pm and 7 am) as well as the following night.

Daily subjective sleep quality was assessed with the Karolinska Sleep Diary (Aili et al., 2017), that is five questions each using a 5-point Likert scale: “How did you sleep?” (Likert scale: very poorly [5] - very well [1]), “Did you have restless sleep?” (Likert scale: not at all [1] – very restless [5]), “Did you have difficulties falling asleep?” (Likert scale: not at all [1] – very difficult [5]), “Did you have a premature awakening?” (Likert scale: not at all [1] – I woke up far too early [5]), “Do you feel refreshed today?” (Likert scale: not at all [5] – completely refreshed [5]). Once per week they completed the MHI-5.

### Wearable-Derived Sleep Metrics

For the duration of the study, participants continuously wore a Garmin Vivosmart 5 or Garmin Venu Sq 2 wearable device, from which we derived objective sleep metrics. They were instructed to wear the device as much as possible except for e.g., charging, sauna, or longer hot baths. Specifically, we looked at Sleep Maintenance Efficiency, which refers to the ratio between sleep duration and the time between the onset of sleep and the final awakening (primary endpoint). Note that Sleep Efficiency referring to the ratio between actual sleep duration and the time between lights off and getting up, which has been used in prior studies, is not provided by Garmin. Secondary outcomes included the percentage of light sleep (i.e., N1 and N2 according to scoring criteria of the American Association of Sleep Medicine [AASM]; Troester et al., 2023), percentage of deep sleep, and percentage of Rapid-Eye Movement (REM) sleep as well as a recovery index, which is the ratio between the actual increase (or decrease in some cases) in “body battery” overnight relative to the available increase. Thus, a recovery index of 100 % means all the available recharge was used. Recovery indices < 0 indicate that the body battery at the end of the night was lower than at the beginning. The “body battery” metric is a black-box algorithm by Garmin, which makes use of heart rate and heart rate variability data. Note that we are well-aware of the inaccuracies these data come with compared to sleep polysomnography (e.g., Chinoy et al., 2020).

### Ethical Approval

Ethical approval for the research project was provided by the cantonal ethics committee (Kantonale Ethikkommission Kanton Zürich; 2025-00561).

### Data Reduction and Analysis

#### Identification of nearest MeteoSwiss weather station

MeteoSwiss weather stations (Federal Office of & Climatology) were assigned based on participants’ geocoded home locations. Each participant was assigned to the geographically nearest eligible MeteoSwiss station, which had to provide hourly observations and be at an altitude of ≤1000 m a.s.l. to avoid assigning high-altitude stations unlikely to represent residential exposure conditions. In total, we identified 15 relevant stations and the mean distance between home addresses and the nearest MeteoSwiss station was 8.6 ± 5.1 km (range 0.7 - 23.8 km).

#### Meteorological exposure assessment

Hourly meteorological data were obtained from MeteoSwiss Open Data for the MeteoSwiss weather station assigned to each participants’ home location. Data were retrieved via the MeteoSwiss STAC API (ch.meteoschweiz.ogd-smn) for the study period from 6 June 2025 (first participant in) to 6 October 2025 (last participant out). From the hourly station files, air temperature at 2 m above ground (tre200h0) and relative humidity at 2 m above ground (ure200h0) were extracted.

Nocturnal exposure windows were defined in local time from 23:00 to 07:00. Each sleep-window date therefore comprised nine hourly observations: 23:00 on the evening of the sleep-window date and 00:00–07:00 on the following day or morning. For each station and sleep-window date, nocturnal mean air temperature was subsequently calculated from the hourly observations. In addition, hourly wet-bulb temperature was estimated from air temperature and relative humidity using the Stull approximation (Stull, 2011), and the nocturnal mean wet-bulb temperature was as the average across the sleep window. Wet-bulb temperature combines temperature and relative humidity and thus may provide a more accurate approximation of how human thermoregulation is affected by heat indoors (Maloney et al., 2024; McGregor & Vanos, 2018). Keeping temperature constant, a higher relative humidity is associated with an increased heat burden.

#### Heat episodes

Daily heat indicators were derived from station-level hourly data aggregated by local calendar day. The 24-h daily mean temperature (Tmean) identified warm-day sequences, while daily maximum temperature identified hot days (≥30°C), heat waves (≥3 consecutive days with Tmean ≥25°C), and hot spells (1-2 consecutive days with Tmean ≥25°C).

#### Data exclusion

Participants were excluded if participants reported pregnancy at baseline or offboarding (*n* = 6) or if the assigned MeteoSwiss station was more than 25 km from the home address (*n* = 8). Participant-nights were excluded if someone did not sleep at home or did not answer this question on a specific day (*n* = 11719). Additionally, nights for which participants reported evening or night work between 19:00 and 07:00 were excluded (*n* = 2264). We additionally excluded the nights following evening or night work (*n* = 1104) and finally excluded nights following a day on which participants reported illness (*n* = 267). This resulted in retention of 14290 nights (46.1 %) from 326 participants. Finally, Garmin recovery index data were only deemed valid after 7 nights of collected data.

#### Statistical Analyses Primary Sleep Analyses

For each sleep outcome, we fitted separate Bayesian multilevel models for two nocturnal heat-exposure metrics, i.e. mean air temperature and mean wet-bulb temperature. The primary same-night model was:

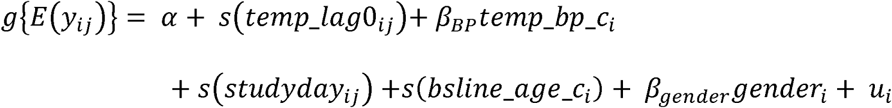

where y_ij denotes the sleep outcome for participant i on night j, and g is the outcome-specific link function. temp_lag0_ij denotes the same-night within-person deviation in the respective exposure from participant i’s mean exposure. Thus, a value of 0 represents a typical night for that participant, whereas positive and negative values represent warmer-and cooler-than-usual nights, respectively. temp_bp_c_i is the participant-level mean nocturnal air temperature or wet-bulb exposure, centred at the sample mean. study_day_ij accounts for temporal trends across the study period. bsline_age_c_i is the participant age at the beginning of the study, centred at the sample mean. gender_i is self-reported gender, and u_i is a participant-level random intercept.

The primary models therefore separated within-person from between-person exposure differences. The same-night within-person exposure term was modelled using a penalised smooth to allow for non-linear associations between temperature and sleep. Study day and baseline age were also included as smooth terms, whereas the between-person exposure component was modelled linearly.

For daily subjective sleep quality, baseline Brief Pittsburgh Sleep Quality Index (B-PSQI) score, centred at the sample mean, was included as an additional covariate:

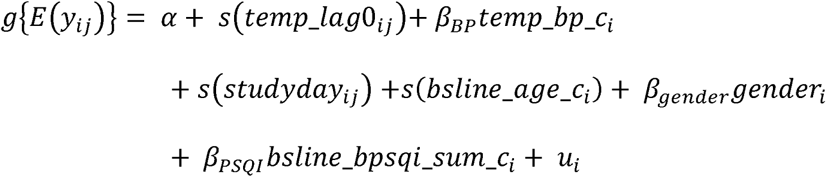

where bsline_bpsqi_sum_c_i is the B-PSQI score at the beginning of the study centred at the sample mean.

Sleep maintenance efficiency (SME) and the percentages of light, deep, and rapid eye movement sleep were modelled using beta regression after transformation to the open unit interval. Posterior predictions were obtained on the response scale, and contrasts for beta-regression outcomes were expressed as percentage-point differences. Total sleep time, recovery index, and daily subjective sleep quality were modelled using Gaussian likelihoods with identity links. Total sleep time was analysed in minutes. Recovery index was analysed on its native scale because it is not a strictly bounded proportion. Daily subjective sleep quality was analysed on its native scale and adjusted for baseline B-PSQI.

Posterior expected differences were estimated for within-person air or wet-bulb temperature deviations from-4°C to +5°C relative to 0°C, where 0°C represented a participant’s usual nocturnal air or wet-bulb temperature exposure. Participant-specific random effects were excluded from these predictions, yielding population-level expected contrasts. For sleep maintenance efficiency and sleep-stage proportions, contrasts are reported in percentage points; for total sleep time, recovery index, and subjective sleep quality, contrasts are reported in their native outcome units.

#### Exploratory and Supplementary Analyses

We conducted several additional analyses to examine potential lagged associations, analyse associations between temperature and weekly mental health, facilitate interpretation of the primary findings, and explore potential effect modification.

To explore potential carry-over associations from preceding nights, we fitted linear lag models that retained the covariate structure of the primary models but replaced the smooth same-night exposure term with separate linear within-person exposure terms for the outcome night and the preceding one or two nights. Specifically, we fitted models including lags 0-1 and lags 0-2:

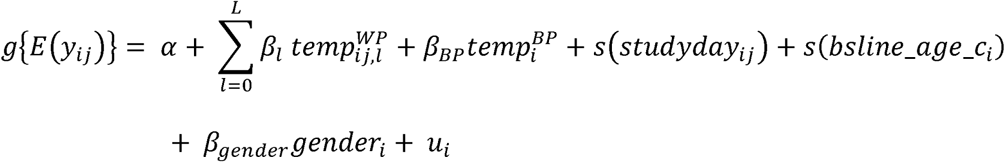

with L = 1 or 2. For the subjective sleep-quality outcome, the exploratory lag models additionally included baseline B-PSQI:

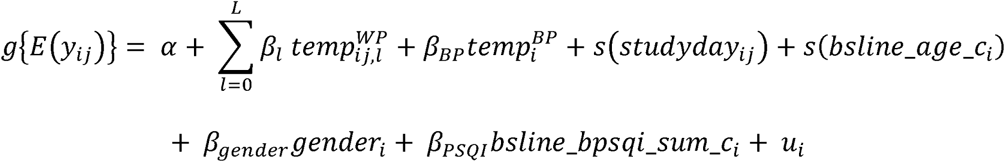

For lag l > 0, temp_lag_lij denotes the within-person deviation in nocturnal wet-bulb or air temperature l sleep nights before the outcome night. Lags were constructed using the sleep date, defined as the date on which the participant fell asleep. Lagged exposure values were retained only when the lagged sleep date was exactly l calendar days before the current sleep date; otherwise, the lagged value was set to missing. Lag models were therefore fitted to outcome-specific complete-case datasets with complete exposure histories for the respective lag window. Linear lag terms were used to limit model complexity and to screen for potential carry-over associations rather than to estimate non-linear distributed lag exposure-response functions.

Mental health was analysed separately with the MHI-5 at the participant-week level because it was assessed weekly rather than nightly. For each weekly mental-health score, we calculated the mean air temperature and mean wet-bulb temperature across the preceding seven-night exposure window. Exposure summaries required at least four valid nights. We fitted two separate Bayesian multilevel Gaussian models with identity links, each including a penalised smooth of one seven-night exposure metric and a participant-specific random intercept. Lagged mental-health models were not fitted.

To facilitate interpretation in terms of absolute temperature, we additionally fitted models using absolute mean nocturnal air temperature centred at 17°C, while retaining the remaining covariates and participant-specific random intercept of the primary same-night models. Posterior expected sleep outcomes were estimated from 11°C to 24°C, with contrasts calculated relative to 17°C. To further illustrate the potential impact of temperature-related changes in total sleep time, we estimated the nightly probability of obtaining less than 7 h or less than 6 h of sleep across the same absolute temperature range using Bayesian multilevel Bernoulli models with logit links. Probabilities and contrasts were reported as percentages and percentage-point differences.

Exploratory heat-episode analyses were restricted to nights occurring during heat-episodes and used linear same-night models. Posterior expected differences represent the change in the sleep outcome associated with a 1°C higher within-person nocturnal temperature, conditional on the participant’s full-sample mean exposure and the remaining covariates.

We also explored whether the REHEAT intervention modified the association between nocturnal temperature and sleep. Analyses were restricted to participants in the Treatment and Control groups (*n* = 318 for subjective sleep quality; *n* = 301 for objective sleep measurements; *n* = 287 for recovery index). For each sleep outcome, separate models were fitted for mean nocturnal air temperature and nocturnal wet-bulb temperature. The primary same-night model was extended to allow the nonlinear within-person temperature-response association to differ between groups, while retaining the participant-level mean exposure, original covariates, and participant-specific random intercepts. Effect modification was quantified as the Treatment-minus-Control difference in posterior expected contrasts across within-person temperature deviations relative to each participant’s usual exposure.

Finally, we explored whether age modified the association between mean nocturnal air temperature and sleep. For each sleep outcome, the primary same-night model was extended by modelling within-person temperature deviation and age using a tensor-product smooth, while retaining the participant-level mean exposure and remaining covariates. Posterior temperature contrasts were estimated at the 25th, 50th, and 75th percentiles of the outcome-specific age distribution. Effect modification was summarised by comparing temperature contrasts between the 75th and 25th age percentiles.

All models were fitted in R version 4.5.1 using brms (Bürkner, 2021). Four Markov chains were run per model. Models used 6000 iterations per chain, including 3000 warm-up iterations, with adapt_delta = 0.995 and max_treedepth = 15. Model convergence and sampling quality were assessed using R, bulk and tail effective sample sizes, divergent transitions, and maximum treedepth warnings. Convergence and sampling quality were assessed using R, bulk and tail effective sample sizes, divergent transitions, and maximum tree depth. For the primary models, these diagnostics were evaluated jointly, and isolated divergent transitions were examined and reported rather than used as an automatic exclusion criterion. Exploratory lag models were treated as directional screening analyses and interpreted cautiously on the basis of posterior estimates, 95% credible intervals, and the posterior probability of the estimated direction.

## Results

### Participant-night sample and temperature exposure distribution

After application of exclusion criteria, the Bayesian analysis dataset comprised 14290 sleep episodes from 326 participants (270 women, 82.8 %). The mean age was 43.3 ± 11.4 years at the beginning of the study (range 18.6-65.3 years). Each participant contributed a median of 46 study nights (interquartile range [IQR] 28-62 nights; range 1-93 nights).

Outcome-specific complete-case datasets differed because of missingness in sleep, diary, or recovery-index variables. For the primary same-night models, the complete-case datasets included 326 participants and 14290 nights for subjective sleep quality, 309 participants and 11547 nights for sleep maintenance efficiency, total sleep time, and the percentages of deep, light, and REM sleep, and 294 participants and 10140 nights for recovery index. The weekly mental health analysis included 318 participants and 2064 assessments. For an overview, see Table 1. Note that analyses of temperature exposure on the preceding one or two nights required complete data across consecutive nights and therefore included fewer participants and nights.

**Table 1.** Descriptives for exposure and sleep variables as well as mental health.

|  | ***N* participants** | ***N* nights/values** | **min** | **5th percen-tile** | **max** | **mean** | **median** | **95th percen-tile** | **sd** |
|---|---|---|---|---|---|---|---|---|---|
| Nocturnal air temperature | 326 | 14290 | 8.8 | 12.3 | 25.8 | 17.4 | 17.5 | 22.6 | 3.3 |
| Nocturnal relative humidity (%) | 326 | 14290 | 33.3 | 57.4 | 100 | 83.2 | 87 | 96.6 | 13.0 |
| Nocturnal wet-bulb temperature | 326 | 14290 | 5.9 | 10.2 | 22.2 | 15.4 | 15.6 | 20.1 | 2.9 |
| Minimum nocturnal air temperature | 326 | 14290 | 6.2 | 10.7 | 24.6 | 16.0 | 16.2 | 21.2 | 3.2 |
| Subjective sleep quality (range 4-20, higher = worse) | 326 | 14290 | 4 | 6 | 20 | 7.3 | 7 | 20 | 4.2 |
| Total sleep time (min) | 309 | 11547 | 87 | 293 | 846 | 430.3 | 431 | 569 | 83.9 |
| Sleep maintenance efficiency | 309 | 11547 | 46.3 | 86.3 | 100 | 95.9 | 97.4 | 100 | 5 |
| Light sleep (%) | 309 | 11547 | 0 | 48.1 | 100 | 63.4 | 63.5 | 78.5 | 9.4 |
| Deep sleep (%) | 309 | 11547 | 0 | 8.8 | 100 | 19.2 | 18.2 | 32.99 | 7.7 |
| REM sleep (%) | 309 | 11547 | 0 | 4.3 | 44.8 | 17.4 | 17.6 | 28.6 | 7.2 |
| Recovery (%; i.e. actual recharge/available recharge) | 294 | 10140 | -540 | 19 | 100 | 55 | 54 | 99 | 25 |
| Mental health score (range 0-100) | 318 | 2064 | 16 | 48 | 100 | 74.8 | 76 | 96 | 15.2 |

Calculations included all available observations. The table reports the number of contributing participants and nights for each variable, followed by the observed mean, median, and standard deviation across all observations. Subjective sleep quality could range from 4-20 with higher values denoting worse sleep quality. Recovery is the actual recharge or overnight increase in body battery relative to the available increase (i.e., difference to 100). Note that participants were only asked to fill in the MHI-5 once per week, explaining the lower number of available values. We also examined extreme values and found no reason to assume they were not valid. sd = standard deviation.

Nocturnal wet-bulb temperature ranged from 5.9°C to 22.2°C, with a mean of 15.4 ± 2.9°C. Within-person same-night wet-bulb deviations ranged from-8.4°C to 6.6°C. The 5th (1st) and 95th (99th) percentiles were-4.3°C (-6.4°C) and 4.0°C (5.0°C), respectively. Nocturnal air temperature ranged from 8.8°C to 25.8°C (mean: 17.4 ± 3.3°C). Within-person same-night air temperature deviations ranged from - 7.0°C to 8.1°C. The 5th (1st) and 95th (99th) percentiles were-4.5°C (-5.7°C) and 4.5°C (5.8°C), respectively. Nocturnal humidity ranged from 33.3 to 100 % (mean: 83.2 ± 13 %). Accordingly, the posterior prediction range from-4°C to +5°C covered most within-person exposure deviations, extending from approximately the 5th to the 99th percentile, with +5°C representing a particularly warm night relative to an individual’s usual exposure. For an overview of the exposure distribution see Table 1.

### Descriptive statistics

Descriptive statistics were likewise calculated at the participant-night level using all available observations for each variable, without requiring complete lagged exposure data. Table 1 summarises the descriptive findings.

### Model diagnostics

Across the primary same-night models, effective sample sizes were adequate and R values were generally acceptable, with a maximum R of 1.01, minimum bulk effective sample size of 637, and minimum tail effective sample size of 1266. Some beta-regression models showed a small number of divergent transitions despite a high adapt_delta, with a maximum of 10 divergences in any primary model; no transitions reached the maximum treedepth. Inspection of posterior diagnostic plots did not indicate that divergences were concentrated in the main within-person wet-bulb terms. Full model-specific diagnostic summaries are provided in Supplementary Tables S2-S3.

Exploratory linear lag models were fitted to screen for potential carry-over associations from preceding nights. Most exploratory lag models showed adequate R and effective sample size values. Some beta-regression lag models showed a small number of divergent transitions despite the high adapt_delta; therefore, these exploratory results were interpreted cautiously and were not used as the primary basis for inference. Full diagnostics for the lag models are likewise provided in Supplementary Tables S2-S3.

### Primary same-night associations

The primary analyses estimated population-level expected differences in sleep outcomes for within-person deviations in same-night nocturnal mean air temperature, relative to a participant’s usual nocturnal air temperature exposure. Figure 1 shows posterior expected differences across the full range of modelled within-person air temperature deviations from-4°C to +5°C. The results are comparable to those found for nocturnal wet-bulb temperature, which we present in Supplementary Figure S1 and Supplementary Table S4.

**Figure 2.**
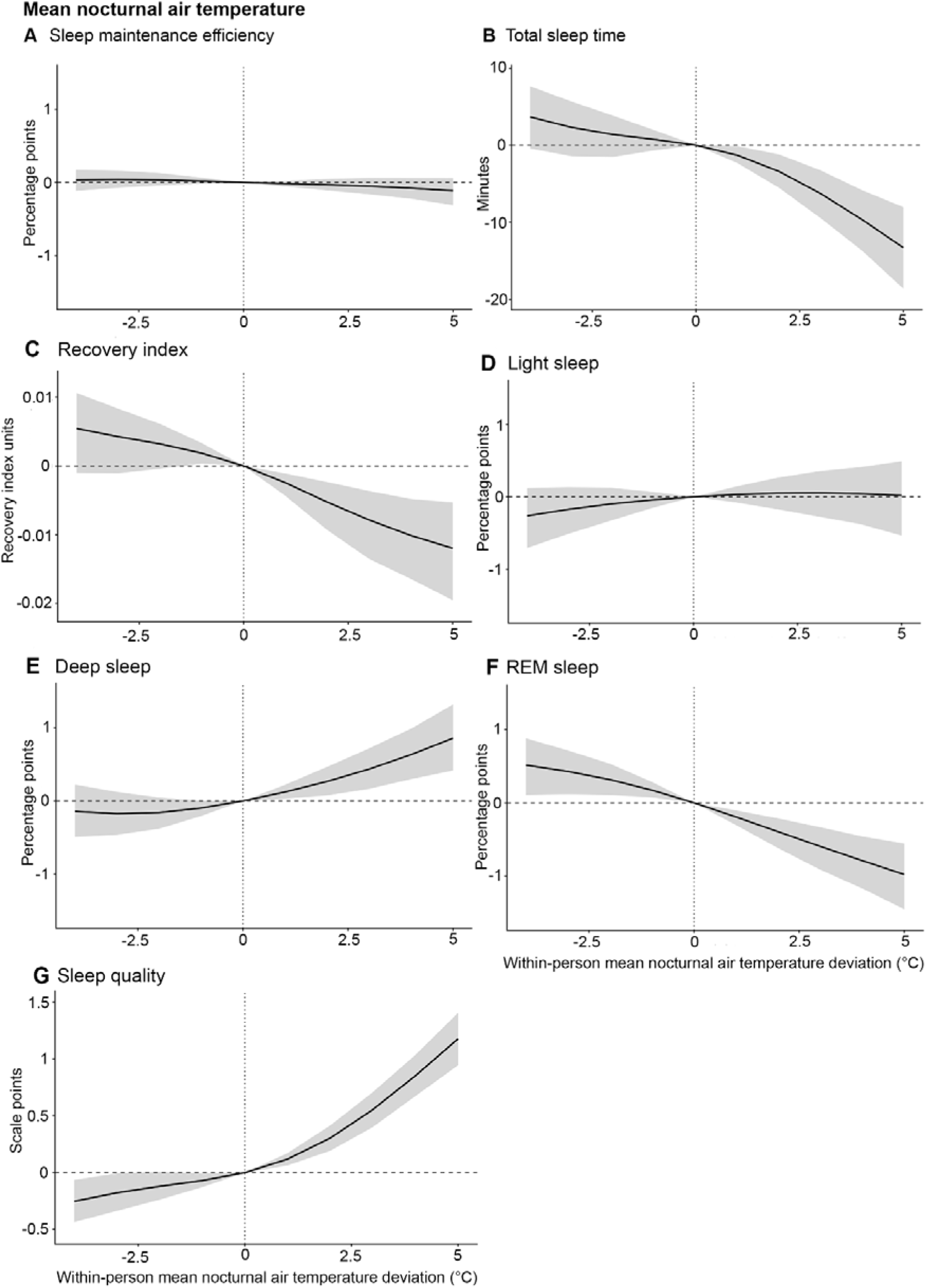
Primary same-night within-person associations between nocturnal air temperature and sleep outcomes. Posterior expected differences in (A) Sleep maintenance efficiency, (B) Total sleep time, (C) Recovery index, (D) Light sleep, (E) Deep sleep, (F) REM sleep, and (G) Subjective sleep quality corrected for baseline B-PSQI values are shown across within-person deviations in nocturnal air temperature from a participant’s usual exposure. Black lines indicate posterior mean differences from the 0°C reference, and shaded areas indicate 95% credible intervals. The vertical dashed line marks a participant’s usual nocturnal air exposure, and the horizontal dashed line marks no difference from this reference. Sleep maintenance efficiency and sleep-stage outcomes are expressed in percentage points; recovery index, total sleep time, and subjective sleep quality are expressed in their native outcome units. Higher subjective sleep-quality scores indicate worse sleep quality. For the results tables, please see Table 2 and Supplementary Table S5.

The primary same-night models showed little evidence of a meaningful association of warmer-than-usual nocturnal air temperature with sleep maintenance efficiency or light sleep. Both outcomes remained largely stable across the modelled exposure range, and estimated differences were negligible in magnitude. Selected posterior expected differences at +1°C, +3°C, and +5°C are presented in Table 2, with estimates across the full modelled range from-4°C to +5°C provided in Supplementary Table S5.

**Table 2.** Posterior expected differences in sleep outcomes on +1°C, +3°C, and +5°C warmer-than-usual nights.

| Outcome | Unit | +1°C | P(direction) | +3°C | P(direction) | +5°C | P(direction) |
|---|---|---|---|---|---|---|---|
| Sleep Maintenance Efficiency | Percentage points | -0.02 [-0.05, 0.02] | 0.84 | -0.05 [-0.16, 0.06] | 0.84 | -0.11 [-0.31, 0.06] | 0.90 |
| Total Sleep Time | Minutes | -1.3 [-2.31, -0.13] | 0.99 | -6.25 [-9.43, -3.25] | 1.00 | -13.29 [-18.6, -8.03] | 1.00 |
| Light Sleep | Percentage points | 0.03 [-0.07, 0.14] | 0.75 | 0.06 [-0.27, 0.35] | 0.66 | 0.02 [-0.53, 0.49] | 0.56 |
| Deep Sleep | Percentage points | 0.12 [0.03, 0.22] | 1.00 | 0.43 [0.16, 0.72] | 1.00 | 0.86 [0.42, 1.32] | 1.00 |
| REM Sleep | Percentage points | -0.19 [-0.3, -0.1] | 1.00 | -0.59 [-0.91, -0.33] | 1.00 | -0.98 [-1.45, -0.55] | 1.00 |
| Recovery Index | Recovery-index units | 0 [-0.01, 0] | 0.97 | -0.02 [-0.03, -0.01] | 1.00 | -0.02 [-0.04, -0.01] | 1.00 |
| Subjective Sleep Quality (BPSQI-adjusted) | Scale Points | 0.12 [0.06, 0.17] | 1.00 | 0.55 [0.4, 0.71] | 1.00 | 1.18 [0.95, 1.41] | 1.00 |

Values are posterior mean expected differences from a participant’s usual nocturnal air temperature exposure, with 95% credible intervals in brackets. For sleep maintenance efficiency and sleep-stage outcomes, values are percentage-point differences. Total sleep time is in minutes. For subjective sleep quality, higher values indicate worse sleep quality. P(direction) is the posterior probability that the expected difference is in the estimated direction.

By contrast, warmer-than-usual nights were associated with shorter total sleep time. Predicted total sleep time decreased from approximately 406 min on a participant’s usual night to 393 min at +5°C, corresponding to an estimated difference of-13.3 min (95% CrI [-18.60,-8.03]). Thus, although changes were modest at small temperature deviations, the estimated reduction became more substantial on markedly warmer-than-usual nights (Table 2; Supplementary Table S5). Recovery index also decreased with increasing temperature, with an estimated difference of-0.024 units at +5°C (95% CrI [-0.039,-0.011]). This corresponds to a small reduction in overnight recovery and was of minimal to modest practical relevance.

Sleep-stage proportions also varied with temperature, although the estimated changes were very small. Deep sleep increased slightly on warmer-than-usual nights, whereas REM sleep decreased. At +5°C, deep sleep was estimated to be 0.86 percentage points higher (95% CrI [0.42, 1.32]) and REM sleep 0.98 percentage points lower (95% CrI [-1.45,-0.55]) than on a participant’s usual night. Despite the consistent direction of these associations, their absolute magnitude was small and rather unlikely to be of practical relevance (Table 2; Supplementary Table S5).

Warmer-than-usual nocturnal air temperature was also associated with worse subjective sleep quality after adjustment for baseline B-PSQI. Predicted scores increased from approximately 11.4 on a participant’s usual night to 12.5 at +5°C, corresponding to an estimated difference of 1.18 scale points (95% CrI [0.95, 1.41]). Cooler-than-usual nights showed the opposite pattern. In contrast to the small changes observed for sleep architecture, the deterioration in subjective sleep quality became increasingly pronounced with larger positive temperature deviations (Table 2; Supplementary Table S5).

Exploratory analyses based on absolute mean nocturnal air temperature were consistent with the within-person analyses. Across temperatures from 11°C to 24°C, higher temperatures were associated with shorter total sleep time, lower recovery index and REM sleep, slightly higher deep sleep, and worse subjective sleep quality, whereas sleep maintenance efficiency and light sleep showed little variation. The largest and potentially most relevant differences were again observed for total sleep time and subjective sleep quality, whereas changes in sleep-stage composition and recovery index remained comparatively small (Supplementary Table S6; Supplementary Figure S2).

Sensitivity analyses restricted to heat episodes yielded broadly similar results. Higher nocturnal temperature was most consistently associated with shorter total sleep time and worse subjective sleep quality, whereas associations with sleep maintenance efficiency and light sleep remained small or uncertain and changes in sleep-stage composition and recovery index were generally modest (Supplementary Table S7-S8; Supplementary Figure S3).

### Probability of short sleep duration

Exploratory analyses showed an increasing probability of short sleep with higher nocturnal air temperature, both across absolute temperatures and on warmer-than-usual nights (Supplementary Table S9; Supplementary Figure S4). At 24°C compared with 17°C, the probability of sleeping <6 h was 5.9 percentage points higher (95% CrI [1.9, 10.6]) and the probability of sleeping <7 h was 8.7 percentage points higher (95% CrI [5.2, 12.6]); across 11°C to 24°C, the latter increased by approximately 15 percentage points. Similarly, at +5°C relative to a participant’s usual exposure, the probabilities of sleeping <6 h and <7 h were 4.9 (95% CrI [1.5, 9.0]) and 6.7 percentage points higher (95% CrI [3.9, 9.9]), respectively. Cooler conditions were associated with a lower probability of short sleep, particularly for the <7 h threshold.

### Between-person nocturnal exposure associations

Between-person associations showed a similar pattern for mean nocturnal air and wet-bulb temperature (Supplementary Tables S10-S11). Participants with higher mean nocturnal temperatures across the study period tended to have shorter total sleep time and lower recovery index values, whereas associations with sleep maintenance efficiency, sleep-stage proportions, and B-PSQI-adjusted subjective sleep quality were small or uncertain. In the primary same-night models, a 1°C higher mean nocturnal air temperature was associated with 3.76 min shorter total sleep time (95% CrI [-6.61,-1.02]) and a 0.03-unit lower recovery index (95% CrI [-0.04,-0.02]). Corresponding estimates for wet-bulb temperature were 5.47 min shorter total sleep time (95% CrI [-9.00,-1.92]) and a 0.03-unit lower recovery index (95% CrI [-0.04,-0.02]). Estimates were similar across the exploratory lag models.

### Additional covariate effects

Covariate associations were highly consistent across the air-temperature and wet-bulb models (Supplementary Tables S12-S13). In the primary same-night models, participants coded as female had approximately 29 min longer total sleep time and reported worse subjective sleep quality than the reference category, whereas gender differences in the remaining sleep outcomes were small. Higher baseline B-PSQI scores were consistently associated with worse daily subjective sleep quality.

Age showed outcome-specific, non-linear associations with sleep (Supplementary Figure S5; Supplementary Table S14). Increasing age was generally associated with lower sleep maintenance efficiency, total sleep time, recovery index, deep sleep, and REM sleep, and with a higher proportion of light sleep. Subjective sleep quality showed an inverted U-shaped association with age, with somewhat better ratings at younger and older ages. These covariate associations reflect overall between-participant differences, potential modification of the temperature-sleep association by age was examined separately.

### Exploratory lag analyses

Exploratory lag models included fewer nights and, for some outcomes, fewer participants because complete consecutive exposure histories were required (Supplementary Table S15). Concurrent air-temperature associations largely reproduced the primary same-night findings, with higher temperature associated with shorter total sleep time, lower recovery index, worse subjective sleep quality, and lower REM sleep, whereas associations with sleep maintenance efficiency and non-REM sleep stages were small or negligible (Supplementary Table S16; Supplementary Figure S6). Delayed associations at Lags 1 and 2 were generally small, inconsistent in direction across outcomes and lag windows, and provided little evidence of a systematic carry-over effect. Wet-bulb temperature showed a similar pattern, with clearer concurrent than delayed associations (Supplementary Table S17; Supplementary Figure S7).

### Weekly mental health analyses

Weekly mental-health scores showed no clear association with mean nocturnal temperature exposure during the preceding seven nights. Estimates for both air and wet-bulb temperature were generally compatible with little or no association across the modelled exposure range, although estimates for air temperature tended toward better mental-health scores at warmer-than-usual exposures (Supplementary Table S18).

### Effect of age

Exploratory analyses suggested limited age-related modification of the association between mean nocturnal air temperature and sleep. At +5°C relative to usual exposure, the temperature-related reduction in REM sleep was 1.49 percentage points greater at the 75th than at the 25th age percentile (95% CrI [-2.58,-0.34]), while the worsening in subjective sleep quality was 0.49 points greater (95% CrI [0.04, 0.92]). These differences were modest (REM sleep) to small (subjective sleep quality) in absolute magnitude. Age-modification estimates for total sleep time and the remaining outcomes were generally uncertain or inconsistent (Supplementary Table S19; Supplementary Figure S8).

### Effect of intervention group

There was no reliable evidence that group membership meaningfully modified the associations of mean nocturnal air or wet-bulb temperature with sleep outcomes. Credible intervals for all intervention-by-temperature interaction terms included zero (cf. Supplementary Figure S9 and Supplementary Table S20). Note that that the intervention and control groups generally did not differ on the sleep outcomes (cf. Supplementary Table 21).

## Discussion

Climate change poses growing risks to human health, and for sleep, nighttime temperatures are particularly relevant. As nights warm rapidly, rising temperatures may increasingly threaten sleep, an important determinant of health (Minor et al., 2026). Leveraging ambient meteorological data from the dense MeteoSwiss monitoring network, objective sleep data and subjective sleep ratings from up to 14,290 participant-nights, this study extends evidence that warmer nights are associated with shorter sleep duration, a higher probability of short sleep, and poorer subjective sleep quality. Cooler nights showed the opposite pattern, while exploratory analyses provided little evidence of short-term compensatory patterns.

More specifically, in our sample of home-care workers in northeastern and southern Switzerland, total sleep time was approximately 6 minutes shorter when nocturnal air temperature was 3°C higher than a participant’s usual exposure and about 13 minutes shorter at +5°C. We consider these differences meaningful, particularly as previous research suggests limited compensation for heat-related sleep loss on subsequent nights (Libert et al., 1988; Minor et al., 2022). At the same time, subjective sleep quality worsened by 0.6 scale points (range 4-20) at +3°C and by 1.2 scale points at +5°C with the deterioration slightly more pronounced in older individuals. This is consistent with previous findings suggesting that heat has a greater impact on sleep in older adults (Li et al., 2025; Minor et al., 2022). Notably, we found little indication that the intervention was able to attenuate these temperature-related changes. This suggests that practical tips (e.g., to take a lukewarm shower before going to bed, to ventilate the room when it is cool outside) alone may have limited potential to reduce the physiological sensitivity of sleep to high ambient temperatures. Exploratory analyses based on absolute nocturnal air temperature supported this pattern: Compared with 17°C, lower temperatures were associated with longer sleep duration and better subjective sleep quality, whereas higher temperatures were associated with shorter sleep duration and worse subjective sleep quality. Across the modelled range, predicted total sleep time decreased by almost 24 minutes from 11°C to 24°C, while subjective sleep quality worsened by approximately 1.8 scale points. Note that these temperatures refer to outdoor air temperatures with nocturnal indoor temperatures likely being even higher, especially during the warm season. Results for nocturnal wet-bulb temperature, which additionally accounts for relative humidity besides air temperature, were broadly comparable in magnitude.

The practical relevance of these results is underscored by the progressively increasing probability of short sleep at higher air temperatures, whether expressed as absolute air temperature or as warmer-than-usual nights. For example, the estimated probability of sleeping less than 7 h, a commonly used threshold for short sleep (Hirshkowitz et al., 2015), increased from about 53 % at 11°C to 68 % at 24°C, a difference of almost 15 percentage points. For the more stringent <6 h threshold, warmer nights were likewise associated with a higher probability of short sleep. Together, these findings suggest that even relatively modest temperature-related reductions in average sleep duration may translate into a meaningful increase in the frequency of nights falling below established sleep-duration thresholds.

Additionally, previous research provided little evidence for short-term compensatory responses or for medium-to long-term adaptation over months or years, suggesting that even moderate reductions in total sleep time may accumulate across repeated warm nights (Libert et al., 1988; Minor et al., 2022). Our exploratory lag analyses supported this interpretation. They assessed whether temperature on preceding nights was associated with sleep on the outcome night after accounting for temperature on that same night. We found little evidence of consistent delayed or compensatory patterns. Higher temperature on the outcome night was associated with shorter total sleep time and worse subjective sleep quality, whereas associations with temperatures on preceding nights were weaker and inconsistent. Some alternating patterns were observed for sleep-stage percentages and the recovery index, but these differences were negligible in magnitude. Overall, the findings provided little evidence of a systematic rebound following warmer nights. However, because the lag analyses examined prior-night temperature rather than prior-night sleep loss, they cannot directly determine whether sleep loss was compensated for on subsequent nights.

The temperature-related reduction in sleep duration observed in our study was larger than that reported by Minor et al. (2022). Linking more than 7 million wearable-derived sleep records from 68 countries to minimum nighttime temperature, they observed a considerably shallower decline in sleep duration, with their published response curve suggesting a reduction of approximately 5 minutes between 11°C and 24°C. Direct quantitative comparison is limited, however, because the studies used different temperature metrics, with mean nocturnal temperature in the present study and minimum daily temperature in Minor and colleagues’ study. Additionally, home care workers’ sleep may be particularly susceptible to heat, as workload often increases during heat episodes, adding psychological stress to the physical strain of their work (Fortunato et al., 2026). In a longitudinal dataset of adults aged over 65 years, the response curve reported by Baniassadi et al. (2023) suggests a decline of roughly 30 minutes in total sleep time between approximately 15°C and 25°C, which is more in line with our estimates. However, unlike the present study, the researchers measured indoor temperature directly in participants’ bedrooms, which limits direct comparison of effect sizes in addition to the participants’ age.

Unlike previous laboratory studies (Ioannou et al., 2024; Libert et al., 1988; Okamoto-Mizuno et al., 2005; Tsuzuki et al., 2004), we found no meaningful changes in sleep maintenance efficiency (SME) or sleep stage proportions with varying ambient temperatures, although temperature-related decreases in REM sleep were more pronounced in older individuals underlining their increased sensitivity to increasing nocturnal temperatures. The absence of clearer effects on sleep macrostructure may partly reflect the limited accuracy of consumer sleep-tracking devices, particularly for sleep-stage classification. An important difference is that previous studies assessed sleep efficiency, defined as total sleep time relative to the entire time spent in bed. In contrast, the wearable-derived SME used here only captures the proportion of sleep between the first and last detected sleep episodes and therefore excludes time spent awake before sleep onset or after final awakening. Additionally, while wearable devices are relatively good at detecting total sleep time, their ability to detect wakefulness after sleep onset is known to be limited (Chinoy et al., 2020). This could have resulted in an overestimation of SME and decreased sensitivity to changes in this metric. Further, consumer devices’ biases may have distorted night-to-night differences resulting, which could explain the absence of meaningful variations in sleep stage proportions with ambient temperature. Specifically, validation studies show that they often systematically overestimate wake after sleep onset (WASO) and light sleep on nights with little wakefulness and objectively better sleep. Conversely, they underestimate WASO and light sleep on ‘worse’ nights with more wakefulness (Chinoy et al., 2020; Kainec et al., 2024). This bias pattern can result in a regression towards the device’s mean estimate, and consequently an underestimation of inter-night variability.

The strengths of this study include the large number of participant-nights and the combination of wearable-derived objective sleep measures with subjective sleep ratings, providing complementary information on sleep under real-world conditions. The longitudinal within-person design allowed us to examine how sleep changed when individuals experienced warmer or cooler nights than usual, thereby reducing the influence of stable between-person differences. It also allowed us to explore possible delayed associations with temperature. Exposure assessment was based on hourly data from the dense MeteoSwiss monitoring network and included both air temperature and humidity, enabling us to additionally examine nocturnal wet-bulb temperature in addition to air temperature.

The main limitations concern exposure and sleep measurement. Weather-station data can only approximate the thermal conditions experienced in participants’ bedrooms. Particularly during warm periods, nighttime indoor temperatures likely remain higher than outdoor temperatures because of heat stored in buildings (Wright et al., 2005). Future studies should therefore directly record bedroom temperature and humidity, for example using iButtons (Maxim Integrated). Objective sleep outcomes were derived from consumer wearables, which are less accurate than polysomnography, particularly for sleep staging and the detection of wakefulness (Chinoy et al., 2020; Kainec et al., 2024). This may have limited our ability to detect small temperature-related changes in these outcomes. At the same time, we argue that consumer wearables provide a feasible and cost-efficient approach for obtaining repeated objective sleep measurements over thousands of nights under real-world conditions.

To conclude, in a large longitudinal study of home-care workers across northeastern and southern Switzerland, higher nocturnal ambient temperatures were associated with shorter total sleep time, a greater probability of short sleep, and poorer subjective sleep quality, with no clear evidence of temporal patterns consistent with short-term compensation. Besides direct physiological effects of heat, this could also reflect increased workload and stress due to high temperatures. Given the widespread exposure to rising nighttime temperatures worldwide and the importance of sufficient, good-quality sleep for health, these findings point to sleep as a potentially important and underrecognized pathway through which exposure to high ambient temperature may affect human health. Whether heat-related sleep impairment contributes to the broader adverse health effects of high temperatures, including excess mortality, hospital admissions, and poorer mental health, remains largely unknown and warrants further investigation.

## Funding

This research was funded by the Swiss National Science Foundation (SNSF; grant number 220630).

## Declaration of generative AI and AI-assisted technologies in the writing process

During the preparation of this manuscript, the authors used ChatGPT (OpenAI, <u>chatgpt.com</u>) to support coding task and for language polishing, including improving clarity, coherence, and wording. All outputs were critically reviewed and edited.

## Data Availability

All data produced in the present study are available upon reasonable request to the authors.

## Contributions

CB: Conceptualization of the present analyses, formal analysis, writing - original draft, writing - review & editing. DM: Data curation, writing - review & editing. RF: Writing - review & editing. TG: Conceptualization of the REHEAT project, investigation, data curation, writing - review & editing. IB: Funding acquisition, conceptualization of the REHEAT project, investigation, writing - review & editing. MSR: Funding acquisition, conceptualization of the REHEAT project and the present analyses, writing - review & editing.

### Competing Interests

CB has had the following commercial interests related to sleep and/or light: honoraria for invited talks and workshops from F.A. Hoffmann-La Roche AG, L’Oréal, Swissline Cosmetics, Ruby Hotels, Vattenfall, Swiss Post, and Idorsia Pharmaceuticals. CB is an elected member of the Daylight Academy.

## Data Availability

All data produced in the present study are available upon reasonable request to the authors.

## Notes

### Competing Interest Statement

The authors have declared no competing interest.

### Author Declarations

The cantonal ethics committee of the canton of Zurich gave ethical approval for this work (Kantonale Ethikkommission Kanton Zuerich; 2025-00561).

## Bibliography

Aili, K., Åström-Paulsson, S., Stoetzer, U., Svartengren, M., & Hillert, L. (2017). Reliability of Actigraphy and Subjective Sleep Measurements in Adults: The Design of Sleep Assessments. Journal of clinical sleep medicine, 13(1), 39–47. 10.5664/jcsm.6384

Aschoff, J. (1956). Wechselwirkungen zwischen Kern und Schale im Wärmehaushalt. Archiv Physikalische Therapie H, 3, 113–133.

Baniassadi, A., Manor, B., Yu, W., Travison, T., & Lipsitz, L. (2023). Nighttime ambient temperature and sleep in community-dwelling older adults. Science of The Total Environment, 899, 165623. 10.1016/j.scitotenv.2023.165623

Barrett, J., Lack, L., & Morris, M. (1993). The Sleep-Evoked Decrease of Body Temperature. Sleep, 16(2), 93–99. 10.1093/sleep/16.2.93

Bobb, J. F., Obermeyer, Z., Wang, Y., & Dominici, F. (2014). Cause-Specific Risk of Hospital Admission Related to Extreme Heat in Older Adults. Jama, 312(24), 2659–2667. 10.1001/jama.2014.15715

Bonegio, R. G., Driver, H. S., King, L. M., Laburn, H. P., & Shapiro, C. M. (1988). Circadian temperature rhythm blunting and sleep composition. Acta Physiol Scand Suppl, 574, 44–47.

Bürkner, P.-C. (2021). Bayesian Item Response Modeling in R with brms and Stan. Journal of Statistical Software, 100(5), 1–54. 10.18637/jss.v100.i05

Chevance, G., Minor, K., Vielma, C., Campi, E., O’Callaghan-Gordo, C., Basagaña, X., Ballester, J., & Bernard, P. (2024). A systematic review of ambient heat and sleep in a warming climate. Sleep medicine reviews, 75, 101915. 10.1016/j.smrv.2024.101915

Chinoy, E. D., Cuellar, J. A., Huwa, K. E., Jameson, J. T., Watson, C. H., Bessman, S. C., Hirsch, D. A., Cooper, A. D., Drummond, S. P. A., & Markwald, R. R. (2020). Performance of seven consumer sleep-tracking devices compared with polysomnography. Sleep, 44(5). 10.1093/sleep/zsaa291

Ebi, K. L., Capon, A., Berry, P., Broderick, C., de Dear, R., Havenith, G., Honda, Y., Kovats, R. S., Ma, W., Malik, A., Morris, N. B., Nybo, L., Seneviratne, S. I., Vanos, J., & Jay, O. (2021). Hot weather and heat extremes: health risks. The Lancet, 398(10301), 698–708. 10.1016/S0140-6736(21)01208-3

Federal Office of, M., & Climatology, M. Station information. MeteoSwiss. https://www.meteoswiss.admin.ch/climate/the-climate-of-switzerland/records-and-extremes/extreme-value-analyses/background-information/station-information.html

Federal Statistical Office. (2026). Mortality Monitoring. Federal Statistical Office. https://www.bfs.admin.ch/bfs/en/home/statistics/health/state-health/mortality-causes-death.html

Fischer, A. M., Strassmann, K. M., Croci-Maspoli, M., Hama, A. M., Knutti, R., Kotlarski, S., Schär, C., Schnadt Poberaj, C., Ban, N., Bavay, M., Beyerle, U., Bresch, D. N., Brönnimann, S., Burlando, P., Casanueva, A., Fatichi, S., Feigenwinter, I., Fischer, E. M., Hirschi, M.,…Zubler, E. M. (2022). Climate Scenarios for Switzerland CH2018 – Approach and Implications. Climate Services, 26, 100288. 10.1016/j.cliser.2022.100288

Fortunato, R., Ragettli, M. S., Gerosa, T., Dratva, J., & Baumann, I. (2026). Occupational heat strain among home health care workers: a cross-sectional study. BMC Public Health, 26(1), 2690. 10.1186/s12889-026-28969-8

Freeman, D., Sheaves, B., Waite, F., Harvey, A. G., & Harrison, P. J. (2020). Sleep disturbance and psychiatric disorders. Lancet Psychiatry, 7(7), 628–637. 10.1016/s2215-0366(20)30136-x

Gerosa, T., Fortunato, R., Cellina, F., Ragettli, M., Marzorati, D., Baldassari, A., Levati, S., Karabegovic, A., & Baumann, I. (2025). An App-Based Intervention to Protect Home Care Workers From Heat-Related Health Risks: Protocol for a Randomized Controlled Trial. *INQUIRY: The Journal of Health Care Organization*, Provision, and Financing, 62, 00469580251405766. 10.1177/00469580251405766

Herberger, S., Kräuchi, K., Glos, M., Lederer, K., Assmus, L., Hein, J., Penzel, T., & Fietze, I. (2019). Effects of sleep on a high-heat capacity mattress on sleep stages, EEG power spectra, cardiac interbeat intervals and body temperatures in healthy middle-aged men. Sleep, 43(5). 10.1093/sleep/zsz271

Hirshkowitz, M., Whiton, K., Albert, S. M., Alessi, C., Bruni, O., DonCarlos, L., Hazen, N., Herman, J., Adams Hillard, P. J., Katz, E. S., Kheirandish-Gozal, L., Neubauer, D. N., O’Donnell, A. E., Ohayon, M., Peever, J., Rawding, R., Sachdeva, R. C., Setters, B., Vitiello, M. V., & Ware, J. C. (2015). National Sleep Foundation’s updated sleep duration recommendations: final report. Sleep Health, 1(4), 233–243. 10.1016/j.sleh.2015.10.004

Ioannou, L. G., Tsoutsoubi, L., Mantzios, K., Ciuha, U., Kenny, G. P., Nybo, L., Flouris, A. D., & Mekjavic, I. B. (2024). Impact of a simulated multiday heatwave on nocturnal physiology, behavior, and sleep: a 10-day confinement study. Applied Physiology, Nutrition, and Metabolism, 49(10), 1394–1408. 10.1139/apnm-2024-0105

Kainec, K. A., Caccavaro, J., Barnes, M., Hoff, C., Berlin, A., & Spencer, R. M. C. (2024). Evaluating Accuracy in Five Commercial Sleep-Tracking Devices Compared to Research-Grade Actigraphy and Polysomnography. Sensors, 24(2), 635.

Kräuchi, K. (2002). How is the circadian rhythm of core body temperature regulated? Clinical Autonomic Research, 12(3), 147–149. 10.1007/s10286-002-0043-9

Kräuchi, K. (2007). The thermophysiological cascade leading to sleep initiation in relation to phase of entrainment. Sleep medicine reviews, 11(6), 439–451. 10.1016/j.smrv.2007.07.001

Kräuchi, K., Cajochen, C., Werth, E., & Wirz-Justice, A. (1999). Physiology - Warm feet promote the rapid onset of sleep. Nature, 401(6748), 36–37. 10.1038/43366

Kräuchi, K., Cajochen, C., Werth, E., & Wirz-Justice, A. (2000). Functional link between distal vasodilation and sleep-onset latency? *American Journal of Physiology - Regulatory*, Integrative and Comparative Physiology, 278(3), R741–R748. 10.1152/ajpregu.2000.278.3.r741

Kräuchi, K., Fattori, E., Giordano, A., Falbo, M., Iadarola, A., Aglì, F., Tribolo, A., Mutani, R., & Cicolin, A. (2018). Sleep on a high heat capacity mattress increases conductive body heat loss and slow wave sleep. Physiology & Behavior, 185, 23–30. 10.1016/j.physbeh.2017.12.014

Kräuchi, K., & Wirz-Justice, A. (2001). Circadian clues to sleep onset mechanisms. Neuropsychopharmacology, 25, S92–S96. 10.1016/S0893-133x(01)00315-3

Lan, L., Pan, L., Lian, Z., Huang, H., & Lin, Y. (2014). Experimental study on thermal comfort of sleeping people at different air temperatures. Building and Environment, 73, 24–31. 10.1016/j.buildenv.2013.11.024

Li, A., Luo, H., Zhu, Y., Zhang, Z., Liu, B., Kan, H., Jia, H., Wu, Z., Guo, Y., & Chen, R. (2025). Climate warming may undermine sleep duration and quality in repeated-measure study of 23 million records. Nature Communications, 16(1), 2609. 10.1038/s41467-025-57781-y

Libert, J. P., Di Nisi, J., Fukuda, H., Muzet, A., Ehrhart, J., & Amoros, C. (1988). Effect of Continuous Heat Exposure on Sleep Stages in Humans. Sleep, 11(2), 195–209. 10.1093/sleep/11.2.195

Liu, J., Kim, H., Hashizume, M., Lee, W., Honda, Y., Kim, S. E., He, C., Kan, H., & Chen, R. (2025). Nonlinear exposure-response associations of daytime, nighttime, and day-night compound heatwaves with mortality amid climate change. Nature Communications, 16(1), 635. 10.1038/s41467-025-56067-7

Liu, J., Varghese, B. M., Hansen, A., Xiang, J., Zhang, Y., Dear, K., Gourley, M., Driscoll, T., Morgan, G., Capon, A., & Bi, P. (2021). Is there an association between hot weather and poor mental health outcomes? A systematic review and meta-analysis. Environment International, 153, 106533. 10.1016/j.envint.2021.106533

Maloney, S. K., Kearney, M. R., & Mitchell, D. (2024). Indices of human heat stress in times of climate change. Acta Physiologica, 240(9), e14196. 10.1111/apha.14196

Mattingly, S. M., Grover, T., Martinez, G. J., Aledavood, T., Robles-Granda, P., Nies, K., Striegel, A., & Mark, G. (2021). The effects of seasons and weather on sleep patterns measured through longitudinal multimodal sensing. npj Digital Medicine, 4(1), 76. 10.1038/s41746-021-00435-2

McGregor, G. R., & Vanos, J. K. (2018). Heat: a primer for public health researchers. Public Health, 161, 138–146. 10.1016/j.puhe.2017.11.005

MeteoSwiss, & E.T.H. Zurich. (2025). Climate CH2025 - Scientific Report. 10.18751/climate/scenarios/ch2025/sr/1.0/

Minor, K., Bjerre-Nielsen, A., Jonasdottir, S. S., Lehmann, S., & Obradovich, N. (2022). Rising temperatures erode human sleep globally. One Earth, 5(5), 534–549. 10.1016/j.oneear.2022.04.008

Minor, K., Blume, C., Prather, A., Escobar Carias, M., Mullins, J. T., Aylward, B., Wortzel, J. R., Boateng, G. O., Weobong, B., Mindell, J. A., Crowley, S. J., Dugas, L. R., Harper, S., Clayton, S. D., Gringras, P., & Penzel, T. (2026). A wake-up call for a global climate and sleep task force. Sleep, 49(7), zsag075. 10.1093/sleep/zsag075

Murphy, P. J., & Campbell, S. S. (1997). Nighttime drop in body temperature: a physiological trigger for sleep onset? Sleep, 20(7), 505–511. 10.1093/sleep/20.7.505

Mutschler, R., Rüdisüli, M., Heer, P., & Eggimann, S. (2021). Benchmarking cooling and heating energy demands considering climate change, population growth and cooling device uptake. Applied Energy, 288, 116636. 10.1016/j.apenergy.2021.116636

Okamoto-Mizuno, K., Tsuzuki, K., Mizuno, K., & Iwaki, T. (2005). Effects of partial humid heat exposure during different segments of sleep on human sleep stages and body temperature. Physiology & Behavior, 83(5), 759–765. 10.1016/j.physbeh.2004.09.009

Ragettli, M. S., Flückiger, B., Vienneau, D., Domingo-Irigoyen, S., Koschenz, M., & Röösli, M. (2024). Vulnerability to heat-related mortality and the effect of prevention measures: a time-stratified case-crossover study in Switzerland. Swiss Medical Weekly, 154(10), 3410. 10.57187/s.3418

Rippstein, V., de Schrijver, E., Eckert, S., & Vicedo-Cabrera, A. M. (2023). Trends in tropical nights and their effects on mortality in Switzerland across 50 years. PLOS Climate, 2(4), e0000162. 10.1371/journal.pclm.0000162

Romanello, M., Walawender, M., Hsu, S.-C., Moskeland, A., Palmeiro-Silva, Y., Scamman, D., Smallcombe, J. W., Abdullah, S., Ades, M., Al-Maruf, A., Ameli, N., Angelova, D., Ayeb-Karlsson, S., Ballester, J., Basagaña, X., Bechara, H., Beggs, P. J., Cai, W., Campbell-Lendrum, D.,…Costello, A. (2025). The 2025 report of the <em>Lancet</em> Countdown on health and climate change: climate change action offers a lifeline. The Lancet, 406(10521), 2804–2857. 10.1016/S0140-6736(25)01919-1

Romanello, M., Walawender, M., Israelsson, J., Hasan, M. M., Battaglia, F., Dutta, A., Hsu, S.-C., Orgen, P., Palmeiro-Silva, Y., Scamman, D., Smallcombe, J. W., Abdullah, S., Ameli, N., Andrew, T. R., Angelova, D., Ayeb-Karlsson, S., Ballester, J., Basagaña, X., Basart, S.,…Costello, A. (2026). The 2026 report of the Lancet Countdown on health and climate change: a decade of escalating health harms and a decisive moment for all. The Lancet. 10.1016/S0140-6736(26)01779-4

Rumpf, H.-J., Meyer, C., Hapke, U., & John, U. (2001). Screening for mental health: validity of the MHI-5 using DSM-IV Axis I psychiatric disorders as gold standard. Psychiatry Research, 105(3), 243–253.

Sancho-Domingo, C., Carballo, J. L., Coloma-Carmona, A., & Buysse, D. J. (2021). Brief version of the Pittsburgh Sleep Quality Index (B-PSQI) and measurement invariance across gender and age in a population-based sample. Psychol Assess, 33(2), 111–121. 10.1037/pas0000959

Saucy, A., Ragettli, M. S., Vienneau, D., de Hoogh, K., Tangermann, L., Schäffer, B., Wunderli, J.-M., Probst-Hensch, N., & Röösli, M. (2021). The role of extreme temperature in cause-specific acute cardiovascular mortality in Switzerland: A case-crossover study. Science of The Total Environment, 790, 147958. 10.1016/j.scitotenv.2021.147958

Stull, R. (2011). Wet-Bulb Temperature from Relative Humidity and Air Temperature. Journal of Applied Meteorology and Climatology, 50(11), 2267–2269. 10.1175/JAMC-D-11-0143.1

Thompson, R., Hornigold, R., Page, L., & Waite, T. (2018). Associations between high ambient temperatures and heat waves with mental health outcomes: a systematic review. Public Health, 161, 171–191. 10.1016/j.puhe.2018.06.008

Thompson, R., Lawrance, E. L., Roberts, L. F., Grailey, K., Ashrafian, H., Maheswaran, H., Toledano, M. B., & Darzi, A. (2023). Ambient temperature and mental health: a systematic review and meta-analysis. Lancet Planet Health, 7(7), e580–e589. 10.1016/s2542-5196(23)00104-3

Troester, M. M., Quan, S. F., Berry, R. B., & American Academy of Sleep, M. (2023). The AASM Manual for the Scoring of Sleep and Associated Events: Rules, Terminology and Technical Specifications, Version 3. American Academy of Sleep Medicine. https://aasm.org/clinical-resources/scoring-manual/

Tsuzuki, K., Okamoto-Mizuno, K., & Mizuno, K. (2004). Effects of humid heat exposure on sleep, thermoregulation, melatonin, and microclimate. Journal of thermal biology, 29(1), 31–36. 10.1016/j.jtherbio.2003.10.003

Wright, A., Young, A., & Natarajan, S. (2005). Dwelling temperatures and comfort during the August 2003 heat wave. Building Services Engineering Research & Technology, 26(4), 285–300. 10.1191/0143624405bt136oa

