## Supplements for "Higher Nocturnal Temperatures Shorten and Worsen Sleep with No Clear Evidence of Short-Term Compensatory Patterns"

Supplementary Methods

**Suppl. Table S1. Overview of the REHEAT domains of intervention and corresponding adaptation measures.**

| **Intervention domains** | **Adaptation measures** |
| --- | --- |
| (1) Personal care | - Real-time self-monitoring of well-being - Info material: recognizing and addressing heat-related symptoms - Reminders to regularly check physiological data - Expert guidance on nutrition and hydration during hot weather (e.g., recipes, push notifications to remind participants to stay hydrated during heat episodes) - Tips for improving sleep quality during heat - Notifications to self-monitor sleep data |
| (2) Heat protection at work | - Recommendations for sunscreen, sunglasses, sun hats, lists of effective cooling devices according to personal needs - Handling work clothing (e.g. spare clothes) - Managing temperature changes, e.g. tips how to reduce heat exposure in vehicles and thus decrease the differences to temperature in air-conditioned spaces |
| (3) Work activity planning | - Weather alert notifications including standardized recommendations (e.g., “Tomorrow is expected to be hot, plan your breaks well and take enough fruit and water to drink”) |
| (4) Refreshment and  (5) leisure time | - Interactive maps of designated rest areas (e.g., bars, shops, libraries, parks), where workers can refresh, take breaks, use facilities - Alerts and map-based information on local leisure services (e.g., swimming pools, gyms) |

Supplementary Model Diagnostics

For models with divergent transitions, pair plots of selected posterior parameters were inspected with divergent transitions highlighted. These plots were used to assess whether divergences clustered in regions involving the main within-person wet-bulb terms, smooth-term variance parameters, random-effect standard deviations, or beta precision parameters.

**Supplementary Table S2. Convergence and sampling diagnostics for Bayesian same-night and exploratory lag models for nocturnal wet-bulb temperature.**

| **Outcome** | **Model label** | **Model ID** | **R̂ max** | **Bulk ESS min** | **Tail ESS min** | **Divergences** | **Max treedepth** |
| --- | --- | --- | --- | --- | --- | --- | --- |
| Sleep Maintenance Efficiency | Same-night smooth | lag0_smooth | 1.005 | 1588 | 2902 | 3 | 0 |
|  | Linear lags 0-1 | lag0_1_linear | 1.003 | 2194 | 3507 | 10 | 0 |
|  | Linear lags 0-2 | lag0_2_linear | 1.004 | 2002 | 3563 | 2 | 0 |
| Total Sleep Time | Same-night smooth | lag0_smooth | 1.004 | 1210 | 2370 | 0 | 0 |
|  | Linear lags 0-1 | lag0_1_linear | 1.004 | 1550 | 2851 | 1 | 0 |
|  | Linear lags 0-2 | lag0_2_linear | 1.003 | 1658 | 3188 | 10 | 0 |
| Light Sleep | Same-night smooth | lag0_smooth | 1.006 | 799 | 1677 | 7 | 0 |
|  | Linear lags 0-1 | lag0_1_linear | 1.004 | 915 | 2063 | 6 | 0 |
|  | Linear lags 0-2 | lag0_2_linear | 1.002 | 1020 | 2789 | 16 | 0 |
| Deep Sleep | Same-night smooth | lag0_smooth | 1.006 | 628 | 1245 | 3 | 0 |
|  | Linear lags 0-1 | lag0_1_linear | 1.008 | 836 | 1493 | 5 | 0 |
|  | Linear lags 0-2 | lag0_2_linear | 1.005 | 836 | 1629 | 7 | 0 |
| REM Sleep | Same-night smooth | lag0_smooth | 1.005 | 939 | 2437 | 11 | 0 |
|  | Linear lags 0-1 | lag0_1_linear | 1.004 | 1056 | 2065 | 0 | 0 |
|  | Linear lags 0-2 | lag0_2_linear | 1.003 | 1483 | 2789 | 0 | 0 |
| Recovery Index | Same-night smooth | lag0_smooth | 1.016 | 808 | 1716 | 0 | 0 |
|  | Linear lags 0-1 | lag0_1_linear | 1.004 | 1229 | 2555 | 0 | 0 |
|  | Linear lags 0-2 | lag0_2_linear | 1.003 | 1607 | 2856 | 0 | 0 |
| Subjective Sleep Quality | Same-night smooth | lag0_smooth | 1.003 | 1367 | 2389 | 2 | 0 |
|  | Linear lags 0-1 | lag0_1_linear | 1.005 | 1247 | 2610 | 0 | 0 |
|  | Linear lags 0-2 | lag0_2_linear | 1.004 | 1888 | 3405 | 3 | 0 |

For each outcome and model specification, the table reports the maximum R̂, minimum bulk and tail effective sample sizes, number of divergent transitions, and number of transitions reaching the maximum treedepth. Same-night smooth models correspond to the primary analyses; linear lag models correspond to exploratory carry-over analyses.

**Supplementary Table S3. Convergence and sampling diagnostics for Bayesian same-night and exploratory lag models for nocturnal air temperature.**

| **Outcome** | **Model label** | **Model ID** | **R̂ max** | **Bulk ESS min** | **Tail ESS min** | **Divergences** | **Max treedepth** |
| --- | --- | --- | --- | --- | --- | --- | --- |
| Sleep Maintenance Efficiency | Same-night smooth | lag0_smooth | 1.002 | 2157 | 4172 | 0 | 0 |
|  | Linear lags 0-1 | lag0_1_linear | 1.002 | 2000 | 3787 | 0 | 0 |
|  | Linear lags 0-2 | lag0_2_linear | 1.003 | 1785 | 3335 | 3 | 0 |
| Total Sleep Time | Same-night smooth | lag0_smooth | 1.003 | 1438 | 2988 | 0 | 0 |
|  | Linear lags 0-1 | lag0_1_linear | 1.002 | 1551 | 2946 | 1 | 0 |
|  | Linear lags 0-2 | lag0_2_linear | 1.004 | 1040 | 2019 | 2 | 0 |
| Light Sleep | Same-night smooth | lag0_smooth | 1.003 | 999 | 1874 | 16 | 0 |
|  | Linear lags 0-1 | lag0_1_linear | 1.007 | 798 | 1794 | 2 | 0 |
|  | Linear lags 0-2 | lag0_2_linear | 1.004 | 1040 | 2460 | 17 | 0 |
| Deep Sleep | Same-night smooth | lag0_smooth | 1.005 | 847 | 1864 | 13 | 0 |
|  | Linear lags 0-1 | lag0_1_linear | 1.006 | 711 | 1644 | 14 | 0 |
|  | Linear lags 0-2 | lag0_2_linear | 1.004 | 847 | 1741 | 16 | 0 |
| REM Sleep | Same-night smooth | lag0_smooth | 1.003 | 1233 | 2401 | 11 | 0 |
|  | Linear lags 0-1 | lag0_1_linear | 1.005 | 836 | 2479 | 0 | 0 |
|  | Linear lags 0-2 | lag0_2_linear | 1.003 | 1365 | 2473 | 0 | 0 |
| Recovery Index | Same-night smooth | lag0_smooth | 1.004 | 902 | 1645 | 0 | 0 |
|  | Linear lags 0-1 | lag0_1_linear | 1.006 | 1377 | 2070 | 0 | 0 |
|  | Linear lags 0-2 | lag0_2_linear | 1.005 | 1684 | 3155 | 1 | 0 |
| Subjective Sleep Quality | Same-night smooth | lag0_smooth | 1.006 | 1030 | 1933 | 3 | 0 |
|  | Linear lags 0-1 | lag0_1_linear | 1.004 | 1730 | 3347 | 0 | 0 |
|  | Linear lags 0-2 | lag0_2_linear | 1.005 | 1570 | 3124 | 1 | 0 |

For each outcome and model specification, the table reports the maximum R̂, minimum bulk and tail effective sample sizes, number of divergent transitions, and number of transitions reaching the maximum treedepth. Same-night smooth models correspond to the primary analyses; linear lag models correspond to exploratory carry-over analyses.

Supplementary Results
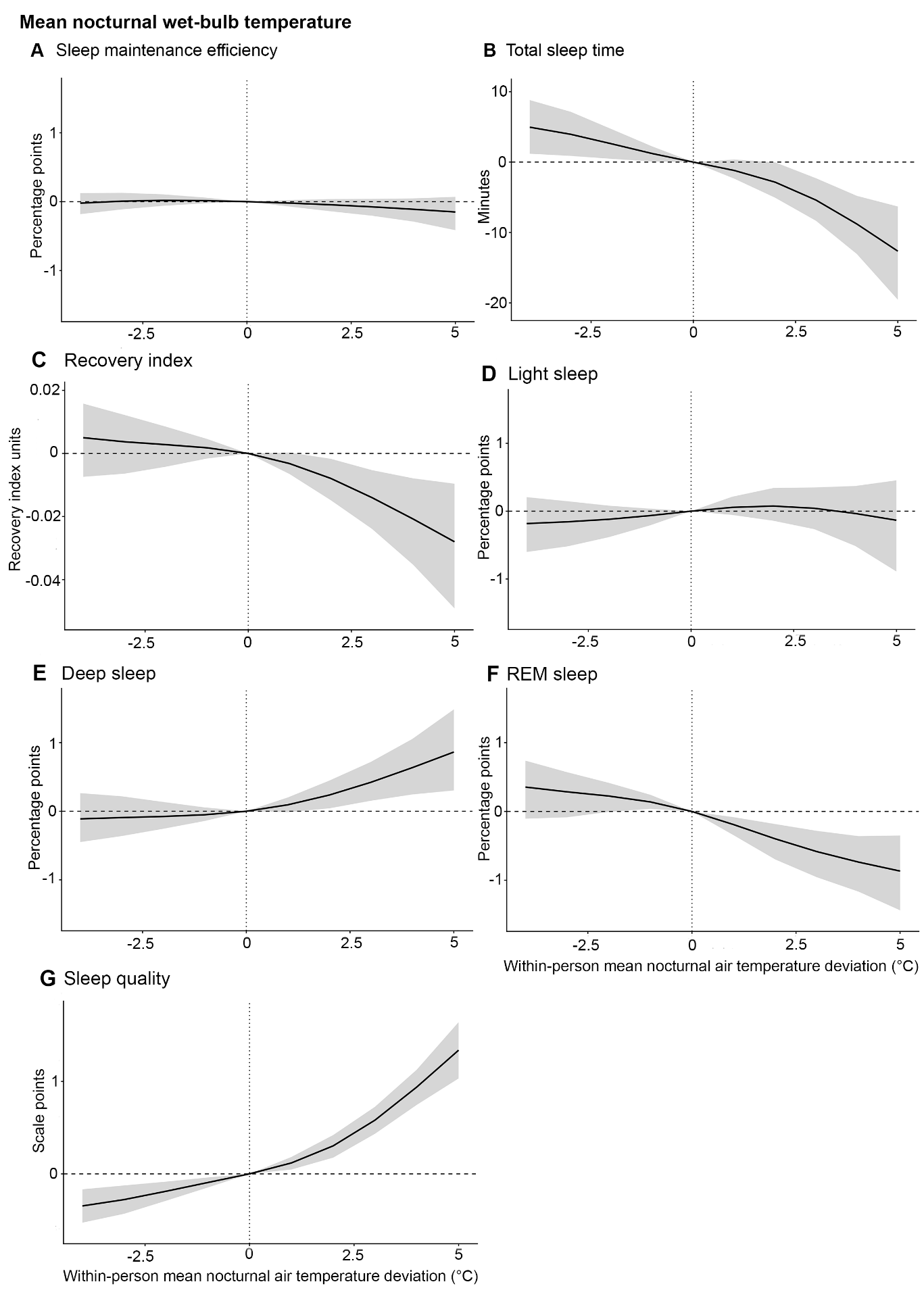

**Supplementary Figure S1. Primary same-night within-person associations between nocturnal wet-bulb temperature and sleep outcomes.** Posterior expected differences in (A) Sleep maintenance efficiency, (B) Total sleep time, (C) Recovery index, (D) Light sleep, (E) Deep sleep, (F) REM sleep, and (G) Subjective sleep quality corrected for baseline B-PSQI values are shown across within-person deviations in nocturnal wet-bulb temperature from a participant’s usual exposure. Black lines indicate posterior mean differences from the 0°C reference, and shaded areas indicate 95% credible intervals. The vertical dashed line marks a participant’s usual nocturnal wet-bulb exposure, and the horizontal dashed line marks no difference from this reference. Sleep maintenance efficiency and sleep-stage outcomes are expressed in percentage points; recovery index, total sleep time and subjective sleep quality are expressed in their native outcome units. Higher subjective sleep-quality scores indicate worse sleep quality. For the results table, please see Supplementary Table S4.

**Supplementary Table S4**. **Posterior expected sleep outcomes and differences from usual nocturnal wet-bulb exposure.**

| **Outcome** | **Unit** | **Wet-bulb deviation** | **Predicted outcome [95% CrI]** | **Difference from usual night, mean [95% CrI]** | **P(direction)** |
| --- | --- | --- | --- | --- | --- |
| Sleep maintenance efficiency | percentage points | -4°C | 96.15 [95.6, 96.65] | -0.02 [-0.18, 0.12] | 0.602 |
|  |  | -3°C | 96.18 [95.64, 96.67] | 0.01 [-0.11, 0.13] | 0.554 |
|  |  | -2°C | 96.19 [95.65, 96.67] | 0.02 [-0.06, 0.11] | 0.679 |
|  |  | -1°C | 96.18 [95.65, 96.67] | 0.01 [-0.02, 0.06] | 0.766 |
|  |  | 0°C reference | 96.17 [95.63, 96.66] |  |  |
|  |  | +1°C | 96.15 [95.61, 96.64] | -0.02 [-0.07, 0.02] | 0.831 |
|  |  | +2°C | 96.12 [95.58, 96.62] | -0.04 [-0.14, 0.04] | 0.864 |
|  |  | +3°C | 96.09 [95.55, 96.6] | -0.08 [-0.2, 0.04] | 0.897 |
|  |  | +4°C | 96.06 [95.49, 96.58] | -0.11 [-0.29, 0.05] | 0.909 |
|  |  | +5°C | 96.02 [95.42, 96.56] | -0.15 [-0.41, 0.07] | 0.9 |
| Total sleep time | minutes | -4°C | 410.58 [396.8, 424.52] | 4.96 [1.2, 8.81] | 0.994 |
|  |  | -3°C | 409.57 [395.97, 423.12] | 3.95 [0.89, 7.15] | 0.993 |
|  |  | -2°C | 408.22 [394.61, 421.61] | 2.6 [0.47, 4.68] | 0.99 |
|  |  | -1°C | 406.83 [393.32, 420.21] | 1.21 [0.09, 2.21] | 0.982 |
|  |  | 0°C reference | 405.62 [392.11, 418.95] |  |  |
|  |  | +1°C | 404.43 [390.99, 417.75] | -1.19 [-2.35, 0.37] | 0.936 |
|  |  | +2°C | 402.77 [389.18, 416.24] | -2.85 [-5.06, -0.04] | 0.976 |
|  |  | +3°C | 400.22 [386.58, 413.68] | -5.4 [-8.34, -2.3] | 0.999 |
|  |  | +4°C | 396.82 [382.96, 410.67] | -8.8 [-13.06, -4.8] | 1 |
|  |  | +5°C | 392.96 [377.99, 407.84] | -12.65 [-19.55, -6.31] | 1 |
| Light sleep | percentage points | -4°C | 63.51 [61.78, 65.21] | -0.18 [-0.6, 0.21] | 0.822 |
|  |  | -3°C | 63.54 [61.81, 65.22] | -0.16 [-0.52, 0.15] | 0.84 |
|  |  | -2°C | 63.57 [61.87, 65.24] | -0.12 [-0.38, 0.08] | 0.862 |
|  |  | -1°C | 63.63 [61.93, 65.28] | -0.07 [-0.21, 0.04] | 0.873 |
|  |  | 0°C reference | 63.69 [62, 65.33] |  |  |
|  |  | +1°C | 63.75 [62.06, 65.39] | 0.06 [-0.06, 0.21] | 0.82 |
|  |  | +2°C | 63.77 [62.07, 65.43] | 0.08 [-0.14, 0.34] | 0.75 |
|  |  | +3°C | 63.74 [62.03, 65.41] | 0.04 [-0.26, 0.35] | 0.621 |
|  |  | +4°C | 63.66 [61.93, 65.35] | -0.03 [-0.51, 0.37] | 0.535 |
|  |  | +5°C | 63.56 [61.75, 65.33] | -0.13 [-0.89, 0.45] | 0.612 |
| Deep sleep | percentage points | -4°C | 19.14 [17.72, 20.58] | -0.11 [-0.45, 0.26] | 0.733 |
|  |  | -3°C | 19.16 [17.76, 20.59] | -0.09 [-0.36, 0.22] | 0.748 |
|  |  | -2°C | 19.17 [17.77, 20.6] | -0.08 [-0.26, 0.13] | 0.786 |
|  |  | -1°C | 19.2 [17.79, 20.62] | -0.05 [-0.14, 0.06] | 0.85 |
|  |  | 0°C reference | 19.25 [17.84, 20.67] |  |  |
|  |  | +1°C | 19.34 [17.93, 20.78] | 0.09 [-0.01, 0.2] | 0.958 |
|  |  | +2°C | 19.48 [18.06, 20.94] | 0.24 [0.04, 0.45] | 0.99 |
|  |  | +3°C | 19.67 [18.22, 21.14] | 0.42 [0.15, 0.72] | 0.999 |
|  |  | +4°C | 19.88 [18.4, 21.41] | 0.64 [0.25, 1.05] | 1 |
|  |  | +5°C | 20.11 [18.57, 21.71] | 0.86 [0.3, 1.48] | 1 |
| REM sleep | percentage points | -4°C | 15.51 [14.04, 17.03] | 0.35 [-0.1, 0.74] | 0.937 |
|  |  | -3°C | 15.44 [14, 16.95] | 0.29 [-0.08, 0.57] | 0.942 |
|  |  | -2°C | 15.38 [13.97, 16.89] | 0.22 [-0.01, 0.42] | 0.973 |
|  |  | -1°C | 15.29 [13.89, 16.8] | 0.14 [0.04, 0.24] | 0.994 |
|  |  | 0°C reference | 15.15 [13.75, 16.65] |  |  |
|  |  | +1°C | 14.96 [13.57, 16.44] | -0.19 [-0.34, -0.08] | 1 |
|  |  | +2°C | 14.75 [13.37, 16.22] | -0.4 [-0.7, -0.18] | 1 |
|  |  | +3°C | 14.57 [13.18, 16.03] | -0.58 [-0.96, -0.28] | 1 |
|  |  | +4°C | 14.42 [13.03, 15.89] | -0.74 [-1.16, -0.36] | 1 |
|  |  | +5°C | 14.29 [12.87, 15.8] | -0.87 [-1.44, -0.35] | 0.999 |
| Recovery index | recovery-index units | -4°C | 0.51 [0.47, 0.56] | 0 [-0.01, 0.02] | 0.795 |
|  |  | -3°C | 0.51 [0.47, 0.56] | 0 [-0.01, 0.01] | 0.782 |
|  |  | -2°C | 0.51 [0.47, 0.55] | 0 [0, 0.01] | 0.805 |
|  |  | -1°C | 0.51 [0.46, 0.55] | 0 [0, 0] | 0.862 |
|  |  | 0°C reference | 0.51 [0.46, 0.55] |  |  |
|  |  | +1°C | 0.5 [0.46, 0.55] | 0 [-0.01, 0] | 0.966 |
|  |  | +2°C | 0.5 [0.46, 0.54] | -0.01 [-0.01, 0] | 0.994 |
|  |  | +3°C | 0.49 [0.45, 0.54] | -0.01 [-0.02, -0.01] | 1 |
|  |  | +4°C | 0.49 [0.44, 0.53] | -0.02 [-0.04, -0.01] | 1 |
|  |  | +5°C | 0.48 [0.43, 0.53] | -0.03 [-0.05, -0.01] | 1 |
| Subjective sleep quality, B-PSQI-adjusted | scale points | -4°C | 11.07 [10.48, 11.68] | -0.35 [-0.53, -0.17] | 1 |
|  |  | -3°C | 11.14 [10.56, 11.74] | -0.28 [-0.43, -0.13] | 1 |
|  |  | -2°C | 11.23 [10.64, 11.82] | -0.19 [-0.29, -0.09] | 1 |
|  |  | -1°C | 11.32 [10.74, 11.92] | -0.09 [-0.15, -0.04] | 1 |
|  |  | 0°C reference | 11.42 [10.83, 12.01] |  |  |
|  |  | +1°C | 11.54 [10.95, 12.14] | 0.12 [0.05, 0.18] | 0.999 |
|  |  | +2°C | 11.72 [11.13, 12.32] | 0.3 [0.18, 0.42] | 1 |
|  |  | +3°C | 12 [11.4, 12.6] | 0.58 [0.43, 0.73] | 1 |
|  |  | +4°C | 12.36 [11.75, 12.97] | 0.94 [0.75, 1.13] | 1 |
|  |  | +5°C | 12.76 [12.11, 13.42] | 1.34 [1.03, 1.64] | 1 |

Values are posterior means with 95% credible intervals. Wet-bulb deviation refers to the within-person deviation in same-night nocturnal wet-bulb temperature from a participant’s own usual nocturnal wet-bulb exposure. A deviation of 0°C therefore represents a usual night for that participant. Differences are expressed relative to this 0°C reference. For sleep maintenance efficiency and sleep-stage outcomes, differences are reported in percentage points; for recovery index and subjective sleep quality, differences are reported in native outcome units. Higher values of subjective sleep quality indicate worse sleep quality. P(direction) is the posterior probability that the expected difference from a participant’s usual night is in the estimated direction; for positive estimates, this is the probability that the difference is above zero, and for negative estimates, it is the probability that the difference is below zero. CrI, credible interval.

**Supplementary Table S5**. **Posterior expected sleep outcomes and differences from usual nocturnal air temperature exposure.**

| **Outcome** | **Unit** | **Wet-bulb deviation** | **Predicted outcome [95% CrI]** | **Difference from usual night, mean [95% CrI]** | **P(direction)** |
| --- | --- | --- | --- | --- | --- |
| Sleep maintenance efficiency | percentage points | -4°C | 96.19 [95.67, 96.69] | 0.03 [-0.11, 0.17] | 0.7 |
|  |  | -3°C | 96.2 [95.68, 96.68] | 0.04 [-0.07, 0.16] | 0.772 |
|  |  | -2°C | 96.19 [95.68, 96.67] | 0.03 [-0.04, 0.13] | 0.812 |
|  |  | -1°C | 96.18 [95.67, 96.66] | 0.02 [-0.02, 0.06] | 0.834 |
|  |  | 0°C reference | 96.16 [95.65, 96.64] |  |  |
|  |  | +1°C | 96.14 [95.63, 96.63] | -0.02 [-0.05, 0.02] | 0.837 |
|  |  | +2°C | 96.13 [95.61, 96.62] | -0.03 [-0.11, 0.05] | 0.83 |
|  |  | +3°C | 96.11 [95.58, 96.6] | -0.05 [-0.16, 0.06] | 0.844 |
|  |  | +4°C | 96.08 [95.55, 96.59] | -0.08 [-0.22, 0.06] | 0.878 |
|  |  | +5°C | 96.05 [95.49, 96.57] | -0.11 [-0.31, 0.06] | 0.9 |
| Total sleep time | minutes | -4°C | 409.86 [395.73, 423.91] | 3.67 [-0.42, 7.64] | 0.962 |
|  |  | -3°C | 408.51 [394.54, 422.46] | 2.32 [-1.42, 5.67] | 0.895 |
|  |  | -2°C | 407.58 [393.69, 421.41] | 1.39 [-1.52, 3.83] | 0.842 |
|  |  | -1°C | 406.94 [393.08, 420.74] | 0.75 [-0.67, 1.94] | 0.861 |
|  |  | 0°C reference | 406.19 [392.27, 419.95] |  |  |
|  |  | +1°C | 404.89 [391.07, 418.72] | -1.3 [-2.31, -0.13] | 0.985 |
|  |  | +2°C | 402.79 [388.82, 416.57] | -3.4 [-5.51, -1.2] | 0.998 |
|  |  | +3°C | 399.93 [385.83, 413.88] | -6.25 [-9.43, -3.25] | 1 |
|  |  | +4°C | 396.55 [382.4, 410.53] | -9.64 [-13.66, -5.83] | 1 |
|  |  | +5°C | 392.9 [378.32, 407.29] | -13.29 [-18.6, -8.03] | 1 |
| Light sleep | percentage points | -4°C | 63.37 [61.62, 65.1] | -0.26 [-0.7, 0.12] | 0.906 |
|  |  | -3°C | 63.46 [61.72, 65.16] | -0.17 [-0.51, 0.13] | 0.874 |
|  |  | -2°C | 63.54 [61.81, 65.23] | -0.1 [-0.33, 0.13] | 0.833 |
|  |  | -1°C | 63.59 [61.87, 65.28] | -0.04 [-0.15, 0.07] | 0.802 |
|  |  | 0°C reference | 63.63 [61.92, 65.33] |  |  |
|  |  | +1°C | 63.67 [61.94, 65.36] | 0.03 [-0.07, 0.14] | 0.746 |
|  |  | +2°C | 63.69 [61.96, 65.37] | 0.05 [-0.17, 0.26] | 0.707 |
|  |  | +3°C | 63.69 [61.94, 65.38] | 0.06 [-0.27, 0.35] | 0.664 |
|  |  | +4°C | 63.68 [61.92, 65.39] | 0.04 [-0.37, 0.41] | 0.609 |
|  |  | +5°C | 63.65 [61.86, 65.38] | 0.02 [-0.53, 0.49] | 0.558 |
| Deep sleep | percentage points | -4°C | 19.09 [17.64, 20.58] | -0.14 [-0.49, 0.22] | 0.782 |
|  |  | -3°C | 19.06 [17.63, 20.53] | -0.17 [-0.46, 0.12] | 0.887 |
|  |  | -2°C | 19.08 [17.65, 20.54] | -0.16 [-0.38, 0.05] | 0.939 |
|  |  | -1°C | 19.14 [17.72, 20.59] | -0.1 [-0.21, 0] | 0.973 |
|  |  | 0°C reference | 19.24 [17.81, 20.69] |  |  |
|  |  | +1°C | 19.36 [17.93, 20.82] | 0.12 [0.03, 0.22] | 0.996 |
|  |  | +2°C | 19.5 [18.07, 20.99] | 0.26 [0.08, 0.46] | 0.996 |
|  |  | +3°C | 19.67 [18.22, 21.17] | 0.43 [0.16, 0.72] | 0.998 |
|  |  | +4°C | 19.87 [18.39, 21.37] | 0.63 [0.3, 0.98] | 1 |
|  |  | +5°C | 20.09 [18.58, 21.64] | 0.86 [0.42, 1.32] | 1 |
| REM sleep | percentage points | -4°C | 15.71 [14.21, 17.29] | 0.51 [0.11, 0.88] | 0.992 |
|  |  | -3°C | 15.62 [14.15, 17.18] | 0.43 [0.12, 0.71] | 0.994 |
|  |  | -2°C | 15.51 [14.06, 17.06] | 0.32 [0.11, 0.53] | 0.996 |
|  |  | -1°C | 15.37 [13.93, 16.9] | 0.17 [0.07, 0.28] | 0.999 |
|  |  | 0°C reference | 15.19 [13.78, 16.71] |  |  |
|  |  | +1°C | 15 [13.59, 16.5] | -0.19 [-0.3, -0.1] | 1 |
|  |  | +2°C | 14.8 [13.4, 16.28] | -0.39 [-0.62, -0.21] | 1 |
|  |  | +3°C | 14.6 [13.2, 16.07] | -0.59 [-0.91, -0.33] | 1 |
|  |  | +4°C | 14.41 [13.03, 15.89] | -0.79 [-1.17, -0.46] | 1 |
|  |  | +5°C | 14.22 [12.82, 15.71] | -0.98 [-1.45, -0.55] | 1 |
| Recovery index | recovery-index units | -4°C | 0.52 [0.48, 0.56] | 0.01 [0, 0.02] | 0.956 |
|  |  | -3°C | 0.52 [0.47, 0.56] | 0.01 [0, 0.02] | 0.951 |
|  |  | -2°C | 0.52 [0.47, 0.56] | 0.01 [0, 0.01] | 0.964 |
|  |  | -1°C | 0.51 [0.47, 0.56] | 0 [0, 0.01] | 0.988 |
|  |  | 0°C reference | 0.51 [0.47, 0.55] |  |  |
|  |  | +1°C | 0.5 [0.46, 0.55] | 0 [-0.01, 0] | 1 |
|  |  | +2°C | 0.5 [0.46, 0.54] | -0.01 [-0.02, 0] | 1 |
|  |  | +3°C | 0.49 [0.45, 0.54] | -0.02 [-0.03, -0.01] | 1 |
|  |  | +4°C | 0.49 [0.44, 0.53] | -0.02 [-0.03, -0.01] | 1 |
|  |  | +5°C | 0.48 [0.44, 0.53] | -0.02 [-0.04, -0.01] | 0.999 |
| Subjective sleep quality, PSQI-adjusted | scale points | -4°C | 11.11 [10.49, 11.72] | -0.25 [-0.44, -0.06] | 0.997 |
|  |  | -3°C | 11.18 [10.57, 11.79] | -0.18 [-0.34, -0.01] | 0.979 |
|  |  | -2°C | 11.24 [10.63, 11.85] | -0.12 [-0.24, 0.01] | 0.967 |
|  |  | -1°C | 11.29 [10.68, 11.89] | -0.07 [-0.13, -0.01] | 0.987 |
|  |  | 0°C reference | 11.36 [10.75, 11.97] |  |  |
|  |  | +1°C | 11.48 [10.87, 12.09] | 0.12 [0.06, 0.17] | 1 |
|  |  | +2°C | 11.66 [11.05, 12.27] | 0.3 [0.19, 0.41] | 1 |
|  |  | +3°C | 11.91 [11.3, 12.53] | 0.55 [0.4, 0.71] | 1 |
|  |  | +4°C | 12.21 [11.59, 12.83] | 0.85 [0.67, 1.04] | 1 |
|  |  | +5°C | 12.54 [11.9, 13.18] | 1.18 [0.95, 1.41] | 1 |

Values are posterior means with 95% credible intervals. Air temperature deviation refers to the within-person deviation in same-night nocturnal air temperature from a participant’s own usual nocturnal air temperature exposure. A deviation of 0°C therefore represents a usual night for that participant. Differences are expressed relative to this 0°C reference. For sleep maintenance efficiency and sleep-stage outcomes, differences are reported in percentage points; for total sleep time, recovery index and subjective sleep quality, differences are reported in native outcome units. Higher values of subjective sleep quality indicate worse sleep quality. P(direction) is the posterior probability that the expected difference from a participant’s usual night is in the estimated direction; for positive estimates, this is the probability that the difference is above zero, and for negative estimates, it is the probability that the difference is below zero. CrI, credible interval.

Absolute mean nocturnal air temperature centred at 17°C

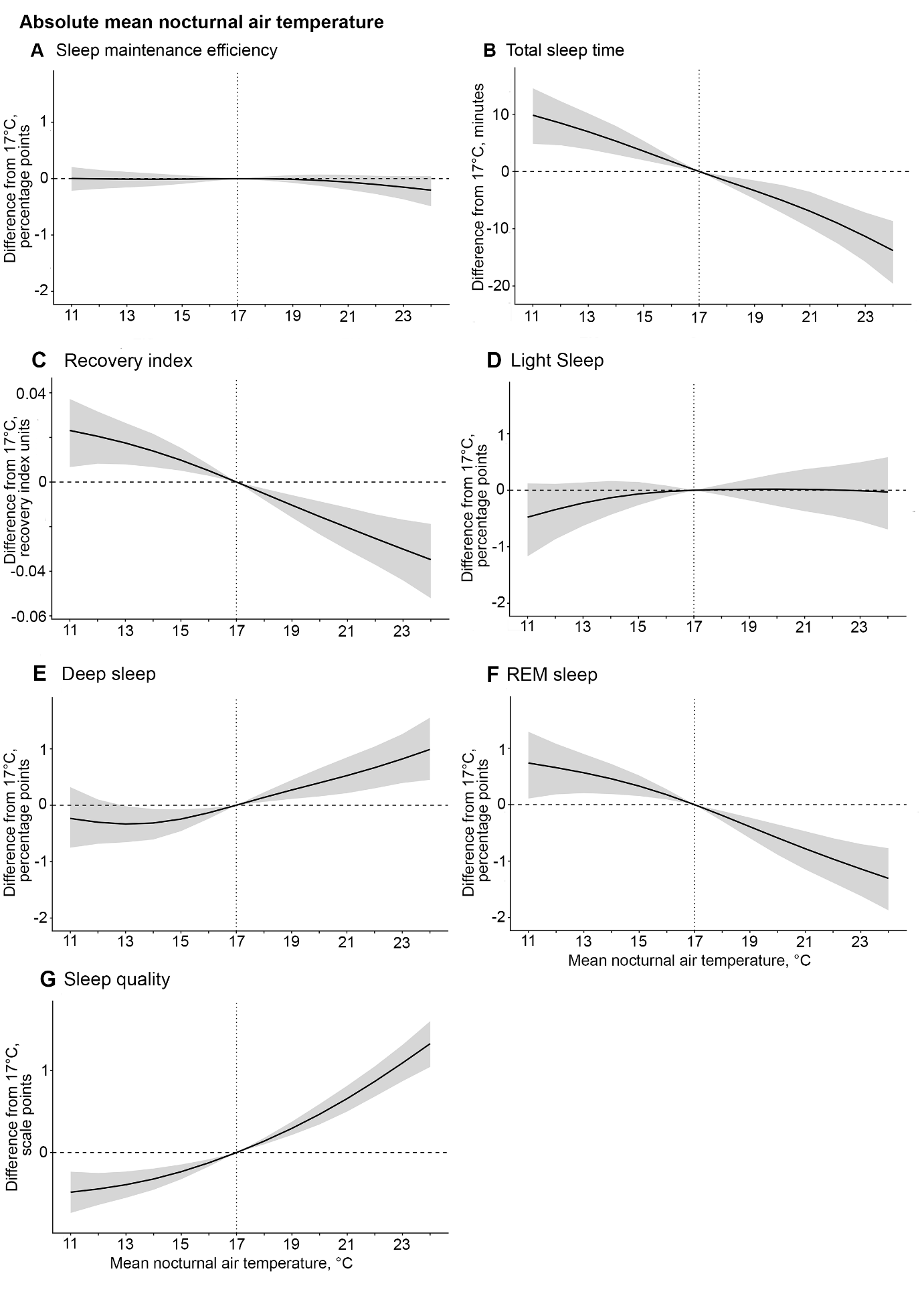

**Supplementary Figure S2. Same-night within-person associations between absolute mean nocturnal air temperature centred at 17 °C and sleep outcomes.** Posterior expected differences in (A) Sleep maintenance efficiency, (B) Total sleep time, (C) Recovery index, (D) Light sleep, (E) Deep sleep, (F) REM sleep, and (G) Subjective sleep quality corrected for baseline B-PSQI values are shown across absolute deviations in nocturnal air temperature from 17 °C. Black lines indicate posterior mean differences from the 17 °C reference, and shaded areas indicate 95% credible intervals. The vertical dashed line marks 17 °C, and the horizontal dashed line marks no difference from this reference. Sleep maintenance efficiency and sleep-stage outcomes are expressed in percentage points; recovery index, total sleep time, and subjective sleep quality are expressed in their native outcome units. Higher subjective sleep-quality scores indicate worse sleep quality. For the results table, please see Supplementary Table S6.

**Supplementary Table S6.** **Posterior expected sleep outcomes at selected mean nocturnal air temperatures and differences relative to 17°C.**

| **Outcome** | **Unit** | **Mean nocturnal air temp.** | **Predicted outcome, mean [95% CrI]** | **Difference from 17°C, mean [95% CrI]** | **P(direction)** |
| --- | --- | --- | --- | --- | --- |
| Sleep maintenance efficiency | percentage points | 11°C | 96.22 [95.64, 96.73] | 0 [-0.21, 0.21] | 0.53 |
|  |  | 12°C | 96.21 [95.65, 96.71] | 0 [-0.18, 0.15] | 0.50 |
|  |  | 13°C | 96.2 [95.66, 96.7] | -0.01 [-0.16, 0.12] | 0.53 |
|  |  | 14°C | 96.2 [95.66, 96.69] | -0.01 [-0.13, 0.09] | 0.55 |
|  |  | 15°C | 96.2 [95.67, 96.69] | -0.01 [-0.09, 0.06] | 0.55 |
|  |  | 16°C | 96.21 [95.68, 96.7] | 0 [-0.05, 0.03] | 0.54 |
|  |  | 17°C, reference | 96.21 [95.68, 96.7] |  |  |
|  |  | 18°C | 96.21 [95.68, 96.7] | 0 [-0.03, 0.04] | 0.56 |
|  |  | 19°C | 96.2 [95.67, 96.69] | -0.01 [-0.07, 0.06] | 0.65 |
|  |  | 20°C | 96.18 [95.65, 96.67] | -0.03 [-0.13, 0.07] | 0.75 |
|  |  | 21°C | 96.15 [95.61, 96.64] | -0.06 [-0.2, 0.06] | 0.84 |
|  |  | 22°C | 96.11 [95.56, 96.61] | -0.1 [-0.27, 0.05] | 0.91 |
|  |  | 23°C | 96.06 [95.49, 96.57] | -0.15 [-0.37, 0.04] | 0.94 |
|  |  | 24°C | 96.01 [95.4, 96.55] | -0.2 [-0.49, 0.04] | 0.94 |
| Total sleep time | minutes | 11°C | 414.81 [400.57, 428.8] | 9.84 [4.83, 14.53] | 1 |
|  |  | 12°C | 413.43 [399.54, 427.18] | 8.45 [4.58, 12.28] | 1 |
|  |  | 13°C | 411.93 [398.16, 425.6] | 6.95 [3.88, 10.15] | 1 |
|  |  | 14°C | 410.28 [396.51, 423.91] | 5.3 [2.95, 7.94] | 1 |
|  |  | 15°C | 408.53 [394.95, 422.04] | 3.55 [1.96, 5.35] | 1 |
|  |  | 16°C | 406.73 [393.15, 420.23] | 1.75 [0.97, 2.58] | 1 |
|  |  | 17°C, reference | 404.98 [391.34, 418.49] |  |  |
|  |  | 18°C | 403.29 [389.61, 416.78] | -1.68 [-2.41, -0.86] | 1 |
|  |  | 19°C | 401.64 [387.96, 415.1] | -3.34 [-4.81, -1.56] | 1 |
|  |  | 20°C | 399.92 [386.24, 413.36] | -5.06 [-7.27, -2.37] | 1 |
|  |  | 21°C | 398.04 [384.36, 411.6] | -6.94 [-9.85, -3.56] | 1 |
|  |  | 22°C | 395.94 [382.14, 409.69] | -9.03 [-12.58, -5.4] | 1 |
|  |  | 23°C | 393.63 [379.6, 407.57] | -11.35 [-15.78, -7.18] | 1 |
|  |  | 24°C | 391.16 [376.61, 405.51] | -13.81 [-19.62, -8.67] | 1 |
| Light sleep | percentage points | 11°C | 63.11 [61.28, 65.03] | -0.48 [-1.17, 0.12] | 0.94 |
|  |  | 12°C | 63.25 [61.47, 65.1] | -0.34 [-0.87, 0.11] | 0.92 |
|  |  | 13°C | 63.36 [61.6, 65.2] | -0.23 [-0.63, 0.14] | 0.89 |
|  |  | 14°C | 63.46 [61.72, 65.28] | -0.13 [-0.43, 0.16] | 0.82 |
|  |  | 15°C | 63.52 [61.77, 65.33] | -0.07 [-0.26, 0.15] | 0.76 |
|  |  | 16°C | 63.57 [61.82, 65.38] | -0.02 [-0.12, 0.08] | 0.70 |
|  |  | 17°C, reference | 63.59 [61.85, 65.4] |  |  |
|  |  | 18°C | 63.6 [61.87, 65.42] | 0.01 [-0.09, 0.1] | 0.62 |
|  |  | 19°C | 63.61 [61.88, 65.43] | 0.02 [-0.18, 0.2] | 0.60 |
|  |  | 20°C | 63.61 [61.88, 65.42] | 0.02 [-0.28, 0.29] | 0.57 |
|  |  | 21°C | 63.6 [61.87, 65.43] | 0.02 [-0.37, 0.37] | 0.55 |
|  |  | 22°C | 63.6 [61.86, 65.42] | 0.01 [-0.45, 0.43] | 0.52 |
|  |  | 23°C | 63.58 [61.82, 65.41] | -0.01 [-0.55, 0.5] | 0.50 |
|  |  | 24°C | 63.56 [61.75, 65.43] | -0.03 [-0.69, 0.58] | 0.52 |
| Deep sleep | percentage points | 11°C | 19 [17.57, 20.48] | -0.23 [-0.75, 0.32] | 0.80 |
|  |  | 12°C | 18.94 [17.55, 20.35] | -0.3 [-0.68, 0.1] | 0.93 |
|  |  | 13°C | 18.91 [17.53, 20.3] | -0.33 [-0.66, -0.02] | 0.98 |
|  |  | 14°C | 18.92 [17.55, 20.33] | -0.32 [-0.61, -0.07] | 0.99 |
|  |  | 15°C | 18.99 [17.63, 20.4] | -0.25 [-0.46, -0.07] | 1.00 |
|  |  | 16°C | 19.1 [17.76, 20.5] | -0.13 [-0.24, -0.05] | 1.00 |
|  |  | 17°C, reference | 19.24 [17.89, 20.64] |  |  |
|  |  | 18°C | 19.38 [18.01, 20.79] | 0.14 [0.06, 0.23] | 1.00 |
|  |  | 19°C | 19.51 [18.13, 20.93] | 0.27 [0.11, 0.45] | 1.00 |
|  |  | 20°C | 19.63 [18.25, 21.07] | 0.4 [0.16, 0.65] | 1.00 |
|  |  | 21°C | 19.76 [18.36, 21.22] | 0.53 [0.21, 0.85] | 1.00 |
|  |  | 22°C | 19.9 [18.48, 21.37] | 0.67 [0.3, 1.04] | 1.00 |
|  |  | 23°C | 20.06 [18.61, 21.54] | 0.82 [0.39, 1.26] | 1.00 |
|  |  | 24°C | 20.23 [18.74, 21.77] | 0.99 [0.45, 1.55] | 1.00 |
| REM sleep | percentage points | 11°C | 16.09 [14.55, 17.74] | 0.74 [0.11, 1.29] | 0.99 |
|  |  | 12°C | 16.01 [14.5, 17.61] | 0.66 [0.18, 1.08] | 1 |
|  |  | 13°C | 15.91 [14.44, 17.5] | 0.57 [0.21, 0.9] | 1 |
|  |  | 14°C | 15.81 [14.35, 17.38] | 0.46 [0.19, 0.72] | 1 |
|  |  | 15°C | 15.68 [14.24, 17.24] | 0.33 [0.15, 0.52] | 1 |
|  |  | 16°C | 15.52 [14.1, 17.07] | 0.18 [0.09, 0.27] | 1 |
|  |  | 17°C, reference | 15.35 [13.93, 16.87] |  |  |
|  |  | 18°C | 15.16 [13.76, 16.67] | -0.19 [-0.29, -0.11] | 1 |
|  |  | 19°C | 14.96 [13.57, 16.44] | -0.39 [-0.59, -0.23] | 1 |
|  |  | 20°C | 14.76 [13.38, 16.24] | -0.59 [-0.89, -0.35] | 1 |
|  |  | 21°C | 14.57 [13.2, 16.04] | -0.78 [-1.15, -0.47] | 1 |
|  |  | 22°C | 14.39 [13.03, 15.84] | -0.96 [-1.38, -0.59] | 1 |
|  |  | 23°C | 14.21 [12.85, 15.66] | -1.14 [-1.61, -0.7] | 1 |
|  |  | 24°C | 14.04 [12.67, 15.52] | -1.31 [-1.87, -0.77] | 1 |
| Recovery index | recovery-index units | 11°C | 0.52 [0.48, 0.57] | 0.02 [0.01, 0.04] | 1 |
|  |  | 12°C | 0.52 [0.47, 0.57] | 0.02 [0.01, 0.03] | 1 |
|  |  | 13°C | 0.52 [0.47, 0.57] | 0.02 [0.01, 0.03] | 1 |
|  |  | 14°C | 0.52 [0.47, 0.56] | 0.01 [0.01, 0.02] | 1 |
|  |  | 15°C | 0.51 [0.46, 0.56] | 0.01 [0.01, 0.02] | 1 |
|  |  | 16°C | 0.51 [0.46, 0.55] | 0.01 [0, 0.01] | 1 |
|  |  | 17°C, reference | 0.5 [0.45, 0.55] |  |  |
|  |  | 18°C | 0.5 [0.45, 0.54] | -0.01 [-0.01, 0] | 1 |
|  |  | 19°C | 0.49 [0.44, 0.54] | -0.01 [-0.02, -0.01] | 1 |
|  |  | 20°C | 0.49 [0.44, 0.53] | -0.02 [-0.02, -0.01] | 1 |
|  |  | 21°C | 0.48 [0.43, 0.53] | -0.02 [-0.03, -0.01] | 1 |
|  |  | 22°C | 0.48 [0.43, 0.52] | -0.03 [-0.04, -0.01] | 1 |
|  |  | 23°C | 0.47 [0.42, 0.52] | -0.03 [-0.04, -0.02] | 1 |
|  |  | 24°C | 0.47 [0.42, 0.51] | -0.03 [-0.05, -0.02] | 1 |
| Subjective sleep quality, BPSQI-adjusted | scale points | 11°C | 10.8 [10.18, 11.45] | -0.48 [-0.74, -0.23] | 1 |
|  |  | 12°C | 10.84 [10.24, 11.47] | -0.44 [-0.64, -0.25] | 1 |
|  |  | 13°C | 10.9 [10.29, 11.52] | -0.39 [-0.55, -0.23] | 1 |
|  |  | 14°C | 10.97 [10.37, 11.58] | -0.32 [-0.45, -0.2] | 1 |
|  |  | 15°C | 11.05 [10.46, 11.66] | -0.23 [-0.32, -0.15] | 1 |
|  |  | 16°C | 11.16 [10.57, 11.78] | -0.12 [-0.17, -0.08] | 1 |
|  |  | 17°C, reference | 11.29 [10.7, 11.9] |  |  |
|  |  | 18°C | 11.43 [10.84, 12.04] | 0.14 [0.1, 0.18] | 1 |
|  |  | 19°C | 11.58 [10.99, 12.19] | 0.29 [0.21, 0.38] | 1 |
|  |  | 20°C | 11.75 [11.16, 12.37] | 0.47 [0.34, 0.59] | 1 |
|  |  | 21°C | 11.95 [11.34, 12.56] | 0.66 [0.5, 0.81] | 1 |
|  |  | 22°C | 12.16 [11.55, 12.77] | 0.87 [0.68, 1.05] | 1 |
|  |  | 23°C | 12.38 [11.76, 13.01] | 1.09 [0.87, 1.31] | 1 |
|  |  | 24°C | 12.62 [11.98, 13.27] | 1.33 [1.04, 1.6] | 1 |

Predicted outcomes are posterior means with 95% credible intervals from the primary same-night models. Differences were calculated relative to the posterior expected outcome at a mean nocturnal air temperature of 17°C, with negative values indicating lower and positive values indicating higher expected outcomes. Sleep maintenance efficiency and sleep-stage percentages are presented on the response scale as percentages; total sleep time, recovery index, and subjective sleep quality are presented in minutes, recovery-index units, and scale points, respectively. Higher subjective sleep-quality scores indicate poorer sleep quality. P(direction) represents the posterior probability that the difference from 17°C is in the estimated direction. CrI, credible interval.

Heat Episodes

**Supplementary Table S7. Descriptives for exposure during heat-episode nights.**

|  | min | max | mean | median | sd |
| --- | --- | --- | --- | --- | --- |
| Nocturnal air temperature (in °C) | 15.6 | 25.8 | 21.0 | 20.8 | 2.1 |
| Minimum air temperature (in °C) | 13.9 | 24.6 | 19.3 | 19.1 | 2.3 |
| Nocturnal wet-bulb temperature (in °C) | 13.1 | 22.1 | 18.4 | 18.0 | 1.9 |

The minimum number of heat-episode nights was 1, the maximum 28. On average, participants experienced 9.4 heat-episode nights. Suppl. Figure S3 provides a histogram of the total number of available heat-episode nights.

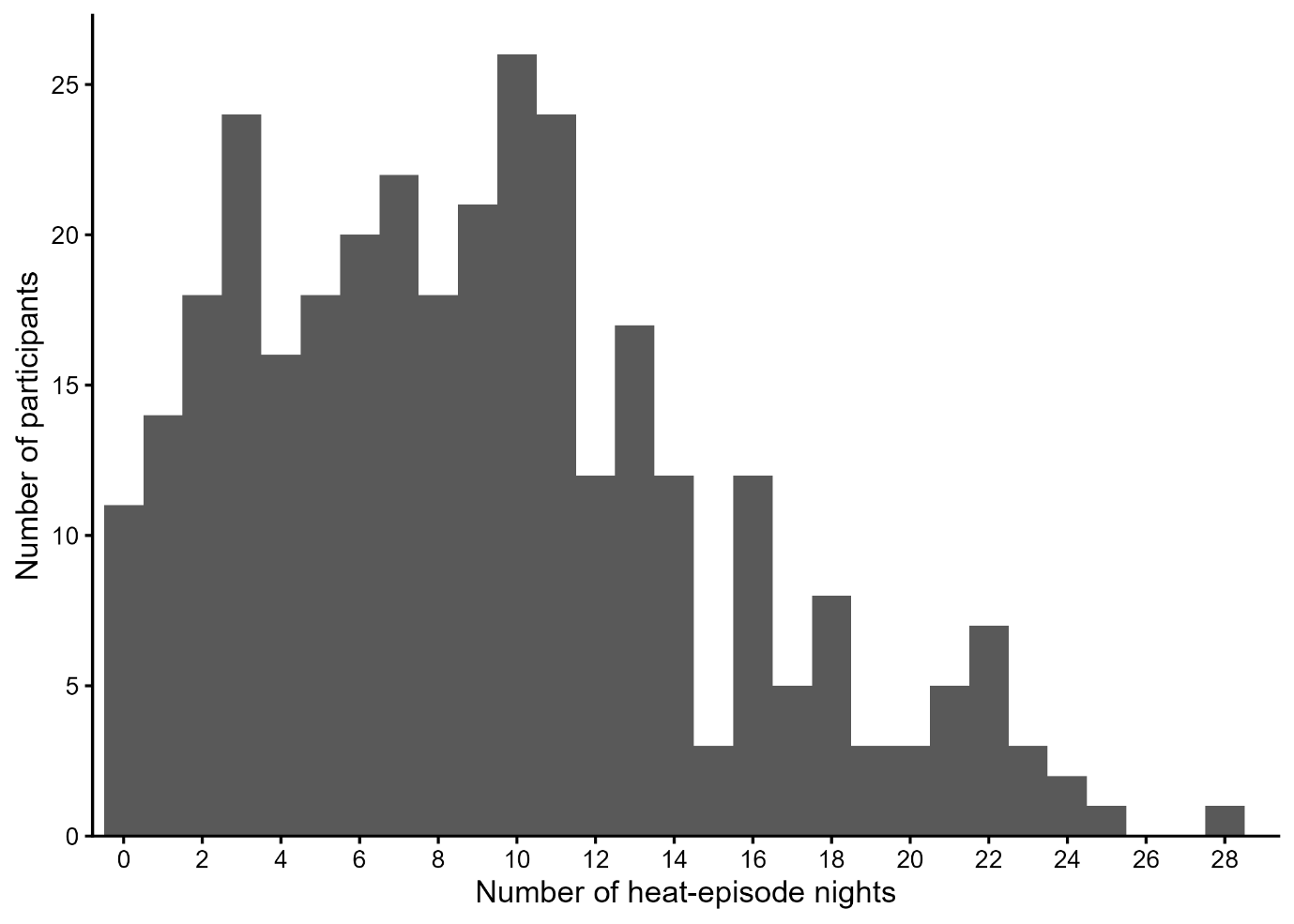

**Supplementary Figure S3. Histogram for the number of heat-episode nights experienced by participants.**

**Supplementary Table S8.** **Associations between nocturnal temperature and sleep outcomes during heat episodes.**

**Panel A. Mean nocturnal air temperature**

| **Outcome** | **Scale** | **Within-person coefficient, mean [95% CrI]; P(direction)** | **Between-person coefficient, mean [95% CrI]; P(direction)** |
| --- | --- | --- | --- |
| Sleep maintenance efficiency (SME) | Beta-regression logit scale | -0.02 [-0.04, 0.01]; 0.93 | 0.02 [-0.02, 0.05]; 0.79 |
| Light sleep | Beta-regression logit scale | 0.0 [-0.01, 0.01]; 0.58 | -0.01 [-0.03, 0.01]; 0.81 |
| Deep sleep | Beta-regression logit scale | 0.01 [-0.01, 0.02]; 0.83 | 0.01 [-0.01, 0.03]; 0.83 |
| REM sleep | Beta-regression logit scale | -0.01 [-0.02, 0.01]; 0.79 | -0.01 [-0.03, 0.02]; 0.70 |
| Recovery index | Recovery-index units | -0.01 [-0.01, 0.0]; 0.95 | -0.03 [-0.04, -0.02]; 1 |
| Total sleep time (TST) | Minutes | -2.06 [-4.05, -0.08]; 0.98 | -3.88 [-7.11, -0.71]; 0.90 |
| Subjective sleep quality, BPSQI-adjusted | Scale points | 0.27 [0.18, 0.36]; 1 | 0.07 [-0.09, 0.23]; 0.81 |

**Banel B. Mean nocturnal wet-bulb temperature**

| **Outcome** | **Scale** | **Within-person coefficient, mean [95% CrI]; P(direction)** | **Between-person coefficient, mean [95% CrI]; P(direction)** |
| --- | --- | --- | --- |
| Sleep maintenance efficiency (SME) | Beta-regression logit scale | -0.01 [-0.04, 0.01]; 0.91 | 0.02 [-0.03, 0.07]; 0.73 |
| Light sleep | Beta-regression logit scale | 0.0 [-0.01, 0.01]; 0.61 | -0.01 [-0.04, 0.01]; 0.88 |
| Deep sleep | Beta-regression logit scale | 0.0 [-0.01, 0.01]; 0.58 | 0.02 [-0.01, 0.05]; 0.90 |
| REM sleep | Beta-regression logit scale | 0.0 [-0.02, 0.01]; 0.66 | -0.01 [-0.05, 0.02]; 0.79 |
| Recovery index | Recovery-index units | -0.01 [-0.01, 0.0]; 0.98 | -0.03 [-0.04, -0.02]; 1 |
| Total sleep time (TST) | Minutes | -1.19 [-2.99, 0.55]; 0.91 | -5.65 [-9.85, -1.56]; 1 |
| Subjective sleep quality, BPSQI-adjusted | Scale points | 0.21 [0.14, 0.29]; 1 | 0.06 [-0.14, 0.26]; 0.71 |

Values are posterior mean coefficients [95% credible intervals] followed by P(direction). Within-person coefficients represent the estimated difference in the sleep outcome associated with a 1°C higher nocturnal temperature relative to the participant’s mean exposure during the heat episode. Between-person coefficients represent the estimated difference associated with a 1°C higher participant-level mean nocturnal temperature. Sleep maintenance efficiency and sleep-stage percentages are reported on the beta-regression logit scale; total sleep time, recovery index, and subjective sleep quality are reported in minutes, recovery-index units, and scale points, respectively. Higher subjective sleep-quality scores indicate poorer sleep quality. Models of subjective sleep quality were additionally adjusted for baseline BPSQI. CrI, credible interval; BPSQI, Brief Pittsburgh Sleep Quality Index; P(direction), posterior probability that the coefficient is in the estimated direction.

Probability of short sleep

**Supplementary Table S9. Posterior probabilities of short sleep in relation to mean nocturnal air temperature.**

**Panel A. Within-person deviations in mean nocturnal air temperature**

| **Air temp. deviation** | **P(TST<7h)  [95% CrI]** | **Difference from usual [95% CrI], pp** | **P(dir)** | **P(TST<6h)  [95% CrI]** | **Difference from usual [95% CrI], pp** | **P(dir)** |
| --- | --- | --- | --- | --- | --- | --- |
| -4°C | 55.7 [46.4, 64.8] | -3.9 [-6.3, -1.0] | 1 | 23.6 [16.9, 31.3] | -0.2 [-2.8, 2.8] | 0.56 |
| -3°C | 56.7 [47.6, 65.5] | -2.9 [-4.7, -0.5] | 1 | 23.4 [16.9, 31] | -0.4 [-2.3, 1.9] | 0.65 |
| -2°C | 57.6 [48.7, 66.4] | -1.9 [-3.2, -0.3] | 0.99 | 23.3 [16.9, 30.7] | -0.4 [-1.8, 1.1] | 0.75 |
| -1°C | 58.5 [49.7, 67.2] | -1 [-1.6, -0.2] | 0.99 | 23.4 [17, 30.8] | -0.3 [-1, 0.3] | 0.86 |
| 0°C ref | 59.6 [50.7, 68.1] |  |  | 23.8 [17.3, 31.2] |  |  |
| +1°C | 60.7 [51.9, 69.2] | 1.1 [0.5, 1.7] | 1 | 24.4 [17.8, 31.9] | 0.6 [0, 1.3] | 0.98 |
| +2°C | 62 [53.3, 70.3] | 2.4 [1.3, 3.7] | 1 | 25.3 [18.5, 32.9] | 1.5 [0.3, 3] | 0.99 |
| +3°C | 63.4 [54.6, 71.5] | 3.8 [2.2, 5.8] | 1 | 26.3 [19.2, 34.3] | 2.6 [0.7, 4.9] | 1.00 |
| +4°C | 64.8 [56, 72.8] | 5.2 [3.1, 7.8] | 1 | 27.5 [20, 35.7] | 3.7 [1.1, 6.9] | 1.00 |
| +5°C | 66.2 [57.4, 74.2] | 6.7 [3.9, 9.9] | 1 | 28.7 [20.8, 37.4] | 4.9 [1.5, 9] | 1.00 |

**Panel B. Absolute mean nocturnal air temperature**

| **Mean nocturnal air temp.** | **P(TST<7h)  [95% CrI]** | **Difference from 17°C [95% CrI], pp** | **P(dir)** | **P(TST<6h)  [95% CrI]** | **Difference from 17°C [95% CrI], pp** | **P(dir)** |
| --- | --- | --- | --- | --- | --- | --- |
| 11°C | 52.6 [43.3, 61.7] | -6.3 [-9.7, -2.4] | 1 | 22.5 [15.6, 30.7] | -2.2 [-5.6, 2.0] | 0.87 |
| 12C | 53.5 [44.4, 62.3] | -5.4 [-8.1, -2.4] | 1 | 22.6 [15.8, 30.4] | -2.1 [-4.7, 1.0] | 0.92 |
| 13°C | 54.4 [45.4, 63.1] | -4.5 [-6.6, -2.1] | 1 | 22.8 [16.1, 30.4] | -1.9 [-4, 0.5] | 0.95 |
| 14°C | 55.4 [46.5, 63.9] | -3.4 [-5.1, -1.7] | 1 | 23.1 [16.5, 30.6] | -1.6 [-3.3, 0.2] | 0.96 |
| 15°C | 56.5 [47.7, 65] | -2.3 [-3.5, -1.2] | 1 | 23.5 [16.9, 31.1] | -1.1 [-2.3, 0] | 0.98 |
| 16°C | 57.7 [48.9, 66] | -1.2 [-1.8, -0.6] | 1 | 24 [17.4, 31.6] | -0.6 [-1.2, -0.1] | 0.98 |
| 17°C | 58.9 [50.2, 67.1] |  |  | 24.6 [18, 32.3] |  |  |
| 18°C | 60.1 [51.4, 68.2] | 1.2 [0.7, 1.8] | 1 | 25.3 [18.5, 33] | 0.6 [0.1, 1.2] | 0.99 |
| 19°C | 61.3 [52.7, 69.3] | 2.4 [1.4, 3.5] | 1 | 26 [19, 33.7] | 1.3 [0.2, 2.4] | 0.99 |
| 20°C | 62.5 [54, 70.4] | 3.7 [2.1, 5.3] | 1 | 26.7 [19.5, 34.6] | 2 [0.4, 3.8] | 0.99 |
| 21°C | 63.8 [55.3, 71.6] | 4.9 [2.9, 7] | 1 | 27.5 [20.2, 35.7] | 2.8 [0.7, 5.2] | 0.99 |
| 22°C | 65 [56.6, 72.8] | 6.1 [3.7, 8.8] | 1 | 28.4 [20.8, 36.9] | 3.8 [1.1, 6.7] | 1 |
| 23°C | 66.3 [57.9, 74] | 7.4 [4.5, 10.6] | 1 | 29.4 [21.5, 38.4] | 4.8 [1.6, 8.5] | 1 |
| 24°C | 67.5 [58.9, 75.3] | 8.7 [5.2, 12.6] | 1 | 30.5 [22.3, 40.1] | 5.9 [1.9, 10.6] | 1 |

Values are posterior means with 95% credible intervals (CrI). In Panel A, temperature deviation represents the within-person deviation in mean nocturnal air temperature from a participant's usual exposure; 0°C therefore represents the participant-specific usual temperature. In Panel B, posterior probabilities were estimated at selected absolute mean nocturnal air temperatures, with 17°C as the reference. Differences in probability are reported in percentage points (pp). Participant-specific random effects were excluded from predictions, yielding population-level estimates. P(dir) denotes the posterior probability that the difference from the respective reference is in the estimated direction. Short sleep was defined as total sleep time (TST) <7 h or <6 h per night.

**(A)**

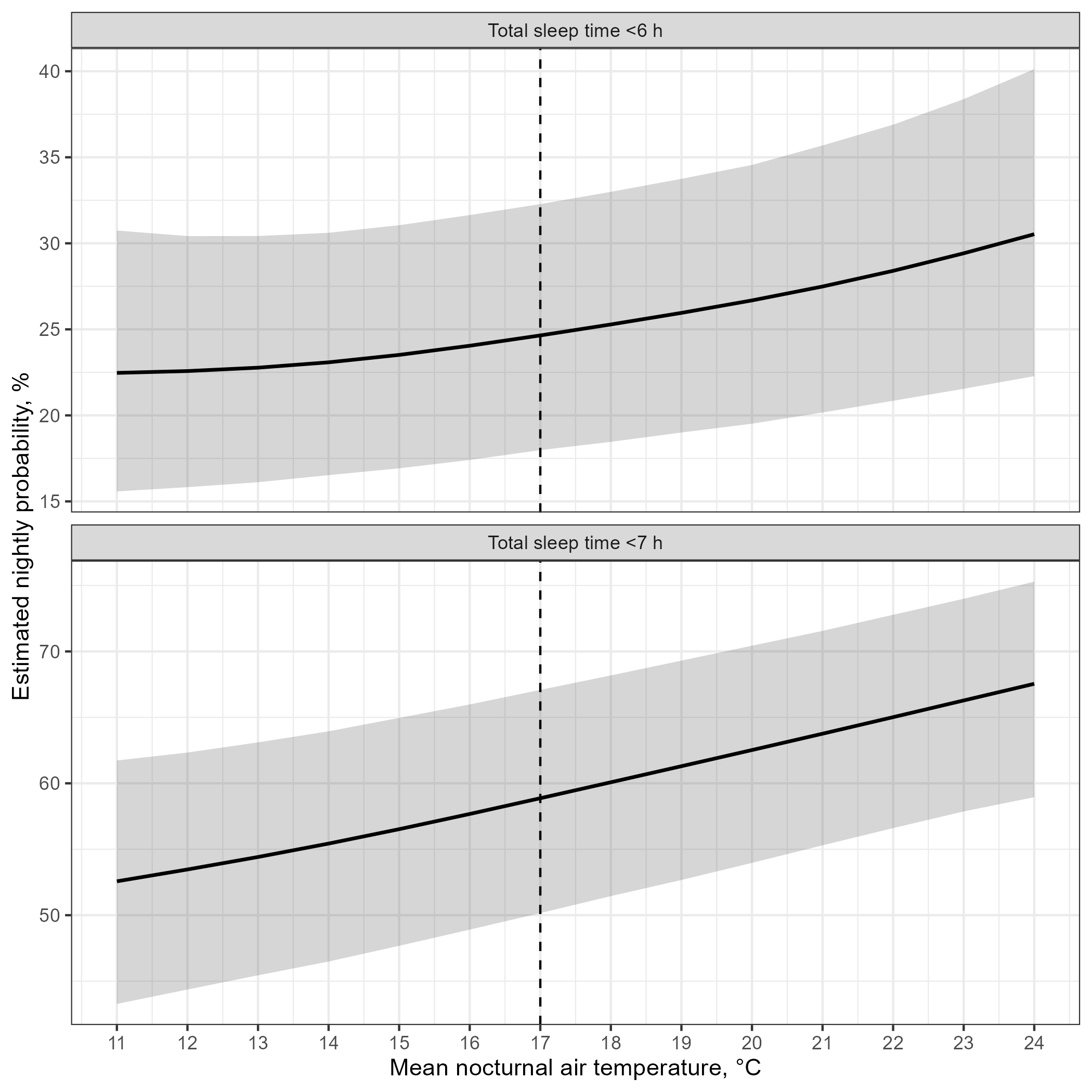

**(B)**

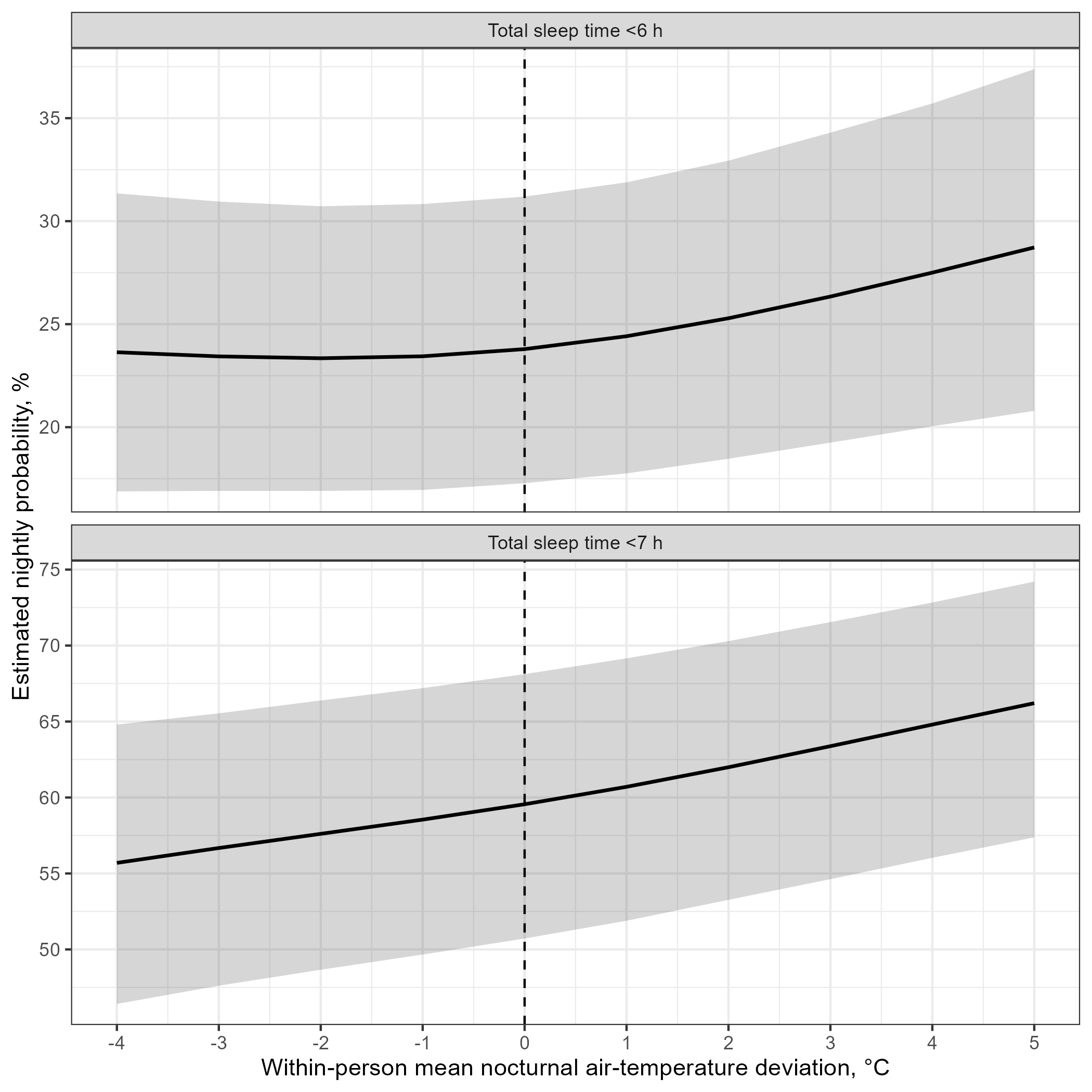

**Supplementary Figure S4. Posterior estimated probabilities of short sleep as a function of mean nocturnal air temperature.** (A) Estimated probability of total sleep time <6 h and (B) <7 h across absolute mean nocturnal air temperatures. (C) Estimated probability of total sleep time <6 h and (D) <7 h across within-person deviations in mean nocturnal air temperature from a participant’s usual exposure. Lines show posterior mean estimates and shaded areas indicate 95% credible intervals. For the absolute-temperature analyses, contrasts were calculated relative to 17°C; for the within-person analyses, 0°C represents a participant’s usual nocturnal temperature exposure. Corresponding numerical estimates and contrasts are provided in Supplementary Table S9.

Between-person nocturnal exposure associations

**Supplementary Table S10.** **Between-person associations between participant-level mean nocturnal wet-bulb exposure and sleep outcomes.**

| **Outcome** | **Model** | **Between-person coefficient, mean [95% CrI]** | **P(direction)** | **Scale** |
| --- | --- | --- | --- | --- |
| Sleep maintenance efficiency | Same-night smooth | 0.08 [-0.04, 0.22] | 0.90 | Percentage points |
|  | Linear lags 0-1 | 0.1 [-0.03, 0.23] | 0.93 |  |
|  | Linear lags 0-2 | 0.12 [-0.03, 0.26] | 0.94 |  |
| Total sleep time | Same-night smooth | -5.47 [-9.00, -1.92] | 1 | Minutes |
|  | Linear lags 0-1 | -4.55 [-9.51, -0.88] | 0.99 |  |
|  | Linear lags 0-2 | -5.38 [-9.22, -1.41] | 1 |  |
| Light sleep | Same-night smooth | -0.26 [-0.71, 0.19] | 0.87 | Percentage points |
|  | Linear lags 0-1 | -0.27 [-0.77, 0.23] | 0.86 |  |
|  | Linear lags 0-2 | -0.29 [-0.83, 0.27] | 0.85 |  |
| Deep sleep | Same-night smooth | 0.27 [-0.13, 0.66] | 0.91 | Percentage points |
|  | Linear lags 0-1 | 0.31 [-0.14, 0.75] | 0.92 |  |
|  | Linear lags 0-2 | 0.41 [-0.11, 0.92] | 0.94 |  |
| REM sleep | Same-night smooth | -0.11 [-0.49, 0.27] | 0.71 | Percentage points |
|  | Linear lags 0-1 | -0.09 [-0.47, 0.30] | 0.68 |  |
|  | Linear lags 0-2 | -0.13 [-0.55, 0.29] | 0.72 |  |
| Recovery index | Same-night smooth | -0.03 [-0.04, -0.02] | 1 | Percentage points |
|  | Linear lags 0-1 | -0.03 [-0.04, -0.02] | 1 |  |
|  | Linear lags 0-2 | -0.03 [-0.04, -0.02] | 1 |  |
| Subjective sleep quality, BPSQI-adjusted | Same-night smooth | 0.08 [-0.07, 0.24] | 0.85 | Scale points |
|  | Linear lags 0-1 | 0.07 [-0.09, 0.23] | 0.81 |  |
|  | Linear lags 0-2 | 0.07 [-0.11, 0.24] | 0.77 |  |

Values are posterior means [95% credible intervals (CrI)] and represent the estimated difference in the sleep outcome associated with a 1°C higher participant-level mean nocturnal wet-bulb temperature, relative to the sample mean. For sleep maintenance efficiency and sleep-stage proportions, differences are reported in percentage points; for total sleep time, recovery index, and subjective sleep quality, differences are reported in minutes, recovery-index units, and scale points, respectively. Higher subjective sleep-quality scores indicate poorer sleep quality. P(direction) denotes the posterior probability that the estimated difference is in the observed direction.

**Supplementary Table S11.** **Between-person associations between participant-level mean nocturnal air temperature exposure and sleep outcomes.**

| **Outcome** | **Model** | **Between-person coefficient, mean [95% CrI]** | **P(direction)** | **Scale** |
| --- | --- | --- | --- | --- |
| Sleep maintenance efficiency | Same-night smooth | 0.07 [-0.03, 0.17] | 0.92 | Percentage points |
|  | Linear lags 0-1 | 0.09 [-0.02, 0.20] | 0.95 |  |
|  | Linear lags 0-2 | 0.09 [-0.02, 0.20] | 0.95 |  |
| Total sleep time | Same-night smooth | -3.76 [-6.61, -1.02] | 1 | Minutes |
|  | Linear lags 0-1 | -3.14 [-5.99, -0.28] | 0.99 |  |
|  | Linear lags 0-2 | -3.76 [-6.82, -0.71] | 0.99 |  |
| Light sleep | Same-night smooth | -0.17 [-0.51, 0.17] | 0.83 | Percentage points |
|  | Linear lags 0-1 | -0.14 [-0.54, 0.26] | 0.75 |  |
|  | Linear lags 0-2 | -0.17 [-0.60, 0.26] | 0.78 |  |
| Deep sleep | Same-night smooth | 0.21 [-0.12, 0.52] | 0.90 | Percentage points |
|  | Linear lags 0-1 | 0.23 [-0.12, 0.58] | 0.90 |  |
|  | Linear lags 0-2 | 0.30 [-0.09, 0.70] | 0.93 |  |
| REM sleep | Same-night smooth | -0.14 [-0.44, 0.15] | 0.83 | Percentage points |
|  | Linear lags 0-1 | -0.19 [-0.51, 0.13] | 0.88 |  |
|  | Linear lags 0-2 | -0.22 [-0.56, 0.11] | 0.91 |  |
| Recovery index | Same-night smooth | -0.03 [-0.04, -0.02] | 1 | Recovery-index units |
|  | Linear lags 0-1 | -0.03 [-0.04, -0.02] | 1 |  |
|  | Linear lags 0-2 | -0.03 [-0.04, -0.02] | 1 |  |
| Subjective sleep quality, BPSQI-adjusted | Same-night smooth | 0.07 [-0.05, 0.19] | 0.88 | Scale points |
|  | Linear lags 0-1 | 0.07 [-0.06, 0.20] | 0.85 |  |
|  | Linear lags 0-2 | 0.07 [-0.07, 0.21] | 0.84 |  |

Values are posterior means [95% credible intervals (CrI)] and represent the estimated difference in the sleep outcome associated with a 1°C higher participant-level mean nocturnal air temperature, relative to the sample mean. For sleep maintenance efficiency and sleep-stage proportions, differences are reported in percentage points; for total sleep time, recovery index, and subjective sleep quality, differences are reported in minutes, recovery-index units, and scale points, respectively. Higher subjective sleep-quality scores indicate poorer sleep quality. P(direction) denotes the posterior probability that the estimated difference is in the observed direction

Covariate effects

**Supplementary Table S12. Posterior summaries of selected covariate effects in Bayesian wet-bulb–sleep models.**

| **Outcome** | **Model** | **Parameter** | **Estimate [95% CrI]** | **P(direction)** | **Scale** |
| --- | --- | --- | --- | --- | --- |
| Sleep maintenance efficiency | Same-night smooth | Gender: female | 0.08 [-0.05, 0.22] | 0.88 | Beta-regression logit scale |
|  | Linear lags 0-1 | Gender: female | 0.07 [-0.08, 0.21] | 0.81 |  |
|  | Linear lags 0-2 | Gender: female | 0.03 [-0.14, 0.19] | 0.63 |  |
| Total sleep time | Same-night smooth | Gender: female | 28.86 [15.26, 42.97] | 1 | Minutes |
|  | Linear lags 0-1 | Gender: female | 28.0 [13.52, 43.02] | 1 |  |
|  | Linear lags 0-2 | Gender: female | 27.96 [12.28, 43.02] | 1 |  |
| Light sleep | Same-night smooth | Gender: female | -0.03 [-0.11, 0.04] | 0.79 | Beta-regression logit scale |
|  | Linear lags 0-1 | Gender: female | -0.03 [-0.11, 0.06] | 0.74 |  |
|  | Linear lags 0-2 | Gender: female | -0.001 [-0.09, 0.09] | 0.52 |  |
| Deep sleep | Same-night smooth | Gender: female | -0.02 [-0.11, 0.08] | 0.68 | Beta-regression logit scale |
|  | Linear lags 0-1 | Gender: female | -0.04 [-0.15, 0.08] | 0.73 |  |
|  | Linear lags 0-2 | Gender: female | -0.05 [-0.17, 0.08] | 0.78 |  |
| REM sleep | Same-night smooth | Gender: female | 0.15 [0.03, 0.26] | 1 | Beta-regression logit scale |
|  | Linear lags 0-1 | Gender: female | 0.16 [0.04, 0.28] | 1 |  |
|  | Linear lags 0-2 | Gender: female | 0.14 [0.02, 0.28] | 0.99 |  |
| Recovery index | Same-night smooth | Gender: female | 0.05 [0.0, 0.09] | 0.98 | Recovery-index units |
|  | Linear lags 0-1 | Gender: female | 0.05 [0.002, 0.10] | 0.98 |  |
|  | Linear lags 0-2 | Gender: female | 0.05 [-0.001, 0.10] | 0.97 |  |
| Subjective sleep quality, BPSQI-adjusted | Same-night smooth | Gender: female | 1.16 [0.58, 1.75] | 1 | Scale points |
|  | Same-night smooth | Baseline BPSQI | 0.40 [0.32, 0.48] | 1 |  |
|  | Linear lags 0-1 | Gender: female | 1.22 [0.59, 1.87] | 1 |  |
|  | Linear lags 0-1 | Baseline BPSQI | 0.41 [0.33, 0.49] | 1 |  |
|  | Linear lags 0-2 | Gender: female | 1.23 [0.53, 1.94] | 1 |  |
|  | Linear lags 0-2 | Baseline BPSQI | 0.43 [0.34, 0.51] | 1 |  |

Values are posterior means with 95% credible intervals. Gender coefficients represent the estimated difference for participants coded as female relative to the model reference category. Baseline BPSQI coefficients represent the estimated difference in daily subjective sleep quality associated with a one-point higher baseline BPSQI score. Baseline BPSQI was included only in models of daily subjective sleep quality. For sleep maintenance efficiency and sleep-stage percentages, coefficients are reported on the beta-regression logit scale and are therefore not directly interpretable as percentage-point differences. Positive coefficients indicate a higher expected proportion, whereas negative coefficients indicate a lower expected proportion. Coefficients for total sleep time, recovery index, and subjective sleep quality are reported in minutes, recovery-index units, and scale points, respectively. Higher subjective sleep-quality scores indicate worse sleep quality. Baseline age was included as a smooth adjustment term and is therefore not summarised as a single coefficient. CrI, credible interval; P(direction), posterior probability that the coefficient is in the estimated direction.

**Supplementary Table S13. Posterior summaries of selected covariate effects in Bayesian air temperature–sleep models.**

| **Outcome** | **Model** | **Parameter** | **Estimate [95% CrI]** | **P(direction)** | **Scale** |
| --- | --- | --- | --- | --- | --- |
| Sleep maintenance efficiency | Same-night smooth | Gender: female | 0.08 [-0.05, 0.22] | 0.88 | Beta-regression logit scale |
|  | Linear lags 0-1 | Gender: female | 0.07 [-0.08, 0.21] | 0.81 |  |
|  | Linear lags 0-2 | Gender: female | 0.03 [-0.14, 0.19] | 0.63 |  |
| Total sleep time | Same-night smooth | Gender: female | 29.5 [15.95, 43.18] | 1 | Minutes |
|  | Linear lags 0-1 | Gender: female | 28.21 [13.33, 43.22] | 1 |  |
|  | Linear lags 0-2 | Gender: female | 27.77 [12.57, 43.20] | 1 |  |
| Light sleep | Same-night smooth | Gender: female | -0.03 [-0.10, 0.05] | 0.78 | Beta-regression logit scale |
|  | Linear lags 0-1 | Gender: female | -0.03 [-0.11, 0.06] | 0.72 |  |
|  | Linear lags 0-2 | Gender: female | -0.001 [-0.09, 0.09] | 0.51 |  |
| Deep sleep | Same-night smooth | Gender: female | -0.02 [-0.11, 0.08] | 0.65 | Beta-regression logit scale |
|  | Linear lags 0-1 | Gender: female | -0.04 [-0.15, 0.08] | 0.73 |  |
|  | Linear lags 0-2 | Gender: female | -0.05 [-0.18, 0.08] | 0.77 |  |
| REM sleep | Same-night smooth | Gender: female | 0.14 [0.03, 0.26] | 0.99 | Beta-regression logit scale |
|  | Linear lags 0-1 | Gender: female | 0.16 [0.03, 0.27] | 0.99 |  |
|  | Linear lags 0-2 | Gender: female | 0.14 [0.01, 0.27] | 0.98 |  |
| Recovery index | Same-night smooth | Gender: female | 0.04 [0.002, 0.09] | 0.98 | Recovery-index units |
|  | Linear lags 0-1 | Gender: female | 0.05 [0.002, 0.10] | 0.98 |  |
|  | Linear lags 0-2 | Gender: female | 0.05 [-0.001, 0.10] | 0.97 |  |
| Subjective sleep quality, BPSQI-adjusted | Same-night smooth | Gender: female | 1.19 [0.59, 1.80] | 1 | Scale points |
|  | Same-night smooth | Baseline BPSQI | 0.4 [0.32, 0.48] | 1 |  |
|  | Linear lags 0-1 | Gender: female | 1.22 [0.57, 1.86] | 1 |  |
|  | Linear lags 0-1 | Baseline BPSQI | 0.41 [0.33, 0.49] | 1 |  |
|  | Linear lags 0-2 | Gender: female | 1.25 [0.56, 1.94] | 1 |  |
|  | Linear lags 0-2 | Baseline BPSQI | 0.43 [0.34, 0.51] | 1 |  |

Values are posterior means with 95% credible intervals. Gender coefficients represent the estimated difference for participants coded as female relative to the model reference category. Baseline BPSQI coefficients represent the estimated difference in daily subjective sleep quality associated with a one-point higher baseline BPSQI score. Baseline BPSQI was included only in models of daily subjective sleep quality. For sleep maintenance efficiency and sleep-stage percentages, coefficients are reported on the beta-regression logit scale and are therefore not directly interpretable as percentage-point differences. Positive coefficients indicate a higher expected proportion, whereas negative coefficients indicate a lower expected proportion. Coefficients for total sleep time, recovery index, and subjective sleep quality are reported in minutes, recovery-index units, and scale points, respectively. Higher subjective sleep-quality scores indicate worse sleep quality. Baseline age was included as a smooth adjustment term and is therefore not summarised as a single coefficient. CrI, credible interval; P(direction), posterior probability that the coefficient is in the estimated direction.

**Supplementary Figure S5. Adjusted associations between baseline age and sleep outcomes in the primary Bayesian same-night models.**

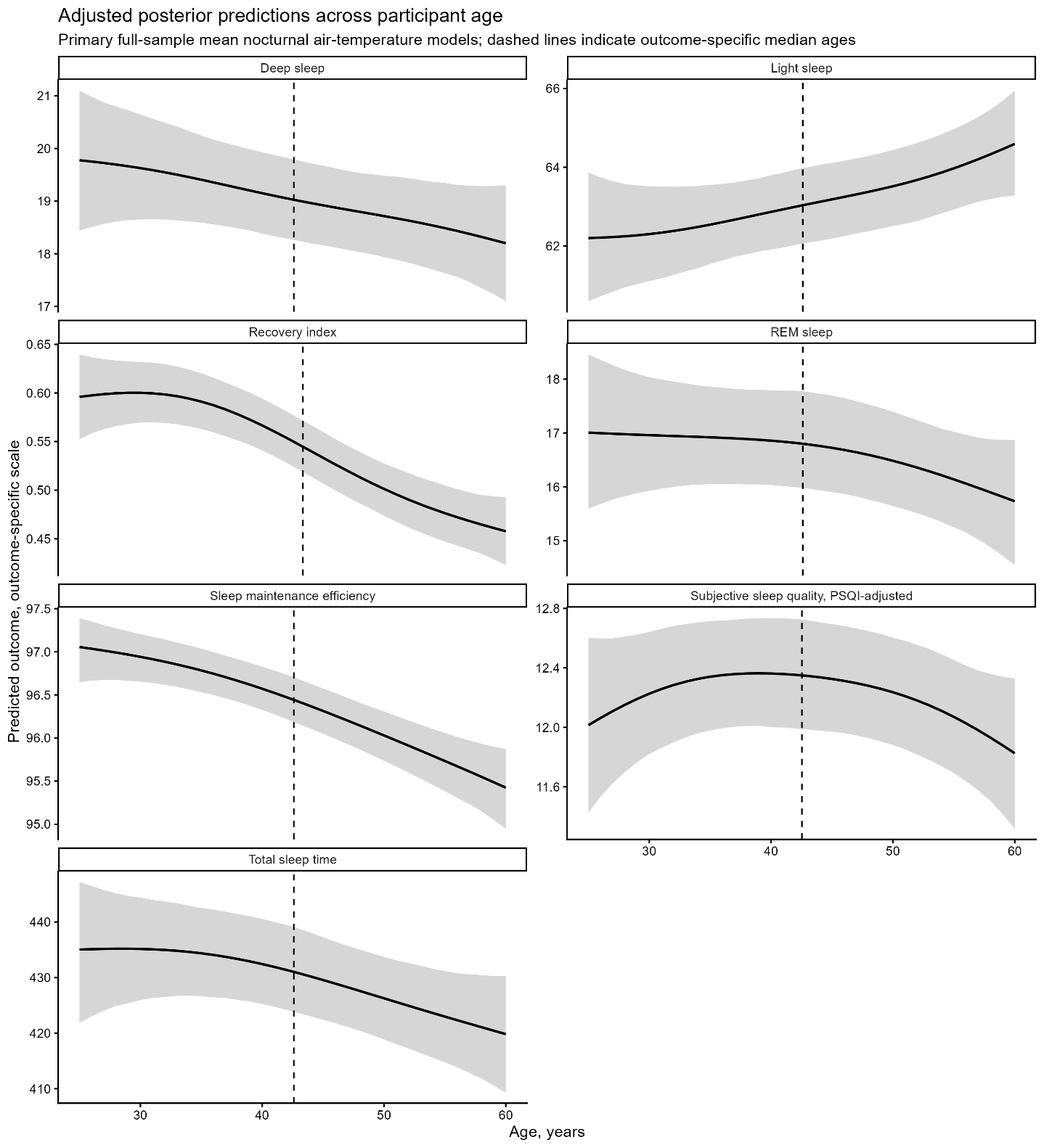

Curves show posterior mean predictions across the observed age range, with shaded areas indicating 95% credible intervals. Predictions were obtained from the smooth age term while holding the remaining covariates constant. Age associations represent overall between-participant differences and should not be interpreted as modification of temperature-related sleep responses.

**Supplementary Table S14. Posterior contrasts in sleep outcomes at selected ages in the primary Bayesian same-night models.**

| **Outcome** | **Age point** | **Age (years)** | **Predicted outcome [95% CrI]** | **Difference from median age, mean [95% CrI]** | **Scale** |
| --- | --- | --- | --- | --- | --- |
| Sleep maintenance efficiency | 25th percentile | 34.9 | 96.79 [96.54, 97.04] | 0.35 [0.14, 0.56] | Percentage points |
|  | Median | 42.6 | 96.44 [96.19, 96.7] | Reference |  |
|  | 75th percentile | 53.0 | 95.86 [95.53, 96.16] | -0.59 [-0.91, -0.32] |  |
| Total sleep time | 25th percentile | 34.9 | 434.41 [426.67, 442.56] | 3.38 [-2.72, 9.59] | Minutes |
|  | Median | 42.6 | 431.02 [423.83, 439.1] | Reference |  |
|  | 75th percentile | 53.0 | 424.31 [416.46, 432.15] | -6.71 [-15.55, 0.1] |  |
| Light sleep | 25th percentile | 34.9 | 62.53 [61.53, 63.55] | -0.5 [-1.44, 0.38] | Percentage points |
|  | Median | 42.6 | 63.03 [62.06, 63.98] | Reference |  |
|  | 75th percentile | 53.0 | 63.76 [62.72, 64.71] | 0.73 [-0.4, 1.73] |  |
| Deep sleep | 25th percentile | 34.9 | 19.41 [18.6, 20.26] | 0.39 [-0.23, 1.12] | Percentage points |
|  | Median | 42.6 | 19.03 [18.26, 19.79] | Reference |  |
|  | 75th percentile | 53.0 | 18.59 [17.77, 19.4] | -0.44 [-1.2, 0.42] |  |
| REM sleep | 25th percentile | 34.9 | 16.92 [16.05, 17.86] | 0.12 [-0.75, 0.79] | Percentage points |
|  | Median | 42.6 | 16.8 [15.98, 17.77] | Reference |  |
|  | 75th percentile | 53.0 | 16.29 [15.44, 17.16] | -0.51 [-1.47, 0.33] |  |
| Recovery index | 25th percentile | 35.0 | 0.59 [0.56, 0.62] | 0.05 [0.02, 0.08] | Recovery-index units |
|  | Median | 43.3 | 0.54 [0.52, 0.57] | Reference |  |
|  | 75th percentile | 53.2 | 0.48 [0.46, 0.51] | -0.06 [-0.09, -0.03] |  |
| Subjective sleep quality, BPSQI-adjusted | 25th percentile | 34.7 | 12.34 [11.98, 12.71] | -0.01 [-0.33, 0.3] | Scale points |
|  | Median | 42.5 | 12.35 [11.99, 12.72] | Reference |  |
|  | 75th percentile | 52.9 | 12.15 [11.78, 12.53] | -0.2 [-0.61, 0.17] |  |

Values are posterior mean differences [95% credible intervals] relative to the sample median age. Contrasts were derived from the penalised smooth of baseline age and are presented at selected ages to facilitate interpretation of the potentially non-linear associations.

Exploratory lag analyses

**Supplementary Table S15.** **Number of participants and observations included in the primary same-night and exploratory linear lag models, by outcome and lag window.**

| **Model** | **Lag (days)** | ***N* participants** | ***N* nights/ values** |
| --- | --- | --- | --- |
| Sleep Maintenance | 0 | 309 | 11547 |
| Efficiency | 1 | 297 | 8940 |
|  | 2 | 287 | 7117 |
| Total sleep time | 0 | 309 | 11547 |
|  | 1 | 297 | 8940 |
|  | 2 | 287 | 7117 |
| Light Sleep | 0 | 309 | 11547 |
|  | 1 | 297 | 8940 |
|  | 2 | 287 | 7117 |
| Deep Sleep | 0 | 309 | 11547 |
|  | 1 | 297 | 8940 |
|  | 2 | 287 | 7117 |
| REM Sleep | 0 | 309 | 11547 |
|  | 1 | 297 | 8940 |
|  | 2 | 287 | 7117 |
| Recovery Index | 0 | 294 | 10140 |
|  | 1 | 281 | 7949 |
|  | 2 | 273 | 6445 |
| Subjective Sleep | 0 | 326 | 14290 |
| Quality | 1 | 320 | 10706 |
|  | 2 | 307 | 8407 |

Lag 0 refers to the primary same-night dataset. Lag 1 refers to the complete-case dataset used for models simultaneously including temperature at Lags 0 and 1, and Lag 2 refers to the dataset used for models simultaneously including temperature at Lags 0, 1, and 2. Lagged exposures were retained only when the corresponding sleep dates were consecutive. Consequently, models with longer lag windows required more complete exposure histories and included fewer participants and observations. Observations represent participant-nights for sleep outcomes and participant-day sleep-diary assessments for subjective sleep quality.

**Supplementary Table S16. Posterior estimates of associations between within-person nocturnal air temperature at lags 0-2 and sleep outcomes in exploratory linear lag models.**

**Panel A. Sleep maintenance efficiency and sleep-stage proportions (percentage points)**

| **Outcome** | **Lags included** | **Lag 0** | **Lag 1** | **Lag 2** |
| --- | --- | --- | --- | --- |
| Sleep maintenance efficiency | Lags 0-1 | -0.03 [-0.06, 0.01]; 0.94 | 0.03 [-0.01, 0.06]; 0.94 |  |
|  | Lags 0-2 | -0.03 [-0.06, 0.01]; 0.93 | 0.02 [-0.02, 0.06]; 0.87 | 0.003 [-0.03, 0.04]; 0.57 |
| Light sleep | Lags 0-1 | 0.10 [0.01, 0.19]; 0.99 | -0.10 [-0.19, -0.01]; 0.98 |  |
|  | Lags 0-2 | 0.12 [0.02, 0.22]; 0.99 | -0.09 [-0.21, 0.03]; 0.92 | -0.01 [-0.11, 0.09]; 0.58 |
| Deep sleep | Lags 0-1 | 0.09 [0.02, 0.16]; 0.99 | -0.01 [-0.08, 0.06]; 0.65 |  |
|  | Lags 0-2 | 0.07 [-0.004, 0.15]; 0.97 | -0.06 [-0.15, 0.03]; 0.91 | 0.08 [0.005, 0.16]; 0.98 |
| REM sleep | Lags 0-1 | -0.22 [-0.30, -0.14]; 1 | 0.11 [0.03, 0.19]; 1 |  |
|  | Lags 0-2 | -0.25 [-0.34, -0.15]; 1 | 0.19 [0.08, 0.30]; 1 | -0.14 [-0.23, -0.05]; 1 |

**Panel B. Total sleep time, recovery index, and subjective sleep quality**

| **Outcome** | **Lags included** | **Lag 0** | **Lag 1** | **Lag 2** |
| --- | --- | --- | --- | --- |
| Recovery index | Lags 0-1 | -0.01 [-0.01, -0.004]; 1 | 0.004 [0.001, 0.01]; 1 |  |
|  | Lags 0-2 | -0.01 [-0.01, -0.004]; 1 | 0.003 [0.0, 0.01]; 0.97 | 0.001 [-0.002, 0.003]; 0.73 |
| Total Sleep Time | Lags 0-1 | -2.19 [-2.94, -1.42]; 1 | 0.64 [-0.11, 1.38]; 0.95 |  |
|  | Lags 0-2 | -2.03 [-2.85, -1.19]; 1 | 0.98 [-0.01, 1.98]; 0.97 | -0.64 [-1.43, 0.18]; 0.93 |
| Subjective sleep quality, B-PSQI-adjusted | Lags 0-1 | 0.13 [0.10, 0.17]; 1 | 0.025 [-0.010, 0.061]; 0.92 |  |
|  | Lags 0-2 | 0.15 [0.11, 0.19]; 1 | -0.02 [-0.07, 0.02]; 0.83 | 0.06 [0.03, 0.10]; 1 |

Values are posterior expected differences [95% credible intervals (CrI)]; P(direction). Lag 0 denotes nocturnal temperature during the night of the sleep outcome, whereas Lags 1 and 2 denote temperatures one and two nights earlier. Models labelled Lags 0-1 and Lags 0-2 simultaneously include all temperature lags within the indicated range. Each estimate represents the expected difference in the sleep outcome associated with a 1°C higher within-person nocturnal temperature at the respective lag, relative to a 0°C deviation, while the other temperature lags included in the model are held at 0°C. For sleep maintenance efficiency and sleep-stage proportions, differences are reported in percentage points; for total sleep time, recovery index, and subjective sleep quality, differences are reported in minutes, recovery-index units, and scale points, respectively. Higher subjective sleep-quality scores indicate poorer sleep quality. Participant-specific random effects were excluded from predictions, yielding population-level expected contrasts. P(direction) denotes the posterior probability that the estimated difference is in the observed direction. Models with longer lag windows require complete consecutive exposure histories and may therefore include different numbers of observations.

**Supplementary Table S17. Posterior estimates of associations between within-person nocturnal wet-bulb temperature at lags 0-2 and sleep outcomes in exploratory linear lag models.**

**Panel A. Sleep maintenance efficiency and sleep-stage proportions (percentage points)**

| **Outcome** | **Lags included** | **Lag 0** | **Lag 1** | **Lag 2** |
| --- | --- | --- | --- | --- |
| Sleep maintenance efficiency | Lags 0-1 | -0.02 [-0.05, 0.02]; 0.85 | 0.02 [-0.01, 0.06]; 0.90 |  |
|  | Lags 0-2 | -0.03 [-0.06, 0.01]; 0.91 | 0.04 [-0.01, 0.08]; 0.96 | -0.02 [-0.06, 0.01]; 0.90 |
| Light sleep | Lags 0-1 | 0.11 [0.01, 0.21]; 0.99 | -0.12 [-0.22, -0.03]; 0.99 |  |
|  | Lags 0-2 | 0.16 [0.05, 0.27]; 1 | -0.19 [-0.32, -0.06]; 1 | 0.09 [-0.02, 0.20]; 0.95 |
| Deep sleep | Lags 0-1 | 0.08 [0.01, 0.16]; 0.99 | -0.03 [-0.10, 0.05]; 0.77 |  |
|  | Lags 0-2 | 0.05 [-0.03, 0.13]; 0.88 | -0.01 [-0.11, 0.09]; 0.57 | 0.01 [-0.08, 0.09]; 0.57 |
| REM sleep | Lags 0-1 | -0.21 [-0.30, -0.13]; 1 | 0.15 [0.07, 0.24]; 1 |  |
|  | Lags 0-2 | -0.25 [-0.35, -0.16]; 1 | 0.25 [0.14, 0.38]; 1 | -0.16 [-0.26, -0.07]; 1 |

**Panel B. Total sleep time, recovery index, and subjective sleep quality**

| **Outcome** | **Lags included** | **Lag 0** | **Lag 1** | **Lag 2** |
| --- | --- | --- | --- | --- |
| Recovery index | Lags 0-1 | -0.01 [-0.01, -0.003]; 1 | 0.01 [0.002, 0.01]; 1 |  |
|  | Lags 0-2 | -0.01 [-0.01, -0.003]; 1 | 0.004 [0.0, 0.01]; 0.98 | 0.001 [-0.002, 0.004]; 0.77 |
| Total Sleep Time | Lags 0-1 | -2.40 [-3.21, -1.60]; 1 | 1.20 [0.40, 1.99]; 1 |  |
|  | Lags 0-2 | -2.47 [-3.37, -1.56]; 1 | 1.63 [0.54, 2.75]; 1 | -0.66 [-1.55, 0.24]; 0.92 |
| Subjective sleep quality, B-PSQI-adjusted | Lags 0-1 | 0.15 [0.11, 0.19]; 1 | -0.01 [-0.05, 0.02]; 0.77 |  |
|  | Lags 0-2 | 0.17 [0.13, 0.21]; 1 | -0.05 [-0.10, 0.004]; 0.97 | 0.05 [0.01, 0.09]; 0.99 |

Values are posterior expected differences [95% credible intervals (CrI)]; P(direction). Lag 0 denotes nocturnal temperature during the night of the sleep outcome, whereas Lags 1 and 2 denote temperatures one and two nights earlier. Models labelled Lags 0-1 and Lags 0-2 simultaneously include all temperature lags within the indicated range. Each estimate represents the expected difference in the sleep outcome associated with a 1°C higher within-person nocturnal temperature at the respective lag, relative to a 0°C deviation, while the other temperature lags included in the model are held at 0°C. For sleep maintenance efficiency and sleep-stage proportions, differences are reported in percentage points; for total sleep time, recovery index, and subjective sleep quality, differences are reported in minutes, recovery-index units, and scale points, respectively. Higher subjective sleep-quality scores indicate poorer sleep quality. Participant-specific random effects were excluded from predictions, yielding population-level expected contrasts. P(direction) denotes the posterior probability that the estimated difference is in the observed direction. Models with longer lag windows require complete consecutive exposure histories and may therefore include different numbers of observations.

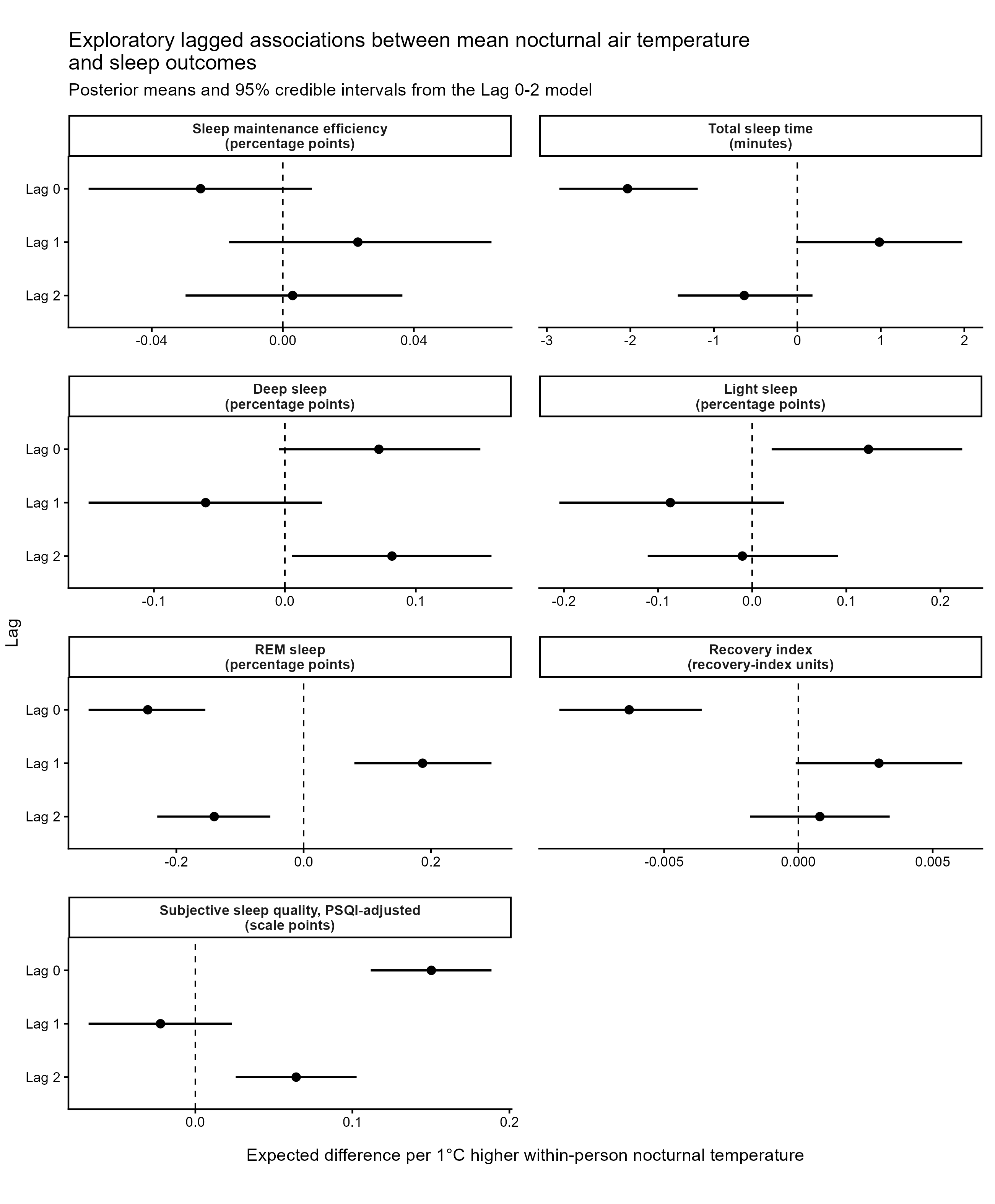

**Supplementary Figure S6. Exploratory lagged associations between mean nocturnal air temperature and sleep outcomes.** Forest plots show posterior mean expected differences and 95% credible intervals from the exploratory linear lag model including same-night air temperature exposure and lags 1-2. Each estimate represents the expected difference in the sleep outcome associated with a 1°C higher within-person nocturnal wet-bulb exposure at the respective lag, relative to a 0°C deviation, while the other lag terms are held at 0°C. Participant-level mean wet-bulb exposure was set to the sample mean, and participant-specific random effects were excluded, yielding population-level expected contrasts. Differences in sleep maintenance efficiency and sleep-stage proportions are reported in percentage points; differences in recovery index, total sleep time, and subjective sleep quality are reported in recovery-index units, minutes, and scale points, respectively. Higher subjective sleep-quality scores indicate worse sleep quality. Exploratory lag models were used to assess potential carry-over associations and were not designed to estimate non-linear distributed lag exposure-response functions.

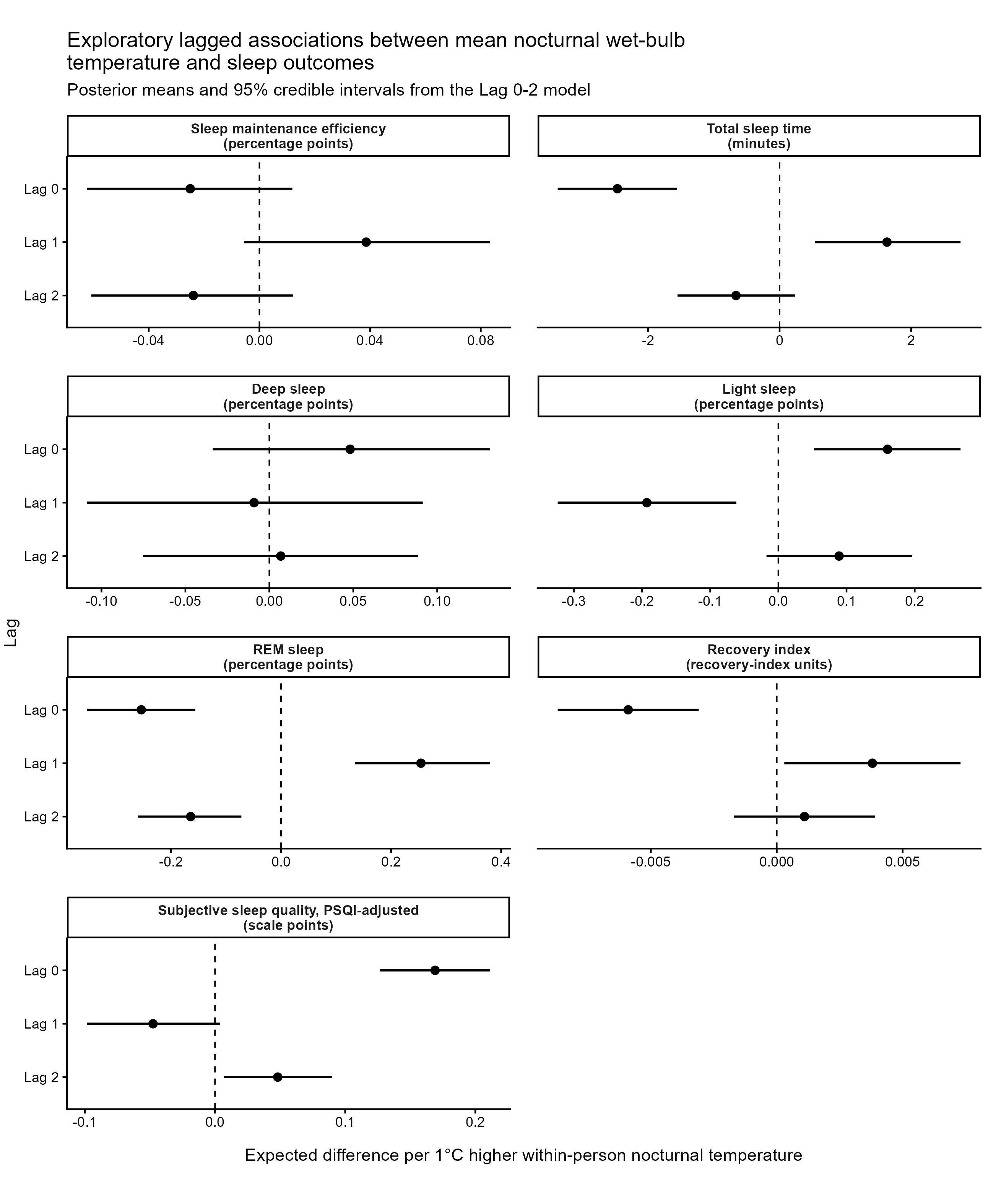

**Supplementary Figure S7. Exploratory lagged associations between mean nocturnal wet-bulb temperature and sleep outcomes.** Forest plots show posterior mean expected differences and 95% credible intervals from the exploratory linear lag model including same-night wet-bulb exposure and lags 1-2. Each estimate represents the expected difference in the sleep outcome associated with a 1°C higher within-person nocturnal wet-bulb exposure at the respective lag, relative to a 0°C deviation, while the other lag terms are held at 0°C. Participant-level mean wet-bulb exposure was set to the sample mean, and participant-specific random effects were excluded, yielding population-level expected contrasts. Differences in sleep maintenance efficiency and sleep-stage proportions are reported in percentage points; differences in recovery index, total sleep time, and subjective sleep quality are reported in recovery-index units, minutes, and scale points, respectively. Higher subjective sleep-quality scores indicate worse sleep quality. Exploratory lag models were used to assess potential carry-over associations and were not designed to estimate non-linear distributed lag exposure-response functions.

Weekly Mental Health Analyses

**Supplementary Table S18**. **Posterior expected mental health outcomes and differences from usual nocturnal temperature exposure.**

**Panel A. Mean nocturnal air temperature**

| **Outcome** | **Unit** | **Air temperature deviation** | **Predicted outcome [95% CrI]** | **Difference from usual night, mean [95% CrI]** | **P(direction)** |
| --- | --- | --- | --- | --- | --- |
| Mental health | Scale points | -4°C | 76.14 [71.87, 80.39] | 0.30 [-1.56, 2.53] | 0.59 |
|  |  | -3°C | 76.18 [72.20, 80.12] | 0.33 [-0.96, 1.94] | 0.65 |
|  |  | -2°C | 76.09 [72.22, 79.89] | 0.25 [-0.65, 1.49] | 0.64 |
|  |  | -1°C | 75.91 [72.04, 79.63] | 0.07 [-0.36, 0.67] | 0.56 |
|  |  | 0°C reference | 75.84 [71.99, 79.60] |  |  |
|  |  | +1°C | 76.05 [72.19, 79.84] | 0.21 [-0.21, 0.73] | 0.82 |
|  |  | +2°C | 76.41 [72.46, 80.28] | 0.57 [-0.25, 1.65] | 0.90 |
|  |  | +3°C | 76.74 [72.75, 80.72] | 0.90 [-0.31, 2.32] | 0.92 |
|  |  | +4°C | 77.02 [72.77, 81.25] | 1.17 [-0.66, 3.35] | 0.89 |
|  |  | +5°C | 77.28 [72.47, 82.02] | 1.43 [-1.26, 4.58] | 0.85 |

**Panel B. Mean nocturnal wet-bulb temperature**

| **Outcome** | **Unit** | **Wet-bulb temperature deviation** | **Predicted outcome [95% CrI]** | **Difference from usual night, mean [95% CrI]** | **P(direction)** |
| --- | --- | --- | --- | --- | --- |
| Mental health | Scale points | -4°C | 76.28 [72.17, 80.45] | 0.38 [-1.42, 2.60] | 0.62 |
|  |  | -3°C | 76.19 [72.32, 80.07] | 0.29 [-0.89, 1.70] | 0.64 |
|  |  | -2°C | 76.04 [72.25, 79.79] | 0.14 [-0.64, 1.14] | 0.59 |
|  |  | -1°C | 75.89 [72.11, 79.62] | -0.01 [-0.42, 0.45] | 0.55 |
|  |  | 0°C reference | 75.90 [72.14, 79.62] |  |  |
|  |  | +1°C | 76.13 [72.37, 79.86] | 0.23 [-0.21, 0.84] | 0.83 |
|  |  | +2°C | 76.43 [72.62, 80.21] | 0.53 [-0.28, 1.58] | 0.88 |
|  |  | +3°C | 76.70 [72.78, 80.65] | 0.80 [-0.53, 2.42] | 0.87 |
|  |  | +4°C | 76.95 [72.70, 81.28] | 1.05 [-1.08, 3.62] | 0.83 |
|  |  | +5°C | 77.20 [72.40, 82.14] | 1.30 [-1.79, 4.90] | 0.81 |

Values are posterior means with 95% credible intervals. Deviation refers to the within-person deviation in nocturnal temperature exposure averaged across the preceding seven nights from a participant’s own usual exposure. A deviation of 0°C therefore represents the participant’s usual preceding-week nocturnal temperature exposure. Differences in weekly mental-health scores are expressed relative to this 0°C reference in scale points. P(direction) is the posterior probability that the expected difference from usual exposure is in the estimated direction; for positive estimates, this is the probability that the difference is above zero, and for negative estimates, it is the probability that the difference is below zero. CrI, credible interval

Effect of Age

**Supplementary Table S19. Posterior probabilities of short sleep in relation to mean nocturnal air temperature.**

| **Outcome** | **Age P25 / P75, years** | **Unit** | **+1°C** | **+3°C** | **+5°C** |
| --- | --- | --- | --- | --- | --- |
| Sleep maintenance efficiency | 34.9 / 53.0 | percentage points | 0.02 [-0.04, 0.12]; 0.714 | 0.08 [-0.12, 0.35]; 0.734 | 0.09 [-0.25, 0.50]; 0.678 |
| Total sleep time | 34.9 / 53.0 | min | -1.37 [-3.31, 0.57]; 0.919 | -3.19 [-8.60, 2.80]; 0.872 | -4.35 [-14.20, 6.37]; 0.810 |
| Light sleep | 34.9 / 53.0 | percentage points | -0.05 [-0.33, 0.16]; 0.618 | 0.13 [-0.54, 0.69]; 0.683 | 0.73 [-0.25, 1.78]; 0.925 |
| Deep sleep | 34.9 / 53.0 | percentage points | 0.30 [0.08, 0.55]; 0.998 | 0.55 [-0.03, 1.18]; 0.968 | 0.27 [-0.70, 1.27]; 0.707 |
| REM sleep | 34.9 / 53.0 | percentage points | -0.30 [-0.52, -0.05]; 0.991 | -0.85 [-1.48, -0.13]; 0.988 | -1.49 [-2.58, -0.34]; 0.994 |
| Recovery index | 35.0 / 53.2 | units | 0.00 [0.00, 0.01]; 0.668 | 0.00 [-0.01, 0.02]; 0.701 | 0.01 [-0.02, 0.03]; 0.659 |
| Subjective sleep quality, B-PSQI-adjusted | 34.7 / 52.9 | scale points | 0.07 [-0.03, 0.17]; 0.926 | 0.24 [-0.05, 0.50]; 0.949 | 0.49 [0.04, 0.92]; 0.984 |

Age modification of the association between mean nocturnal air temperature and sleep. Values are posterior mean differences [95% CrI] in the temperature-response contrast between the outcome-specific 75th (P75) and 25th (P25) percentiles of the age distribution. Median age was 42.6 years for objective sleep measures, 43.3 years for recovery index, and 42.5 years for subjective sleep quality. Note that slight differences in age were due to data availability for these measures. Positive values indicate a more positive, or less negative, temperature-associated change at the older age, whereas negative values indicate a more negative, or less positive, change. Contrasts are shown for +1°C, +3°C, and +5°C relative to each participant's usual nocturnal air temperature. For sleep maintenance efficiency and sleep-stage proportions, differences are expressed in percentage points; for total sleep time, recovery index, and subjective sleep quality, differences are expressed in minutes, recovery-index units, and scale points, respectively. Higher subjective sleep-quality scores indicate worse sleep quality. P(direction) denotes the posterior probability that the estimated age-modification contrast is in the observed direction.

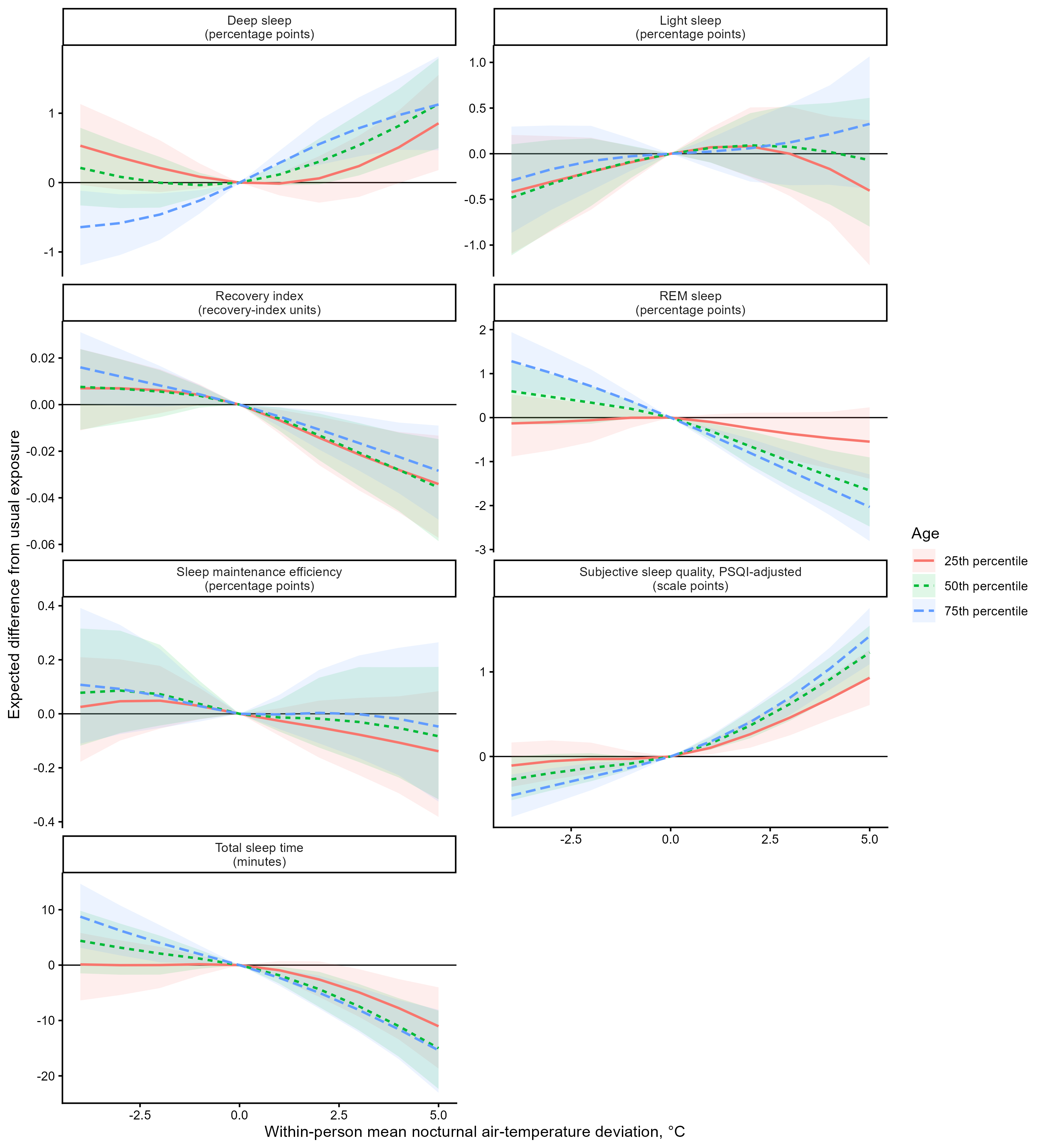

**Supplementary Figure S8. Age-specific associations between within-person nocturnal air-temperature deviations and sleep outcomes.** Lines show posterior mean expected differences in each sleep outcome relative to a participant’s usual nocturnal air-temperature exposure (0°C deviation), evaluated at the 25th, 50th, and 75th percentiles of age. Shaded areas indicate 95% credible intervals. Sleep maintenance efficiency and sleep-stage outcomes are presented as percentage-point differences, total sleep time in minutes, recovery index in recovery-index units, and subjective sleep quality in scale points. Higher subjective sleep-quality scores indicate poorer sleep quality. The figure illustrates the age-by-temperature interaction, with all other model covariates held constant; subjective sleep quality was additionally adjusted for baseline B-PSQI.

Intervention Effects

**Supplementary Table S20**. **Posterior estimates of intervention effect modification of the association between nocturnal temperature and sleep outcomes.**

**Panel A. Mean nocturnal air temperature**

| **Outcome** | **Unit** | **Air temp. deviation** | **Control: difference from usual [95% CrI]** | **Treatment: difference from usual [95% CrI]** | **Treatment - Control difference [95% CrI]** | **P**  **(dir)** |
| --- | --- | --- | --- | --- | --- | --- |
| Sleep maintenance efficiency | Percent-age points | -4°C | 0.00 [-0.20, 0.17] | 0.10 [-0.10, 0.32] | 0.10 [-0.13, 0.38] | 0.77 |
|  |  | -3°C | 0.02 [-0.13, 0.15] | 0.09 [-0.07, 0.29] | 0.08 [-0.09, 0.30] | 0.78 |
|  |  | -2°C | 0.02 [-0.08, 0.12] | 0.07 [-0.05, 0.21] | 0.05 [-0.07, 0.21] | 0.76 |
|  |  | -1°C | 0.01 [-0.04, 0.06] | 0.03 [-0.03, 0.10] | 0.02 [-0.04, 0.09] | 0.73 |
|  |  | 0°C ref |  |  |  |  |
|  |  | +1°C | -0.01 [-0.06, 0.03] | -0.01 [-0.07, 0.04] | 0.00 [-0.06, 0.06] | 0.55 |
|  |  | +2°C | -0.03 [-0.12, 0.07] | -0.02 [-0.12, 0.11] | 0.01 [-0.10, 0.15] | 0.55 |
|  |  | +3°C | -0.05 [-0.19, 0.08] | -0.02 [-0.17, 0.16] | 0.03 [-0.13, 0.25] | 0.61 |
|  |  | +4°C | -0.09 [-0.28, 0.08] | -0.03 [-0.23, 0.18] | 0.06 [-0.17, 0.34] | 0.64 |
|  |  | +5°C | -0.14 [-0.40, 0.08] | -0.07 [-0.32, 0.21] | 0.07 [-0.22, 0.44] | 0.65 |
| Total sleep time | min | -4°C | 5.95 [0.70, 10.77] | 2.81 [-2.41, 7.98] | -3.14 [-9.05, 2.92] | 0.86 |
|  |  | -3°C | 4.17 [-0.31, 8.06] | 1.37 [-3.21, 5.57] | -2.80 [-7.74, 1.63] | 0.90 |
|  |  | -2°C | 2.73 [-0.56, 5.43] | 0.59 [-2.96, 3.62] | -2.15 [-5.94, 0.80] | 0.92 |
|  |  | -1°C | 1.47 [-0.13, 2.77] | 0.30 [-1.45, 1.82] | -1.17 [-3.09, 0.29] | 0.94 |
|  |  | 0°C ref |  |  |  |  |
|  |  | +1°C | -2.04 [-3.19, -0.75] | -0.89 [-2.24, 0.66] | 1.15 [-0.21, 2.88] | 0.95 |
|  |  | +2°C | -4.79 [-7.24, -2.36] | -2.68 [-5.45, 0.35] | 2.11 [-0.65, 5.54] | 0.93 |
|  |  | +3°C | -8.16 [-11.97, -4.75] | -5.33 [-9.40, -1.21] | 2.82 [-1.41, 7.45] | 0.91 |
|  |  | +4°C | -11.89 [-16.90, -7.29] | -8.55 [-13.63, -3.52] | 3.34 [-2.40, 9.15] | 0.88 |
|  |  | +5°C | -15.76 [-22.22, -9.46] | -12.02 [-18.58, -5.54] | 3.74 [-4.01, 11.33] | 0.84 |
| Light sleep | Percent-age points | -4°C | -0.11 [-0.64, 0.37] | -0.45 [-1.03, 0.12] | -0.33 [-1.00, 0.31] | 0.85 |
|  |  | -3°C | -0.06 [-0.44, 0.32] | -0.28 [-0.73, 0.21] | -0.22 [-0.72, 0.30] | 0.82 |
|  |  | -2°C | -0.02 [-0.28, 0.26] | -0.15 [-0.47, 0.22] | -0.13 [-0.46, 0.25] | 0.78 |
|  |  | -1°C | 0.00 [-0.13, 0.14] | -0.06 [-0.22, 0.12] | -0.06 [-0.22, 0.13] | 0.77 |
|  |  | 0°C ref |  |  |  |  |
|  |  | +1°C | 0.00 [-0.14, 0.11] | 0.05 [-0.11, 0.20] | 0.05 [-0.12, 0.22] | 0.76 |
|  |  | +2°C | -0.01 [-0.27, 0.23] | 0.10 [-0.23, 0.41] | 0.11 [-0.24, 0.45] | 0.76 |
|  |  | +3°C | -0.02 [-0.40, 0.33] | 0.15 [-0.31, 0.59] | 0.17 [-0.34, 0.67] | 0.77 |
|  |  | +4°C | -0.03 [-0.52, 0.42] | 0.20 [-0.37, 0.74] | 0.23 [-0.42, 0.88] | 0.77 |
|  |  | +5°C | -0.05 [-0.68, 0.53] | 0.23 [-0.51, 0.94] | 0.28 [-0.58, 1.13] | 0.76 |
| Deep sleep | Percent-age points | -4°C | -0.10 [-0.57, 0.42] | -0.26 [-0.82, 0.24] | -0.17 [-0.92, 0.46] | 0.66 |
|  |  | -3°C | -0.17 [-0.54, 0.23] | -0.31 [-0.81, 0.12] | -0.14 [-0.78, 0.34] | 0.66 |
|  |  | -2°C | -0.18 [-0.45, 0.09] | -0.26 [-0.64, 0.04] | -0.08 [-0.53, 0.25] | 0.62 |
|  |  | -1°C | -0.12 [-0.26, 0.0] | -0.14 [-0.32, 0.0] | -0.02 [-0.23, 0.14] | 0.54 |
|  |  | 0°C ref |  |  |  |  |
|  |  | +1°C | 0.18 [0.06, 0.3] | 0.12 [-0.02, 0.26] | -0.06 [-0.22, 0.09] | 0.78 |
|  |  | +2°C | 0.39 [0.15, 0.65] | 0.21 [-0.17, 0.49] | -0.18 [-0.55, 0.11] | 0.87 |
|  |  | +3°C | 0.63 [0.27, 1.02] | 0.29 [-0.17, 0.69] | -0.34 [-0.92, 0.10] | 0.93 |
|  |  | +4°C | 0.89 [0.43, 1.38] | 0.39 [-0.16, 0.88] | -0.50 [-1.25, 0.10] | 0.94 |
|  |  | +5°C | 1.17 [0.56, 1.18] | 0.52 [-0.14, 1.15] | -0.17 [-1.58, 0.14] | 0.94 |
| REM sleep | Percent-age points | -4°C | 0.45 [-0.17, 0.98] | 0.95 [0.38, 1.56] | 0.50 [-0.18, 1.31] | 0.92 |
|  |  | -3°C | 0.39 [-0.08, 0.78] | 0.74 [0.28, 1.25] | 0.35 [-0.15, 0.98] | 0.91 |
|  |  | -2°C | 0.30 [-0.02, 0.57] | 0.51 [0.18, 0.88] | 0.21 [-0.13, 0.64] | 0.89 |
|  |  | -1°C | 0.17 [0.02, 0.31] | 0.26 [0.10, 0.44] | 0.09 [-0.08, 0.29] | 0.86 |
|  |  | 0°C ref |  |  |  |  |
|  |  | +1°C | -0.21 [-0.35, -0.08] | -0.26 [-0.41, -0.11] | -0.05 [-0.22, 0.13] | 0.74 |
|  |  | +2°C | -0.44 [-0.74, -0.18] | -0.50 [-0.82, -0.19] | -0.06 [-0.39, 0.32] | 0.67 |
|  |  | +3°C | -0.68 [-1.13, -0.31] | -0.73 [-1.16, -0.27] | -0.04 [-0.52, 0.54] | 0.60 |
|  |  | +4°C | -0.93 [-1.49, -0.45] | -0.94 [-1.48, -0.37] | -0.01 [-0.63, 0.77] | 0.54 |
|  |  | +5°C | -1.18 [-1.91, -0.56] | -1.15 [-1.82, -0.42] | 0.04 [-0.77, 1.04] | 0.50 |
| Recovery index | Recove-ry  index units | -4°C | 0.02 [0.00, 0.03] | 0.00 [-0.01, 0.02] | -0.01 [-0.03, 0.00] | 0.95 |
|  |  | -3°C | 0.01 [0.00, 0.02] | 0.00 [-0.01, 0.02] | -0.01 [-0.03, 0.00] | 0.96 |
|  |  | -2°C | 0.01 [0.00, 0.02] | 0.00 [-0.01, 0.01] | -0.01 [-0.02, 0.001] | 0.97 |
|  |  | -1°C | 0.01 [0.00, 0.01] | 0.00 [0.00, 0.01] | 0.00 [-0.01, 0.0003] | 0.97 |
|  |  | 0°C ref |  |  |  |  |
|  |  | +1°C | -0.01 [-0.01, 0.00] | 0.00 [-0.01, 0.00] | 0.00 [0.00, 0.01] | 0.95 |
|  |  | +2°C | -0.01 [-0.02, -0.01] | -0.01 [-0.02, 0.001] | 0.01 [0.00, 0.02] | 0.93 |
|  |  | +3°C | -0.02 [-0.03, -0.01] | -0.01 [-0.02, 0.00] | 0.01 [0.00, 0.02] | 0.92 |
|  |  | +4°C | -0.03 [-0.04, -0.01] | -0.02 [-0.03, 0.00] | 0.01 [-0.01, 0.03] | 0.91 |
|  |  | +5°C | -0.03 [-0.05, -0.02] | -0.02 [-0.04, 0.001] | 0.01 [-0.01, 0.04] | 0.89 |
| Subjective sleep quality, BPSQI-adjusted | scale points | -4°C | -0.20 [-0.43, 0.03] | -0.37 [-0.59, -0.14] | -0.16 [-0.42, 0.11] | 0.90 |
|  |  | -3°C | -0.15 [-0.33, 0.05] | -0.26 [-0.45, -0.06] | -0.11 [-0.31, 0.10] | 0.87 |
|  |  | -2°C | -0.10 [-0.24, 0.04] | -0.17 [-0.31, -0.02] | -0.07 [-0.20, 0.09] | 0.85 |
|  |  | -1°C | -0.06 [-0.13, 0.01] | -0.10 [-0.16, -0.02] | -0.04 [-0.10, 0.04] | 0.85 |
|  |  | 0°C ref |  |  |  |  |
|  |  | +1°C | 0.11 [0.05, 0.17] | 0.15 [0.08, 0.21] | 0.04 [-0.03, 0.10] | 0.89 |
|  |  | +2°C | 0.28 [0.15, 0.40] | 0.37 [0.23, 0.50] | 0.09 [-0.05, 0.22] | 0.92 |
|  |  | +3°C | 0.50 [0.33, 0.68] | 0.65 [0.47, 0.84] | 0.15 [-0.05, 0.35] | 0.94 |
|  |  | +4°C | 0.77 [0.55, 1.00] | 0.99 [0.77, 1.21] | 0.22 [-0.03, 0.49] | 0.96 |
|  |  | +5°C | 1.07 [0.77, 1.35] | 1.35 [1.06, 1.64] | 0.28 [-0.04, 0.67] | 0.96 |

**Panel B. Mean nocturnal wet-bulb temperature**

| **Outcome** | **Unit** | **Air temp. deviation** | **Control: difference from usual [95% CrI]** | **Treatment: difference from usual [95% CrI]** | **Treatment - Control difference [95% CrI]** | **P**  **(dir)** |
| --- | --- | --- | --- | --- | --- | --- |
| Sleep maintenance efficiency | Percent-age points | -4°C | 0.00 [-0.20, 0.17] | -0.02 [-0.21, 0.18] | -0.02 [-0.24, 0.24] | 0.57 |
|  |  | -3°C | 0.02 [-0.12, 0.15] | 0.01 [-0.14, 0.18] | -0.01 [-0.17, 0.19] | 0.56 |
|  |  | -2°C | 0.03 [-0.07, 0.12] | 0.02 [-0.08, 0.14] | 0.00 [-0.12, 0.13] | 0.55 |
|  |  | -1°C | 0.02 [-0.03, 0.07] | 0.02 [-0.04, 0.08] | 0.00 [-0.06, 0.06] | 0.57 |
|  |  | 0°C ref |  |  |  |  |
|  |  | +1°C | -0.03 [-0.08, 0.02] | -0.02 [-0.08, 0.04] | 0.01 [-0.06, 0.07] | 0.63 |
|  |  | +2°C | -0.06 [-0.16, 0.04] | -0.03 [-0.15, 0.08] | 0.03 [-0.10, 0.15] | 0.69 |
|  |  | +3°C | -0.10 [-0.25, 0.03] | -0.04 [-0.19, 0.11] | 0.06 [-0.12, 0.25] | 0.75 |
|  |  | +4°C | -0.15 [-0.37, 0.04] | -0.05 [-0.26, 0.16] | 0.10 [-0.14, 0.39] | 0.78 |
|  |  | +5°C | -0.21 [-0.53, 0.06] | -0.06 [-0.36, 0.25] | 0.15 [-0.19, 0.59] | 0.79 |
| Total sleep time | min | -4°C | 6.23 [1.33, 10.85] | 5.23 [0.05, 11.02] | -1.00 [-7.01, 6.38] | 0.65 |
|  |  | -3°C | 5.19 [1.53, 8.88] | 3.94 [-0.21, 8.60] | -1.25 [-5.85, 4.06] | 0.72 |
|  |  | -2°C | 3.64 [1.21, 6.09] | 2.28 [-0.65, 5.20] | -1.36 [-4.58, 1.82] | 0.81 |
|  |  | -1°C | 1.83 [0.60, 3.03] | 0.83 [-0.76, 2.26] | -1.00 [-2.84, 0.46] | 0.90 |
|  |  | 0°C ref |  |  |  |  |
|  |  | +1°C | -1.88 [-3.30, -0.30] | -0.46 [-2.18, 1.69] | 1.42 [-0.27, 3.93] | 0.94 |
|  |  | +2°C | -4.16 [-6.95, -1.23] | -1.45 [-4.64, 2.58] | 2.71 [-0.59, 7.48] | 0.93 |
|  |  | +3°C | -7.15 [-11.10, -3.55] | -3.74 [-7.78, 0.87] | 3.41 [-1.22, 9.29] | 0.91 |
|  |  | +4°C | -10.83 [-16.26, -5.90] | -7.38 [-12.65, -1.92] | 3.46 [-3.14, 10.66] | 0.85 |
|  |  | +5°C | -14.90 [-23.06, -7.65] | -11.77 [-20.20, -3.84] | 3.13 [-7.12, 12.79] | 0.76 |
| Light sleep | Percent-age points | -4°C | -0.17 [-0.70, 0.33] | -0.20 [-0.78, 0.38] | -0.04 [-0.70, 0.65] | 0.55 |
|  |  | -3°C | -0.14 [-0.58, 0.24] | -0.16 [-0.63, 0.32] | -0.02 [-0.53, 0.53] | 0.54 |
|  |  | -2°C | -0.11 [-0.42, 0.14] | -0.12 [-0.45, 0.20] | -0.01 [-0.35, 0.37] | 0.54 |
|  |  | -1°C | -0.06 [-0.22, 0.07] | -0.07 [-0.23, 0.09] | -0.01 [-0.18, 0.19] | 0.54 |
|  |  | 0°C ref |  |  |  |  |
|  |  | +1°C | 0.05 [-0.09, 0.22] | 0.06 [-0.12, 0.25] | 0.01 [-0.18, 0.20] | 0.57 |
|  |  | +2°C | 0.06 [-0.22, 0.37] | 0.10 [-0.23, 0.45] | 0.04 [-0.32, 0.41] | 0.59 |
|  |  | +3°C | 0.01 [-0.40, 0.40] | 0.09 [-0.36, 0.55] | 0.08 [-0.44, 0.62] | 0.62 |
|  |  | +4°C | -0.07 [-0.68, 0.45] | 0.05 [-0.56, 0.66] | 0.13 [-0.58, 0.91] | 0.63 |
|  |  | +5°C | -0.18 [-1.12, 0.54] | 0.00 [-0.92, 0.87] | 0.18 [-0.80, 1.32] | 0.63 |
| Deep sleep | Percent-age points | -4°C | 0.02 [-0.47, 0.54] | -0.19 [-0.71, 0.31] | -0.21 [-0.92, 0.42] | 0.72 |
|  |  | -3°C | -0.01 [-0.39, 0.40] | -0.15 [-0.58, 0.26] | -0.13 [-0.70, 0.35] | 0.68 |
|  |  | -2°C | -0.05 [-0.30, 0.24] | -0.11 [-0.41, 0.16] | -0.07 [-0.45, 0.25] | 0.63 |
|  |  | -1°C | -0.05 [-0.17, 0.09] | -0.07 [-0.23, 0.07] | -0.02 [-0.21, 0.14] | 0.56 |
|  |  | 0°C ref |  |  |  |  |
|  |  | +1°C | 0.14 [-0.01, 0.28] | 0.10 [-0.07, 0.28] | -0.04 [-0.21, 0.17] | 0.68 |
|  |  | +2°C | 0.37 [0.11, 0.65] | 0.20 [-0.11, 0.52] | -0.16 [-0.52, 0.20] | 0.83 |
|  |  | +3°C | 0.68 [0.30, 1.08] | 0.28 [-0.12, 0.68] | -0.39 [-0.93, 0.09] | 0.94 |
|  |  | +4°C | 1.05 [0.47, 1.63] | 0.34 [-0.22, 0.88] | -0.71 [-1.53, 0.04] | 0.97 |
|  |  | +5°C | 1.45 [0.59, 2.34] | 0.38 [-0.47, 1.19] | -1.07 [-2.34, 0.03] | 0.97 |
| REM sleep | Percent-age points | -4°C | 0.40 [-0.23, 0.98] | 0.45 [-0.16, 1.05] | 0.05 [-0.68, 0.83] | 0.54 |
|  |  | -3°C | 0.36 [-0.12, 0.79] | 0.32 [-0.19, 0.79] | -0.04 [-0.62, 0.53] | 0.57 |
|  |  | -2°C | 0.31 [0.01, 0.58] | 0.24 [-0.10, 0.56] | -0.06 [-0.45, 0.31] | 0.63 |
|  |  | -1°C | 0.19 [0.05, 0.34] | 0.16 [-0.01, 0.32] | -0.04 [-0.23, 0.15] | 0.66 |
|  |  | 0°C ref |  |  |  |  |
|  |  | +1°C | -0.27 [-0.46, -0.11] | -0.22 [-0.43, -0.04] | 0.04 [-0.18, 0.25] | 0.68 |
|  |  | +2°C | -0.56 [-0.95, -0.25] | -0.45 [-0.84, -0.10] | 0.12 [-0.30, 0.53] | 0.73 |
|  |  | +3°C | -0.84 [-1.35, -0.41] | -0.59 [-1.07, -0.14] | 0.25 [-0.31, 0.85] | 0.81 |
|  |  | +4°C | -1.09 [-1.73, -0.52] | -0.65 [-1.25, -0.05] | 0.44 [-0.32, 1.29] | 0.86 |
|  |  | +5°C | -1.32 [-2.18, -0.55] | -0.66 [-1.53, 0.30] | 0.66 [-0.39, 1.93] | 0.88 |
| Recovery index | Recove-ry  index units | -4°C | 0.01 [-0.01, 0.02] | 0.00 [-0.01, 0.02] | -0.01 [-0.03, 0.02] | 0.73 |
|  |  | -3°C | 0.01 [0.00, 0.02] | 0.00 [-0.01, 0.01] | -0.01 [-0.02, 0.01] | 0.85 |
|  |  | -2°C | 0.01 [0.00, 0.01] | 0.00 [-0.01, 0.01] | -0.01 [-0.02, 0.00] | 0.94 |
|  |  | -1°C | 0.00 [0.00, 0.01] | 0.00 [-0.01, 0.00] | -0.01 [-0.01, 0.00] | 0.97 |
|  |  | 0°C ref |  |  |  |  |
|  |  | +1°C | -0.01 [-0.01, 0.00] | 0.00 [0.00, 0.01] | 0.01 [0.0001, 0.02] | 0.98 |
|  |  | +2°C | -0.01 [-0.02, 0.00] | 0.00 [-0.01, 0.01] | 0.01 [0.00, 0.03] | 0.97 |
|  |  | +3°C | -0.02 [-0.03, -0.01] | -0.01 [-0.02, 0.01] | 0.01 [0.00, 0.03] | 0.93 |
|  |  | +4°C | -0.03 [-0.04, -0.01] | -0.02 [-0.04, 0.0002] | 0.01 [-0.01, 0.03] | 0.81 |
|  |  | +5°C | -0.03 [-0.06, -0.01] | -0.03 [-0.06, 0.00] | 0.01 [-0.03, 0.04] | 0.64 |
| Subjective sleep quality, BPSQI-adjusted | scale points | -4°C | -0.29 [-0.52, -0.06] | -0.49 [-0.74, -0.25] | -0.19 [-0.50, 0.09] | 0.91 |
|  |  | -3°C | -0.24 [-0.42, -0.06] | -0.39 [-0.60, -0.20] | -0.15 [-0.39, 0.07] | 0.91 |
|  |  | -2°C | -0.17 [-0.29, -0.05] | -0.26 [-0.40, -0.13] | -0.09 [-0.24, 0.07] | 0.88 |
|  |  | -1°C | -0.09 [-0.15, -0.03] | -0.12 [-0.19, -0.05] | -0.03 [-0.10, 0.05] | 0.81 |
|  |  | 0°C ref |  |  |  |  |
|  |  | +1°C | 0.12 [0.04, 0.19] | 0.14 [0.04, 0.22] | 0.02 [-0.10, 0.10] | 0.71 |
|  |  | +2°C | 0.28 [0.14, 0.42] | 0.34 [0.16, 0.50] | 0.06 [-0.16, 0.21] | 0.77 |
|  |  | +3°C | 0.52 [0.34, 0.70] | 0.66 [0.45, 0.85] | 0.14 [-0.11, 0.36] | 0.89 |
|  |  | +4°C | 0.81 [0.55, 1.06] | 1.07 [0.83, 1.32] | 0.26 [-0.03, 0.59] | 0.96 |
|  |  | +5°C | 1.13 [0.73, 1.52] | 1.54 [1.17, 1.94] | 0.41 [-0.02, 0.96] | 0.97 |

Posterior expected differences in sleep outcomes were estimated for within-person nocturnal temperature deviations relative to 0°C, representing each participant’s usual exposure, separately for the Control and Treatment groups. The Treatment - Control difference represents the difference between these group-specific temperature contrasts and thus quantifies modification of the temperature-sleep association by intervention assignment. For beta-regression outcomes, differences are reported in percentage points; for Gaussian outcomes, differences are reported in native outcome units. Values are posterior means with 95% credible intervals (CrI). P(dir) denotes the posterior probability that the Treatment - Control difference is in the estimated direction.

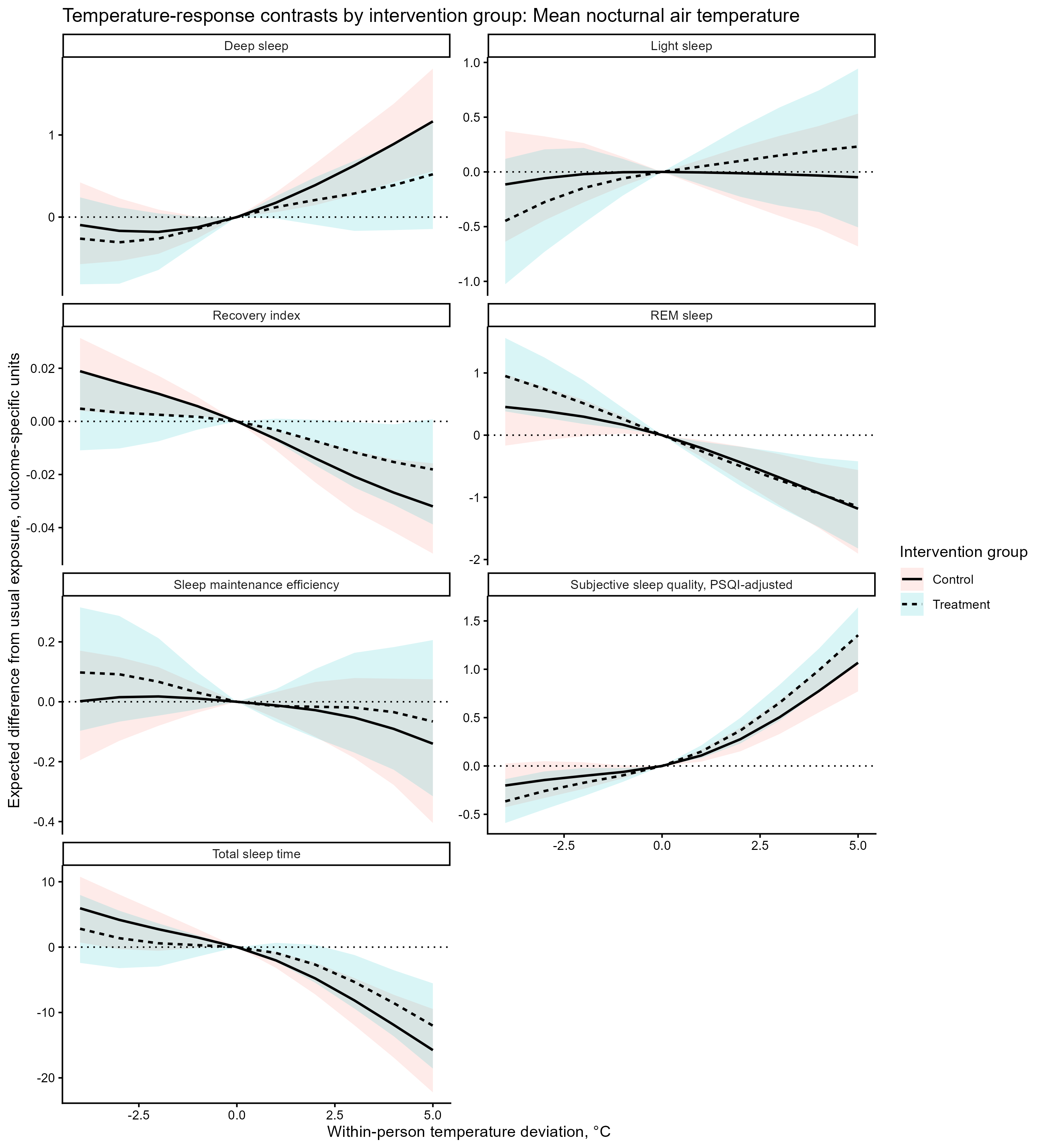

**Supplementary Figure S9. Posterior temperature-response contrasts by intervention group for mean nocturnal air temperature.** Solid lines show posterior mean estimates for the control group, whereas dashed lines show the estimates for the treatment group. Shaded areas indicate 95% credible intervals (red = control group, blue = treatment group). For the within-person analyses, 0°C represents a participant’s usual nocturnal temperature exposure. Differences are expressed in percentage points for sleep maintenance efficiency and light, deep, and REM sleep; minutes for total sleep time; recovery-index units for recovery index; and scale points for subjective sleep quality. Higher subjective sleep-quality scores indicate worse sleep quality. Participant-specific random effects were excluded from predictions, yielding population-level expected contrasts.

Group Differences

**Supplementary Table S21**. **Adjusted overall differences in sleep outcomes between the Treatment and Control groups.**

| **Outcome** | **N, Control/ Treatment** | **Control, adjusted mean [95% CrI]** | **Treatment, adjusted mean [95% CrI]** | **Treatment - Control difference [95% CrI]** | **P**  **(dir)** |
| --- | --- | --- | --- | --- | --- |
| Sleep maintenance efficiency | 145/156 | 96.16 [95.87, 96.42] | 96.34 [96.07, 96.59] | 0.18 [-0.19, 0.56] | 0.83 |
| Total sleep time | 145/156 | 424.64 [417.08, 432.24] | 426.82 [419.38, 434.12] | 2.18 [-7.94, 12.75] | 0.65 |
| Light sleep | 145/156 | 62.56 [61.61, 63.49] | 63.22 [62.29, 64.13] | 0.66 [-0.67, 1.97] | 0.84 |
| Deep sleep | 145/156 | 19.41 [18.62, 20.2] | 19.03 [18.27, 19.8] | -0.38 [-1.47, 0.76] | 0.75 |
| REM sleep | 145/156 | 17.23 [16.36, 18.13] | 16.39 [15.55, 17.24] | -0.84 [-2.02, 0.38] | 0.92 |
| Recovery index | 140/147 | 0.54 [0.52, 0.57] | 0.55 [0.53, 0.57] | 0.01 [-0.03, 0.04] | 0.65 |
| Subjective Sleep quality, B-PSQI-adjusted | 156/162 | 12.18 [11.87, 12.51] | 12.47 [12.17, 12.77] | 0.29 [-0.16, 0.73] | 0.89 |

Values are posterior expected means and Treatment-minus-Control differences with 95% credible intervals (CrI). Group means were standardised over the observed distribution of mean nocturnal air temperature and the remaining covariates in the eligible analysis sample, such that the same observations were predicted under the Control and Treatment conditions and subsequently averaged with equal participant weighting. Models were adjusted for within- and between-person mean nocturnal air temperature, study day, age, and gender; models of subjective sleep quality additionally adjusted for baseline B-PSQI. Differences are expressed in percentage points for sleep maintenance efficiency and sleep-stage proportions, minutes for total sleep time, recovery-index units for recovery index, and scale points for subjective sleep quality. Higher subjective sleep-quality scores indicate worse sleep quality. Positive Treatment-minus-Control values indicate higher values in the Treatment group. P(direction) denotes the posterior probability that the estimated group difference is in the observed direction.
